# Effects of interdisciplinary early developmental intervention programs on behavior, executive functioning and participation in children born preterm: A systematic review with meta-analysis

**DOI:** 10.64898/2026.06.02.26354617

**Authors:** Leonie Schirle, Marie Babel, Jana-Susann Briem, Nina Gawehn, Heidrun Janka, Maria-Inti Metzendorf, Eva Trunk, Judith Wohlleben, Stephanie Weibel, Juliane Spiegler

## Abstract

**Aim:** To systematically evaluate evidence on the effects of post-discharge early developmental intervention programs (EI) on behavioral development, quality of life, participation, executive functioning, parent–child interaction, and use of medical services from infancy through adolescence in children born preterm.

**Method:** Four bibliographic databases and one trial registry were systematically searched for randomized controlled trials up to April 23, 2024. Two reviewers independently screened studies and extracted data. In clinically and methodologically comparable studies, random-effects meta-analysis were performed. Risk of bias was assessed with the Cochrane RoB 2 tool, and certainty of evidence with the GRADE approach.

**Results:** Twenty-six studies met inclusion criteria, eleven studies including 2,315 preterm born infants reported relevant outcomes, and seven contributed to meta-analyses. Most reported results showed some concerns or high risk of bias; certainty of evidence ranged from very low to moderate across outcomes. EI may offer small benefits for selective attention, behavioral problems and parent-child interaction. Little to no effect was found for special educational needs, language skills, executive functioning and the use of medical services. No included studies evaluated the effect of EI on ADHD, quality of life, or participation related to mobility or leisure activities.

**Interpretation:** EI may improve problems typically seen in preterm children and should be offered especially to those with additional medical or social risk factors. High-quality, contemporary trials are needed to establish reliable clinical recommendations regarding EI strategies and complementary interventions throughout childhood.

## 1. Introduction

Given that in 2020, approximately one in ten babies worldwide was born prematurely [1], preterm birth remains a global health concern. Despite advances in neonatal care, complications caused by prematurity are still the leading cause for mortality in children under five years of age [2]. Additionally, premature birth is associated with adverse long-term morbidity like neurodevelopmental delay [3], behavioral and psychiatric disorders [4–6], impaired quality of life [7], and a higher need in special educational support [4, 8]. Data concerning the effect of preterm birth on executive functioning (EF) are inconclusive [9]. The severity of developmental impairments increases with decreasing gestational age [9–11]. With continuously improving survival rates of premature babies, the prevalence of neurodevelopmental disorders increases [12]. For example, despite continuous improvements in neonatal care over the past 30 years, cognitive development following premature birth has remained unchanged [13]. This emphasizes the need to improve effective strategies for post-discharge developmental care in this growing population with special needs. One effective strategy to improve developmental outcomes and participation in premature infants may be early developmental intervention programs (EI).

EI are interdisciplinary concepts to support infants and toddlers in their neurological and social development and participation. This includes the early detection and, if possible, reduction of potential developmental disorders through elements of physical therapy, occupational therapy and speech therapy, as well as advice and educational support for parents and the stimulation of an environment that promotes participation of the child. There is evidence that EI has a positive effect on cognitive outcomes up to preschool age, whereas positive effects on motor outcomes are limited to infancy and do not persist at later ages [14]. Despite these positive effects on cognition, in Germany EI is mostly implemented as tertiary prevention but rarely as secondary prevention measure [15]. Notably, the likelihood of EI utilization correlates with the active recommendation of such interventions by discharging neonatal clinics [15]. This highlights the need for standardized guidelines on recommendation and implementation of EI.

This work is part of an evidence-based guideline development for post-discharge follow-up care of preterm born infants in Germany. With respect to the German health care system the guideline group (see acknowledgements) deemed the following 5 interventions as highest priority to initiate systematic reviews:

In preterm born infants born before 37 weeks of gestation compared to standard of care does

1. Intervention: interdisciplinary early intervention programs (EI)
2. Intervention: isolated physical therapy (0-6 years)
3. Intervention: isolated occupational therapy (3-6 years)
4. Intervention: early entry in professional childcare (before age 3 years versus later)
5. Intervention: School entry (timely versus late)

improve the following outcomes (if appropriate for the intervention): cognitive development, motor development, parent-child-interaction, executive functioning, behavior, quality of life, participation (focus on communication, leisure activity, special educational needs).

This work addresses the first PICO “EI”. Despite growing evidence that EI benefits early motor and longer-term cognitive development [14], important knowledge gaps remain regarding its impact on parent–child interaction, EF, behavioral development, quality of life, and participation.

## 2. Methods

A systematic review was conducted to address the above-mentioned research question. The corresponding protocol was registered in advance (PROSPERO: CRD42024562388).

### 2.1 Eligibility criteria

#### 2.1.1 Types of studies

To mitigate the risk of confounding, only randomized controlled trials (RCTs) were included. Eligible studies include those that were published or reported in trial registries as still ongoing or completed but unpublished (‘awaiting classification’). Publications with insufficient information for reliable inclusion also were categorized as ‘awaiting classification’. In such cases, study authors were contacted for clarification. Publications in languages other than English or German were translated using Google Translate.

#### 2.1.2 Participants / Population

We included studies investigating:

- Children born preterm (<37 weeks gestational age (GA), regardless of birth weight)
- Children born preterm AND with low birth weight (LBW <2500g)
- Children born preterm AND/OR with very low birth weight (VLBW <1500g)
- Children born preterm AND/OR with extremely low birth weight (ELBW <1000g)

Studies involving mixed populations were eligible if subgroup data based on gestational age were available.

If the population consisted of full-term infants (≥ 37 weeks GA) or of low-birth-weight infants without specific information on gestational age at birth, studies were excluded.

Eligibility criteria for participants applied to both the intervention and comparison groups.

#### 2.1.3 Intervention(s)

Eligible EI could include, but are not limited to, physical therapy, occupational therapy, speech therapy, neurodevelopmental interventions, multisensory stimulation, home support programs, and educational-behavioral interventions for caregivers. The interventions had to be either developed by an interdisciplinary team (e.g. IBAIP) or implemented by at least two different healthcare disciplines. They had to be provided in either a hospital or at home or at a public institution. Interventions were bound to commence within the first year of life, had to target the child directly, and could include parental participation. They also could begin during the primary hospital stay but had to include a minimum of six sessions following discharge.

#### 2.1.4 Comparator(s)

Eligible comparators were:

- no treatment
- standard of care, e.g. follow-up care by pediatricians
- active comparators, e.g. any other type or mode of EI methods and concepts

Standard of care had to be comparable between study groups.

#### 2.1.5 Outcomes

The main outcome set comprised:

- Behavior: Attention Deficit/Hyperactivity Disorder (ADHD)
- Quality of life
- Participation: leisure activities, special educational needs (SEN)
- Executive functioning
- Parent-child interaction (PCI)

As additional outcomes we included:

- Behavior: autism spectrum disorder (ASD); internalizing problems (e.g. anxiety, depression)
- Participation: communication, mobility
- Frequency of medical services after initial therapy

The outcomes of behavior, quality of life, and PCI were required to be assessed at or beyond six months corrected age, while participation and EF assessments were to be conducted no earlier than four years corrected age. Because of the different stages in a child’s development, outcomes were pooled according to predefined age groups as follows:

- infancy: age 0-3 years
- preschool age: 4-6 years
- early school age: 7-12 years
- adolescence: 13-18 years

After publication of the protocol, we decided to extract data exclusively from the latest time point if outcomes were measured multiple times during the same age group and using the same instrument within one study.

Eligible studies that did not cover at least one of the predefined outcomes were excluded from data extraction but are reported separately in the supplementary materials S1. All excluded studies and the reason for exclusion are reported in the supplementary material S2.

For certain outcomes, specific assessment instruments were predefined. However, due to the limited number of results, other instruments were evaluated for eligibility after completion of the screening process by experienced clinicians (JS, NG). All assessment tools included are described in the supplementary material S3, as well as a summary of findings concerning the primary and secondary outcomes (supplementary material S4).

### 2.2 Search methods

A systematic search was conducted using the following bibliographic databases and trial registries from date of database inception to April 23, 2024: MEDLINE, PsycINFO, and CINAHL, Cochrane Central Register of Controlled Trials, and the WHO International Clinical Trials Registry Platform (ICTRP). Database-specific search filters were applied to identify RCTs [16–18]. Detailed search strategies are provided in the supplementary material S5. Additionally, reference lists of all included primary studies and identified systematic reviews were screened for further eligible studies.

All publications identified as ongoing or completed but unpublished in April 2024 were searched again on June 18, 2025, for updates.

According to the defined eligibility criteria and the Cochrane Handbook for Systematic Reviews of Interventions [19], two reviewers independently screened the titles and abstracts of all identified records. Full-text articles of potentially eligible studies were retrieved and independently assessed by two reviewers. Disagreements at either stage were resolved through discussion or, if necessary, consultation of a third reviewer.

Screening was conducted using the Covidence® systematic review software.[20]

### 2.3 Data extraction

Two reviewers independently extracted data on study characteristics, population characteristics, interventions, comparators, and outcomes using a piloted data extraction template. Further details of the data collection process are provided in the protocol. Differences between reviewers were addressed through discussion. When necessary, additional information was sought from the study authors.

Data extraction was executed with Covidence® for study characteristics and more detailed for the outcome results in Excel 365 (Version 16.100.1; 2025) [21].

### 2.4 Risk of bias assessment

Two reviewers independently assessed the risk of bias (RoB) of each extracted outcome using the Cochrane Risk of Bias 2 (RoB2) tool [22]. The risk of bias was classified as low, some concerns or high for each individual RoB domain and the overall RoB. Disagreements were resolved via discussion or consultation of a third reviewer.

Deviating from the protocol, also the secondary outcomes were assessed for risk of bias.

### 2.5 Data synthesis

Meta-analysis was conducted for clinical and methodological sufficiently homogenous studies. Patient age at intervention, the outcome assessment tools, and their scales had to be comparable.

Binary outcomes were analyzed using the Mantel-Haenszel method under a random-effects model to report pooled risk ratios (RR) with 95% confidence interval (95%-CI).

For continuous outcomes, mean difference (MD) with 95% CIs for results of studies referring to one scale, and standardized mean difference (SMD) with 95% CIs for results of studies referring to different scales were used. Data reported only as medians were converted to means if the distribution was normal; otherwise, they were not used for analysis. Meta-analysis was performed using the inverse variance method under a random-effects model. We used R package meta version 7.0-0 for analysis.

Statistical heterogeneity was evaluated using the χ^2^ test and the I^2^ statistic, with heterogeneity defined as P < 0.05 or an I^2^ value ≥ 40%. A 95% prediction interval (PI) was calculated for meta-analyses with at least four studies and ≥ 200 participants.

Subgroup and sensitivity analyses, as well as assessments of reporting bias, were considered not meaningful due to the limited number of included studies and were therefore not conducted.

### 2.6 Certainty of Evidence

The certainty of evidence (CoE) for each outcome eligible for data synthesis was independently assessed by two reviewers using the *Grading of Recommendations, Assessment, Development and Evaluations* (GRADE) approach. This framework categorizes the certainty of evidence into four levels: high, moderate, low, or very low. Evidence was downgraded by one to three levels based on the presence and severity of risk of bias, inconsistency, imprecision, indirectness, or suspected publication bias. Any disagreements between reviewers were resolved through discussion. The final ratings are presented in the Summary of Findings tables (supplementary material S4).

## 3. Results

### 3.1 Search

The search strategy identified 10,537 database records and 1,290 records in trial registries. After removing duplicates, 8,763 titles and abstracts were screened, with 8,180 deemed irrelevant. We sought 583 full-text articles for retrieval and assessed 574 for eligibility, along with eight additional entries from websites (**Error! Reference source not found**.).

A total of 407 reports were excluded. Forty-three records remained unclassified due to being unpublished or lacking retrievable data despite author contact. Twelve trials were still ongoing.

A total of 26 studies (107 full-text articles) met the inclusion criteria. However, only eleven RCTs provided outcome data suitable for synthesis, with seven eligible for meta-analysis. Fifteen studies could not be synthesized due to missing data, use of inappropriate assessment tools, or lack of prioritized outcomes.

### 3.2 Characteristics of included studies

A total of eleven studies with 2315 infants (mean GA 27-33 weeks) were eligible for data synthesis. They were published between 1990 and 2022 and conducted in the Republic of Korea [23], Taiwan [24], the Netherlands [25], France [26], Canada [27], the United States [28, 29], Brazil [30], Australia [31, 32], and the United Kingdom [33]. Further characteristics of the included studies can be found in the supplementary material S1.

Outcomes were assessed over a period ranging from three months to 13 years corrected age (CA) and included: PCI, special educational needs (participation), EF, problematic behavior, and internalizing behavior problems. In addition to the predefined outcomes, we examined attentional and externalizing behavior problems, as well as mother-to-child attachment.

Meta-analysis could be conducted for PCI, special educational needs, problematic behavior, internalizing and externalizing behavior problems. Additional non-poolable data were available for EF, attention deficits, use of medical services, and PCI.

A summary of all findings is presented in the results section, while a more detailed analysis of the predefined outcomes is provided in the supplementary material S4.

Additionally, a description of the assessment tools that were used in the included studies can be found in supplementary material S3.

### 3.3 Risk of bias (RoB)

Among the study results addressing the primary outcomes, 8.3% demonstrated low risk of bias, 33.3% were rated as ‘some concerns’, and 58.3% were assessed as having a high risk of bias (Table 1).

**Table 1.**
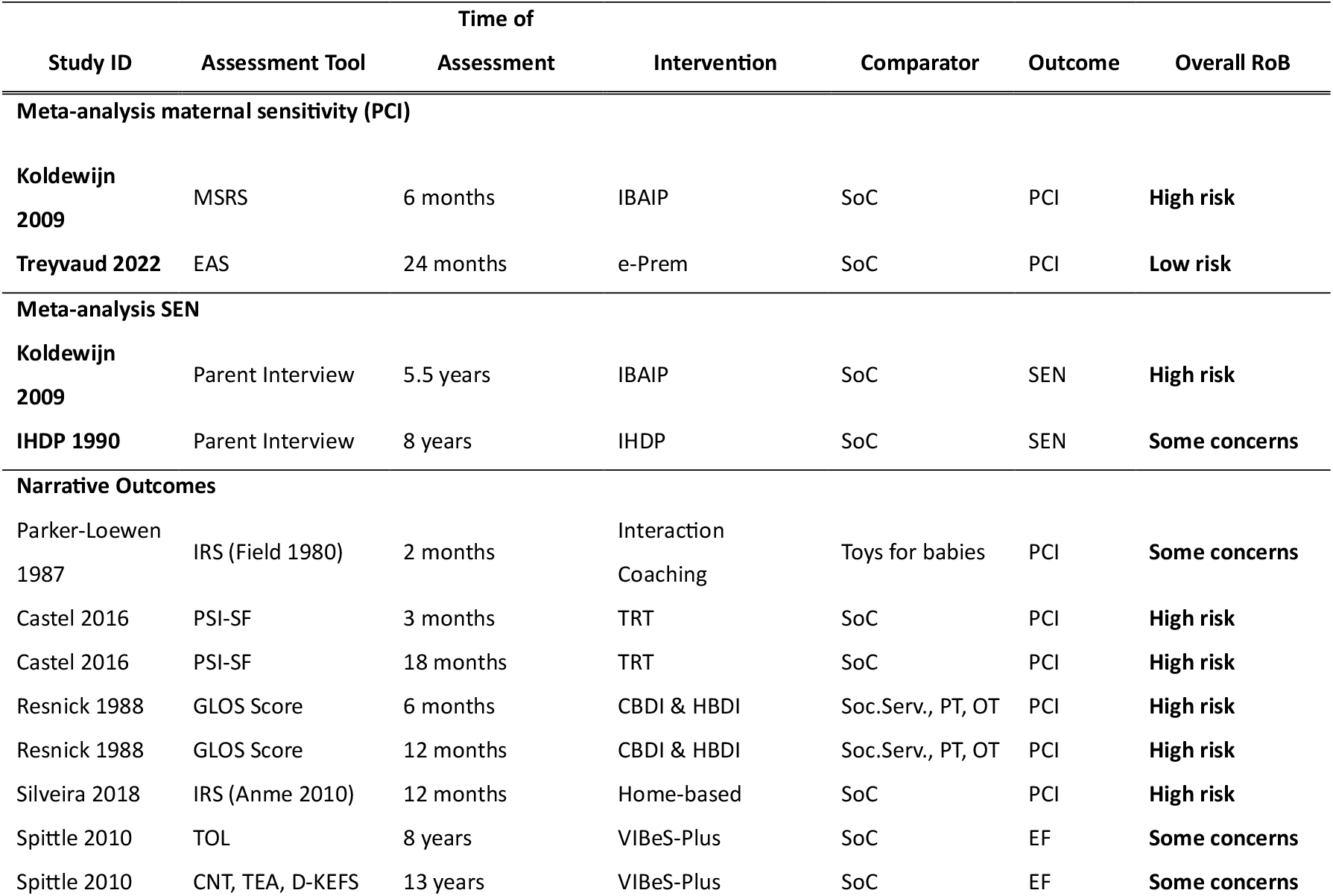
Risk of bias of primary outcomes.

For the study results related to secondary outcomes, 47.4% were rated as ‘high risk of bias’, and 52.6% were rated as having some concerns. No results were assessed as having a low risk of bias.

More details about the RoB assessment can be found in supplementary material S1.

### 3.4 Primary outcomes

#### Behavior

No studies reported the effect of EI on ADHD. However, attentional problems were assessed in one study including 81 participants (GA < 30 weeks) at 13 years of corrected age (CA). The study focused on child development and PCI, comparing the VIBeS Plus intervention with SoC. As assessment tool, the Test of Everyday Attention for children (TEA-Ch) was used.

We are uncertain about the effect of EI on attention. The results of the TEA-Ch indicate marginal effects for selective attention (n = 43; adjusted MD = 2.1 [95% CI: 0.2 – 4.0]) but not for other attention domains.[34] Certainty of evidence (CoE) was very low due to serious risk of bias, serious indirectness, and serious imprecision. Thus, the evidence is very uncertain about the effect of EI on attention.

#### Quality of life

No studies reported the effect of EI on Quality of life.

#### Participation

Special educational needs (SEN), as one aspect of participation, were assessed in two studies including 1011 participants (GA <36 weeks) at 5.5 to eight years CA. The studies focused on PCI [35] and child development [36], comparing the EI programs IBAIP [35] and IHDP [36] to SoC. As assessment tools, parent and teacher questionnaires were used.

We conducted meta-analysis for those studies (Figure 1). In deviation from the original protocol, we combined two distinct age groups, based on the assumption that the presence of SEN at one time point indicates a persistent condition. The results indicate that extensive EI probably has little to no effect on SEN at 5.5 and eight years CA (n = 1011; RR=0.91; 95%-CI [0.71; 1.16], I^2^=6.6%) [35, 36]. CoE was moderate due to high risk of bias.

**Figure 1.**
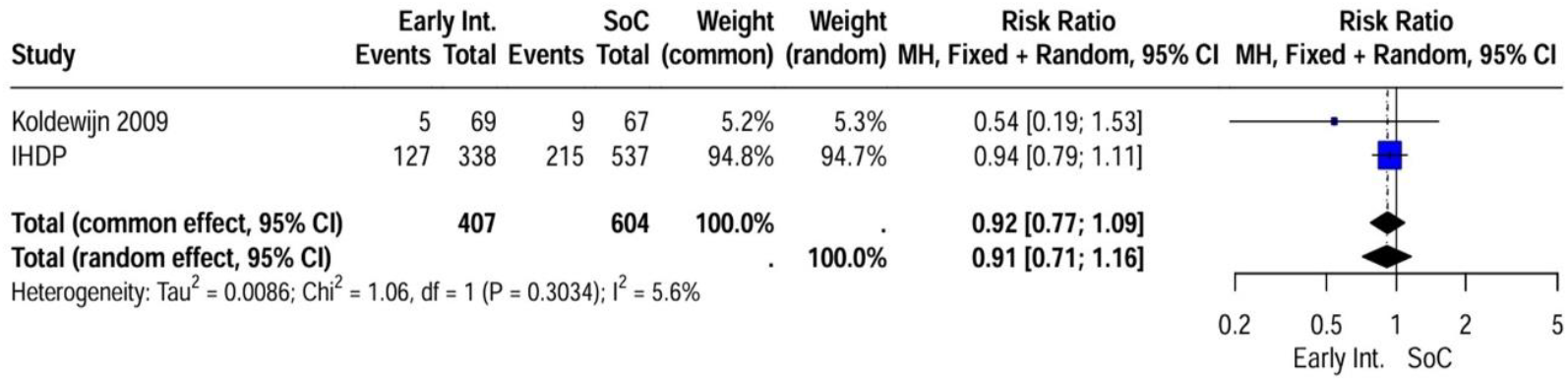
Special educational needs in preschool and school age

No studies reported the effect of EI on leisure activities.

#### Executive functioning

EF was assessed in one study including 81-100 participants (GA <34 weeks) at the age of 8 years [37] and 13 years [34]. The study focused on child development and PCI, comparing the VIBeS Plus intervention to SoC. At the 8-year assessment, the TOL and WMTB-C were used as assessment tools. At the 13-year assessment, the BRIEF2, WMTB-C, D-KEFS Tower Test, and CNT Arrow Switching were used as assessment tools. Because of the different age groups and as the results belong to the same study, no meta-analysis was conducted.

We are uncertain about the effect of EI on executive functioning. The results indicate little to no effect of EI on EF (Table Appendix B 1) but the evidence is very uncertain due to serious risk of bias and very serious imprecision.

#### Parent-child interaction

PCI was assessed in seven studies.

Maternal sensitivity, as one aspect of PCI, was assessed in two studies including 180 participants (GA < 34 weeks) at six to 24 months CA. The studies focused primarily on PCI, comparing the home-based EI program IBAIP [38] and the online parent-education program e-Prem [32] with SoC. As assessment tools, the Maternal Sensitivity and Responsivity Scale [38] and the Emotional Availability Scale [32] were used. We conducted a meta-analysis for these studies (Figure 2).

**Figure 2.**
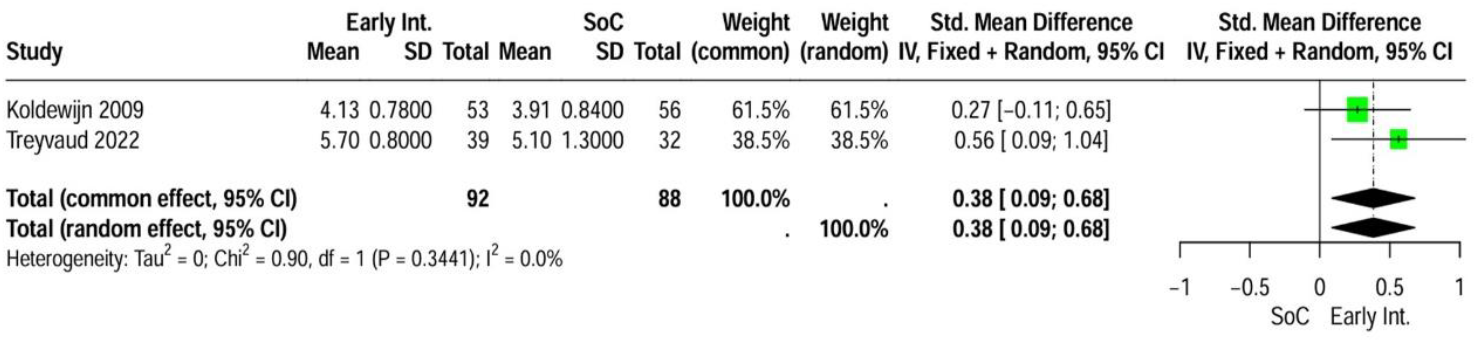
Maternal sensitivity in infancy (6-24 months CA)

The evidence suggests that EI may increase maternal sensitivity slightly (n = 180; SMD = 0.38 [95% CI: 0.09–0.68], I^2^ = 0.0%). CoE was low due to high risk of bias and serious imprecision.

Further outcomes related to PCI were assessed in five studies including 295 participants (GA < 37 weeks) at the age of 12 to 24 months CA. The studies focused on both child development and PCI, comparing several EI programs to SoC. Because of the use of non-comparable assessment tools across these studies, no meta-analysis was conducted.

Two studies implemented seven to nine sessions over a period of three to six months. As assessment tools the Interaction Rating Scale (Field, 1980) [27] and the Nijmeegse Ouderlijke Stress Index [39] were used. Three studies implemented 20 to >24 sessions over a period of 12 to 18 months. As assessment tools, the Parenting Stress Index – short form [26], the video-based Greenspan-Lieberman Observation System [29], and the Interaction Rating Scale (Anme, 2010) [40] were used.

We are uncertain about the effect of EI on PCI. The results indicate that short-term EI with less than ten interventions may have little to no effect while long-term interventions with ≥20 interventions may increase PCI, but the evidence is very uncertain due to very serious risk RoB and very substantial imprecision (Table Appendix B 2).

Mother-to-child attachment was assessed in two studies including 240 participants (GA < 35 weeks) at the age of 6-12 months CA. The studies focused on PCI, comparing a music therapy program [41] and the Mother-Infant Transaction Program based on physical therapy with SoC. As assessment tools, the Mother-to-Child Attachment Score [23] and the Postpartum Bonding Questionnaire [41] were used.

Because of insecure comparability of the assessment tools and the different intervention approaches, no meta-analysis was conducted. We are uncertain about the effect of EI on mother-to-child attachment. The results indicate that EI may have little to no effect on parent-child attachment (Table Appendix B ***2***), but the evidence is very uncertain due to serious risk of bias and very substantial imprecision.

### 3.4 Secondary outcomes

#### Behavior

Behavioral difficulties were assessed in four studies including 1268 participants (GA < 37 weeks) at the age of two to three years CA [24, 42] and at four years CA [43, 44] respectively. The studies focused on both child development and PCI, comparing different EI programs to SoC. As assessment tool, the age-appropriate versions of the Child Behavior Checklist were used.

We conducted meta-analyses for behavioral problems in infancy (Figure 3A), and preschool age (Figure 3B).

**Figure 3.**
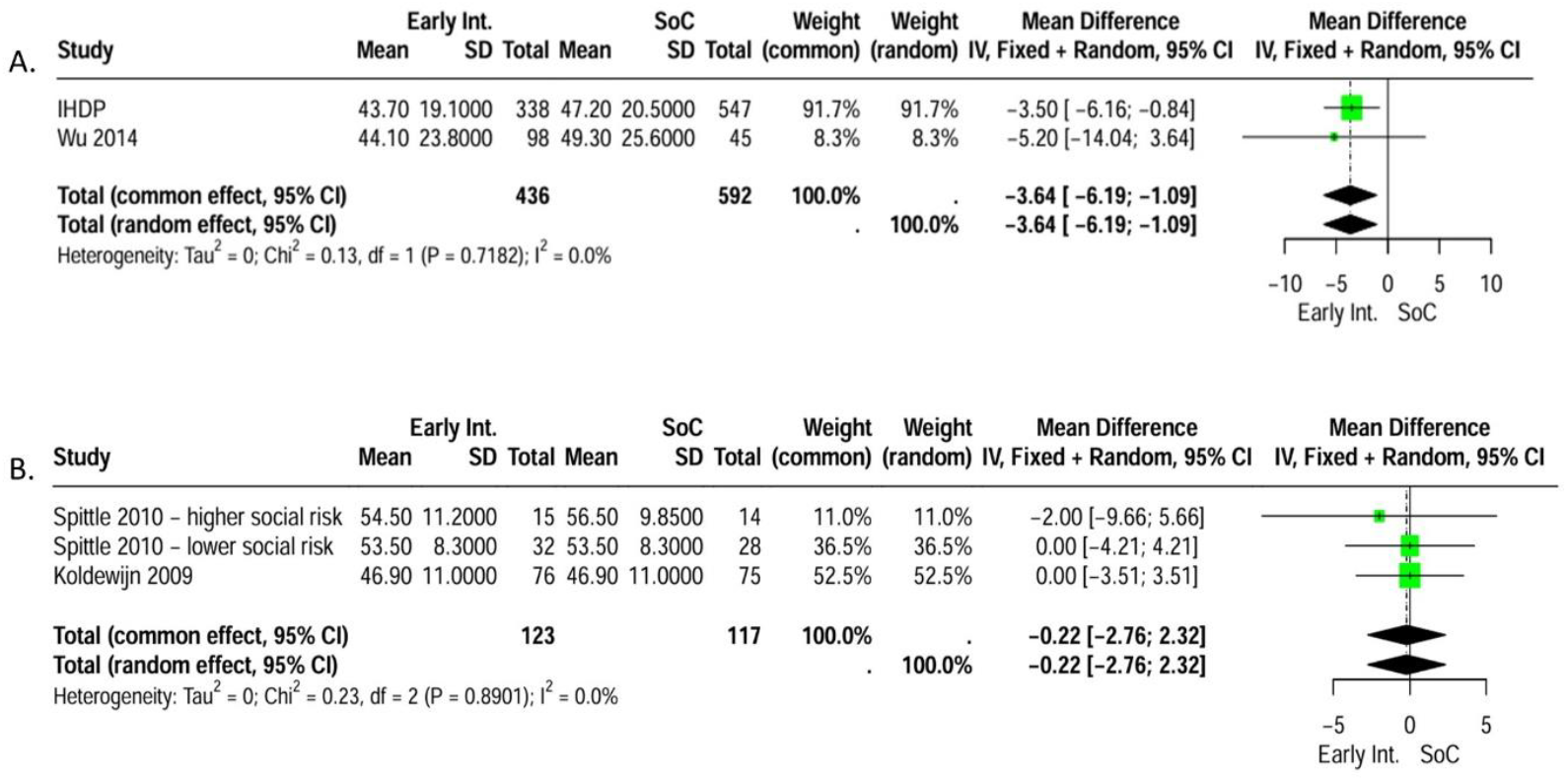
Problematic behavior in infancy (A) and preschool age (B)

In infancy, EI may reduce behavioral difficulties slightly (Figure 3A; n = 1028, MD = –3.64 [95% CI: –6.19 to –1.09], I^2^ = 0.0%) [24, 42]. CoE was low due to very severe risk of bias.

In preschool age, we are uncertain about the effect of EI on behavioral difficulties. EI may have little to no effect on behavioral difficulties (Figure 3B; n = 239, MD = –0.22 [95% CI: –2.76 to 2.32], I^2^ = 0.0%) [43, 44], but evidence is very uncertain due to serious risk of bias and imprecision.

Internalizing and externalizing behavior in infancy was assessed in four studies including 438 participants (GA <30 to <37 weeks) at the age of zero to three years CA [23, 24, 31, 32]. The studies, focusing on both child development and PCI, compared the effect of different EI programs to SoC: the VIBeS-Plus program [23, 31], a program based on physical therapy and implemented either at home or in the clinic ePREM [24], and the ePREM program [32]. As assessment tools, age-appropriate versions of the Child Behavior Checklist and the Infant-Toddler Social Emotional Assessment were used.

We conducted meta-analyses for these studies (Figure 4). However, we are uncertain about the effect of EI on internalizing and externalizing behavior in infancy. The results indicate little to no effect of EI on internalizing and externalizing behavior in infancy (internalizing: MD = – 0.17 [95% CI: –0.41 to 0.07], I^2^ = 34.4%; externalizing: MD = –0.05 [95% CI: –0.33 to 0.23], I^2^ = 52.2%) but evidence is very uncertain due to very serious RoB.

**Figure 4.**
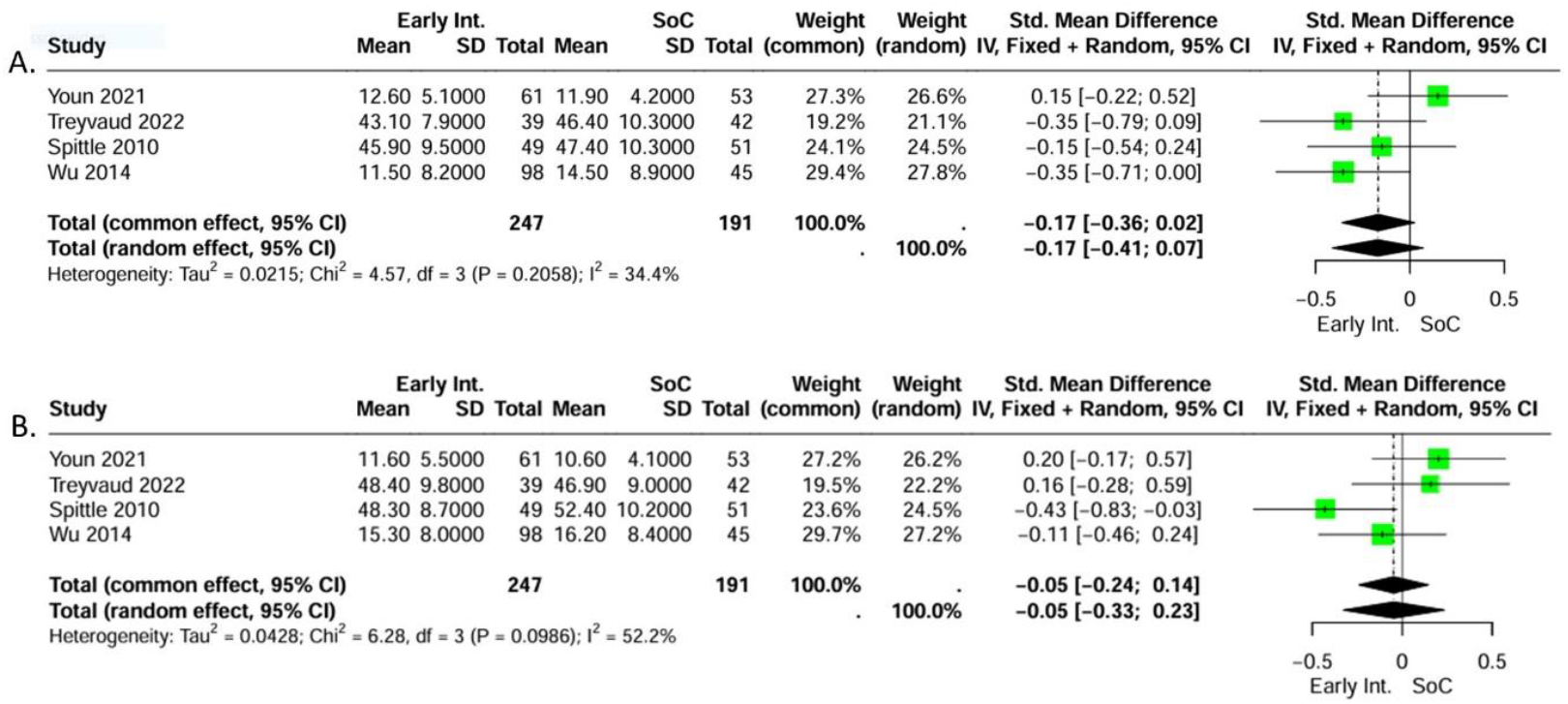
Internalizing (A) and externalizing (B) behavior in infancy

Internalizing and externalizing behavior in preschool age was assessed in two studies including 140 participants (GA <32 weeks) at four years CA [43, 44]. The studies, focusing primarily on PCI, compared the effect of the VIBeS-Plus program [43] and the IBAIP [44] to SoC. As assessment tools, age-appropriate versions of the Child Behavior Checklist were used. We conducted meta-analyses for these studies (Figure 5). We are uncertain about the effect of EI on internalizing and externalizing behavior.

**Figure 5.**
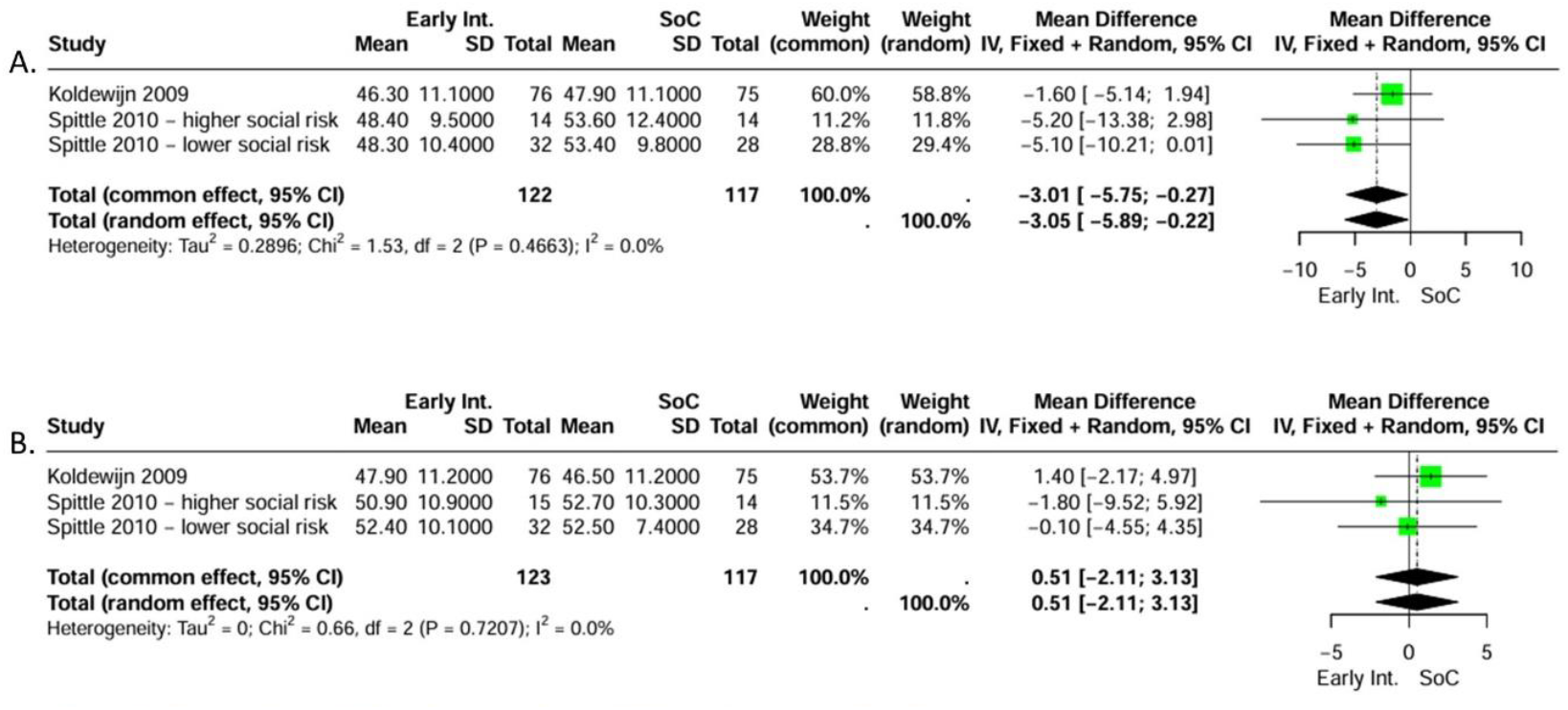
Internalizing (A) and externalizing (B) behavior in preschool age

The results indicate that EI may reduce internalizing but not externalizing behavior difficulties in preschool age (internalizing: MD = –3.05 [95% CI: –5.89 to –0.22], I^2^ = 0.0%; externalizing: MD = 0.51 [95% CI: –2.11 to 3.13], I^2^ = 0.0%) but the evidence is very uncertain due to very serious RoB.

Overall, EI may have little to no effect on internalizing or externalizing behavior. Nevertheless, one study examining socioeconomic subgroups found that, at age two, children from disadvantaged families that received EI showed greater improvements in externalizing behavior compared to those from lower risk backgrounds (higher social risk: MD = −8.7 [95% CI: −16.4 to −1.1]; lower social risk: MD = −0.9 [95% CI: –4.6 to 2.8]) [43]. No socioeconomic differences were observed for internalizing or overall behavioral difficulties at any age.

#### Participation

Communication skills as one aspect of participation were assessed in one study including 104 participants (GA < 30 weeks) at the age of four years CA. The study focused on both child development and PCI, comparing the VIBeS-Plus program to SoC. As assessment tool, the Differential Ability Scale was used.

We are uncertain about the effect of EI on communication skills. The results indicate that EI may have little to no effect on communication skills at four years CA (n = 104, MD = 4.1 [95% CI: –2.2 to 10.4]) [45] but evidence is very uncertain due to serious risk of bias and very serious imprecision. At age of eight years, evidence is still very uncertain about the effect of EI on communication skills due to very serious RoB and very substantial imprecision. But effects are more pronounced, particularly among children from socioeconomically disadvantaged backgrounds (higher social risk: MD = 9.0 [95% CI: −0.1 to 18.1]; lower social risk: MD = −3.1 [95% CI: −9.4 to 3.2]) [43].

No studies reported the effect of EI on mobility as one aspect of participation.

#### Use of medical services

The use of medical services was assessed in one study including 136 participants (GA < 32 weeks) at 5.5 years CA. The study focused on PCI, comparing the IBAIP to SoC. As assessment tools, parent interviews were used.

We are uncertain about the effect of EI on the use of medical services. The results indicate that EI may have little to no effect on usage of physical, occupational, or speech therapy (n = 136, MD = −0.039 [95%-CI −0.154 to 0.151]); EI also may have little to no effect on the need for psychological support in the intervention group during the preschool years (n = 136, MD = 0.026 [95%-CI −0.048 to 0.227]) [35]. But evidence is very uncertain due to very serious risk of bias and serious imprecision.

A summary of the results is provided in the supplementary materials S4.

## 4. Discussion

Prematurity and its long-term adverse consequences are a global health concern. Improvements in cognitive and motor outcomes in preterm born children due to EI have already been demonstrated [14]. The primary objective of this systematic review (SR) was to aggregate evidence of RCTs concerning the effect of EI on behavior, QoL, participation, EF, PCI and use of medical services in preterm born infants. To our knowledge, no previous SR has addressed these outcomes exclusively based on RCTs. Although 26 studies met inclusion criteria, only seven provided sufficiently comparable data to be included in meta-analysis. Four additional studies reported relevant but non-poolable results. The remaining studies were excluded due to non-prioritized outcomes, insufficient data, or use of ineligible assessment tools.

### Behavior

Current literature about the effects of EI on children’s behavioral development is encouraging. Although the reported effects are generally small, prior studies showed benefits especially for preterm born children [46–49]. However, robust evidence for abiding effects is limited. While there are studies reporting effects in preschool and early school age [47, 49], positive effects of EI appear to decrease when the children grow older.

Our findings supported those data, indicating modest effects on internalizing and externalizing behavior [23, 24, 31, 32], as well as a reduction of problematic behavior in both infancy [24, 42] and preschool age [43, 44]. However, the effects receded with increasing age of the participants.

This attenuation suggests that the initial benefits associated with EI diminish over time. Whether the effect could be stabilized by booster-sessions in larger intervals during childhood has not been studied yet. A declining effect of an EI in infancy is reasonable with regard to the broader developmental context in which children are growing up. According to ecological theories of child development [50], children are shaped by continuous interactions with their environment. This includes interactions with caregivers, family and peer relationships, as well as exposure to adverse childhood experiences and socioeconomic background [51]. All of these will influence the initial effect of EI during infancy.

Socioeconomic background represents an important factor influencing child development. Children exposed to socioeconomic adversity are at increased risk for developmental challenges across behavioral domains, including general behavioral difficulties [52], anxiety disorders [53], and ADHD [54, 55]. The accumulation of biological and social risk factors is thought to exacerbate developmental vulnerability [56, 57], rendering socially disadvantaged and preterm born children a key target group for EI. Even so, the current evidence of EI effects in socially high-risk groups is inconsistent. Some studies have indicated that especially children exposed to higher social risk benefit from EI [28, 58], whereas previous SRs have not identified a consistent association between socioeconomic risk and intervention effects, emphasizing the limited and heterogenous evidence base [59, 60]. Due to limited datasets, we couldn’t include socioeconomic risk as a subgroup in meta-analysis. But one study reported that preterm born children of families with higher socioeconomic risk benefit more in behavioral development and communication skills than low-risk children [43]. The inconsistent evidence regarding the effect of EI on socially disadvantaged children may be due to the diversity of study populations, as well as the varying definitions of socioeconomic risk factors. Given that both preterm birth and lower socioeconomic background are associated with an increased risk of neurodevelopmental difficulties, it is important to take both risk profiles into account when evaluating the potential benefits of EI.

Therefore, considering the impact of environmental influences, EI may be more effective when conceptualized as part of a sustained, multi-component intervention program throughout the entire childhood. Such complementary interventions may reinforce and maintain beneficial effects of EI.

### Parent-child relationship

The infant-caregiver relationship represents a critical determinant of child development [48, 51, 61–63]. Prior systematic reviews have shown that responsive PCI is associated with improved neurodevelopmental outcomes in preterm born infants, particularly in domains such as attention and social-emotional functioning [64, 65]. This emphasises the importance of fostering a supportive parent-child relationship – especially in this vulnerable population.

Preterm birth, however, may place parents at increased risk for mental health difficulties [66–68]. Mental health problems, such as post-partum depression and anxiety, can adversely affect child development [46, 51, 69] and may impair PCI [68, 70]. However, prior systematic reviews suggested that EIs involving caregivers in the therapeutic process can improve parental mental health, by reducing parenting stress and providing a social support system [46, 48]. Improvements in parental mental health, in turn, have been linked to more favorable developmental outcomes in preterm born children [51, 69, 71].

Evidence regarding the impact of EI specifically on PCI has been inconsistent. Some studies report beneficial effects [72, 73], whereas others find no significant association [74, 75]. Our results mirror this inconsistency: while some studies indicated a positive effect of EI on PCI during infancy [29, 32, 38], others demonstrated no substantial effect [27]. This inconsistency may be linked to the substantial heterogeneity across studies. Trials vary in their choice of outcome measures, timing of assessments, and intervention approaches. As a result, multiple additional moderating factors must be considered, e.g. intervention duration and intensity.

### Intervention duration and intensity

The effect of the span and intensity of service application already has been object of previous studies. Direct evidence on the impact of intervention intensity, particularly the frequency of delivered session, is scarce. However, higher intervention adherence – leading to a higher number of sessions and therefore intervention intensity – has been associated with more favorable outcomes [14, 23, 76, 77] in preterm born groups. Meanwhile the findings on a potential dose-response relationship based on intervention duration remain inconclusive in preterm born as well as non-preterm born groups. SRs have suggested a positive association in communication-focused interventions [78], but reviews examining cognitive and motor outcomes [14] and interventions for children with autism spectrum disorder [79, 80] have not demonstrated consistent effects connected to intervention duration.

Studies included in this review indicated that long-term EI lasting ≥ 6 months positively affects PCI during infancy [29, 32, 38], whereas shorter interventions of 3-4 months did not demonstrate comparable effects [27]. Given the established relevance of PCI for neurobehavioral development, this association warrants further investigation.

### Outcomes with insufficient evidence

Although preterm born infants are at increased risk of EF impairments [81–84], our findings regarding the effect of EI on EF were scarce. Similarly, the available literature did not provide sufficient evidence on the effects of EI on ADHD, ASD, utilization of medical services, quality of life, and participation. These gaps reflect both limited study numbers and substantial methodological heterogeneity.

### Strengths and limitations

Major strength of our systematic review is the adherence to established methodological standards. The research was conducted according to the Cochrane Handbook for Systematic Reviews of Interventions [19] and the PRISMA 2020 guidelines [85], based on a pre-registered protocol with transparent reporting of deviations. Screening and data extraction was performed by two independent reviewers, ensuring the transparency of the review.

Nonetheless, several limitations must be considered. Despite identifying many potentially eligible records (n = 574 full texts), only a small subset of studies could be included in data synthesis. Exclusion was primarily due to non-prioritized outcomes, missing or non-transferable data (especially in older studies), and a substantial heterogeneity in population, intervention characteristics, and assessment tools. This heterogeneity restricted the feasibility of meta-analyses and may limit the generalizability of our results. Moreover, most of the included outcomes present a low or very low certainty of evidence and a high or medium risk of bias, accompanied by relatively small effect sizes. Some meta-analyses were additionally affected by severe imbalances in study size (e.g. behavioral problems in infancy or special educational needs), potentially affecting pooled estimates. Meta-analysis for subgroups differentiating between intervention and population characteristics was not possible due to the limited dataset.

### Implications for research and practice

The limitations highlight the need for high-quality, long-term RCTs with reliable data addressing a broader range of developmental and health-related outcomes. In particular, outcomes such as quality of life and behavioral disorders (e.g. ADHD and autism) as well as health care system–related endpoints, including the utilization of medical services and special educational support, are yet to be investigated.

Future systematic reviews may benefit from focusing on a single, well-defined outcome domain (e.g. EF), allowing for more inclusive eligibility criteria and more detailed analysis of subdomains. Given the practical challenges of blinding EI research, the inclusion of high-quality quasi-RCTs or well-designed cohort studies might also strengthen the evidence base. From a clinical perspective, comparative evaluation of different intervention strategies, such as intervention duration, intensity, delivery mode (e.g. home visits and remote programs), and complementary interventions throughout childhood are particularly interesting.

## Conclusion

The available evidence, though limited and of low certainty, suggests potential benefits of EI in preterm born children for parent–child interaction and behavioral development in preschool age. Since positive effects on cognitive function up until preschool age and short-term motor function have already been shown the guideline group consented the following recommendation:

- *Early intervention programs can be recommended for preterm born (<37 week of gestation) to improve cognitive development and parent-child interaction*.
- *Early intervention programs should be recommended for any preterm infant if medical and/or social risk factors are present*.

To establish reliable clinical recommendations regarding length, duration and content high-quality long-term RCTs are urgently needed, focusing on a broader range of developmental and health-related outcomes aligned with the International Classification of Functioning, Disability, and Health.

## Supporting information

S5 - Search strategy

S1 - RoB and characteristics of included studies

S2 - Characteristics of excluded studies

S3 - Predefined and included assessment tools

S4 - Summary of Findings

## Abbreviations

ADHD: Attention Deficit/Hyperactivity Disorder
ASD: Autism Spectrum Disorder
BRIEF2: Behavior Rating Inventory of Executive Functioning, 2^nd^ edition
BW: Birth Weight
CA: Corrected Age
CBCL: Child Behavior Checklist
CoE: Certainty of Evidence
DAS: Differential Ability Scale
EAS: Emotional Availability Scale
EI: Early Intervention
ELBW: Extremely-Low-Birth-Weight (< 1000g)
EF: Executive Functioning
GA: Gestational Age
GRADE: Grading of Recommendations, Assessment, Development and Evaluations
GLOS: Greenspan-Lieberman Observation System
IRS: Interaction Rating Scale
ITSEA: Infant-Toddler Social and Emotional Assessment
LBW: Low Birth Weight
MCA: Mother-to-Child Attachment score
MD: Mean Difference
MSRS: Maternal Sensitivity and Responsivity Scale
NOSI: Nijmeegse Ouderlijke Stress Index
PBQ: Postpartum Bonding Questionnaire
PCI: Parent-Child Interaction
PSI-SF: Parenting Stress Index – short form
RCT: Randomized Controlled Trial
RR: Risk Ratio
RoB: Risk of Bias
SD: Standard Deviation
SDQ: Strengths and Difficulties Questionnaire
SR: Systematic Review
TEA-Ch: Test of Everyday Attention for children
TOL: Tower of London
SMD: Standardized Mean Difference
VLBW: Very-Low-Birth-Weight (< 1500g)
WMTB: Working Memory Test Battery, CNT arrow switching

## Data availability statement

The data that supports the findings of this systematic review are available in the supplementary material of this article.

## Declaration of generative AI and AI-assisted technologies in the manuscript preparation process

During the preparation of this work the authors used ChatGPT (OpenAI, 2025) to support language editing, including improvements in grammar and readability. After using this tool, the authors reviewed and edited the content as needed and take full responsibility for the content of the published article.

## APPENDIX A PRISMA flow diagram of screening process

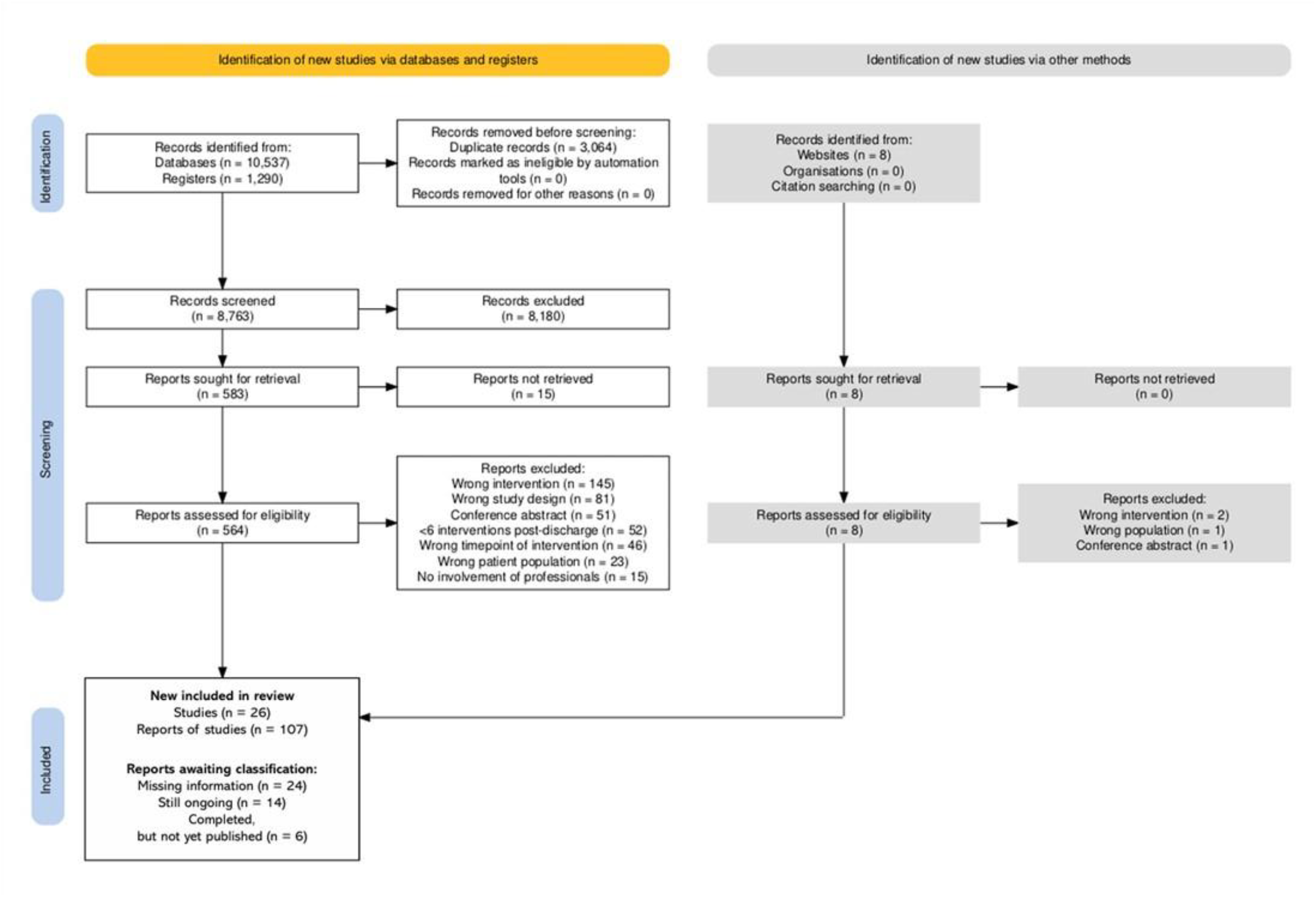

## APPENDIX B Effects of narrative outcomes (EF, PCI)

**Table Appendix B 1.** Executive functioning reported in VIBeS Plus study.

| Outcome | n | Adjusted mean group difference, (95% CI) | Interaction P |
| --- | --- | --- | --- |
| **8-year follow-up** |  |  |  |
| **Tower-of-London** |  |  |  |
| N correct in 60s | 32 | -0.5 (-1.6 to 0.6)<sup>a</sup> | 0.36 |
| N correct on 1st attempt | 31 | -0.8 (-1.7 to 0.7)<sup>a</sup> | 0.07 |
| **Working Memory Test Battery for Children** |  |  |  |
| Digit recall | 33 | 4.8 (-3.0 to 12.6) | 0.23 |
| Block recall | 33 | -1.7 (9.8 to 6.4) | 0.68 |
| Backward Digit recall | 33 | 5.4 (-2.3 to 13.2) | 0.17 |
| (Adapted from Spittle et al., Pediatrics, 2016) [37] |  |  |  |
| **13-year follow-up** |  |  |  |
| **BRIEF2** |  |  |  |
| Inhibit | 43 | -5.2 (-11.9 to 1.5) | 0.48 |
| Self-monitor | 43 | -2.5 (-9.7 to 4.8) | 0.79<sup>b</sup> |
| Shift | 43 | -4.6 (-12.4 to 3.2) | 0.34 |
| Emotional control | 43 | -6.6 (-13.8 to 0.5) | 0.70 |
| Initiate | 43 | -2.5 (-8.7 to 3.6) | 0.52 |
| Working memory | 43 | -3.2 (-9.6 to 3.2) | 0.40 |
| Planning & Organization | 43 | -3.3 (-9.7 to 3.2) | 0.20 |
| Task monitoring | 43 | -2.6 (-8.8 to 3.6) | 0.73 |
| Organization of materials | 43 | -3.0 (-8.6 to 2.5) | 0.59 |
| **Working Memory Test Battery for Children** |  |  |  |
| Digit recall | 43 | 2.9 (-6.2 to 12.0) | 0.44 |
| Block recall | 43 | 4.4 (-4.5 to 13.2) | 0.54 |
| Backward Digit recall | 43 | 2.7 (-4.3 to 9.7) | 0.38 |
| **D-KEFS Tower Test** |  |  |  |
| Planning | 43 | 0.3 (-0.8 to 1.4) | 0.63 |
| **CNT Arrow Switching** |  |  |  |
| Cognitive flexibility and inhibition | 43 | -0.2 (-0.7 to 0.4) | 0.89 |
| (Adapted from Stedall et al., J. of Pediatrics, 2022) [34] |  |  |  |

<sup>a</sup> Adjusted for age at assessment.
<sup>b</sup> Estimates from linear regression models with robust standard errors that allowed for correlation between twins rather than fitted using generalized estimating equations, due to convergence issues.

**Table Appendix B 2.**
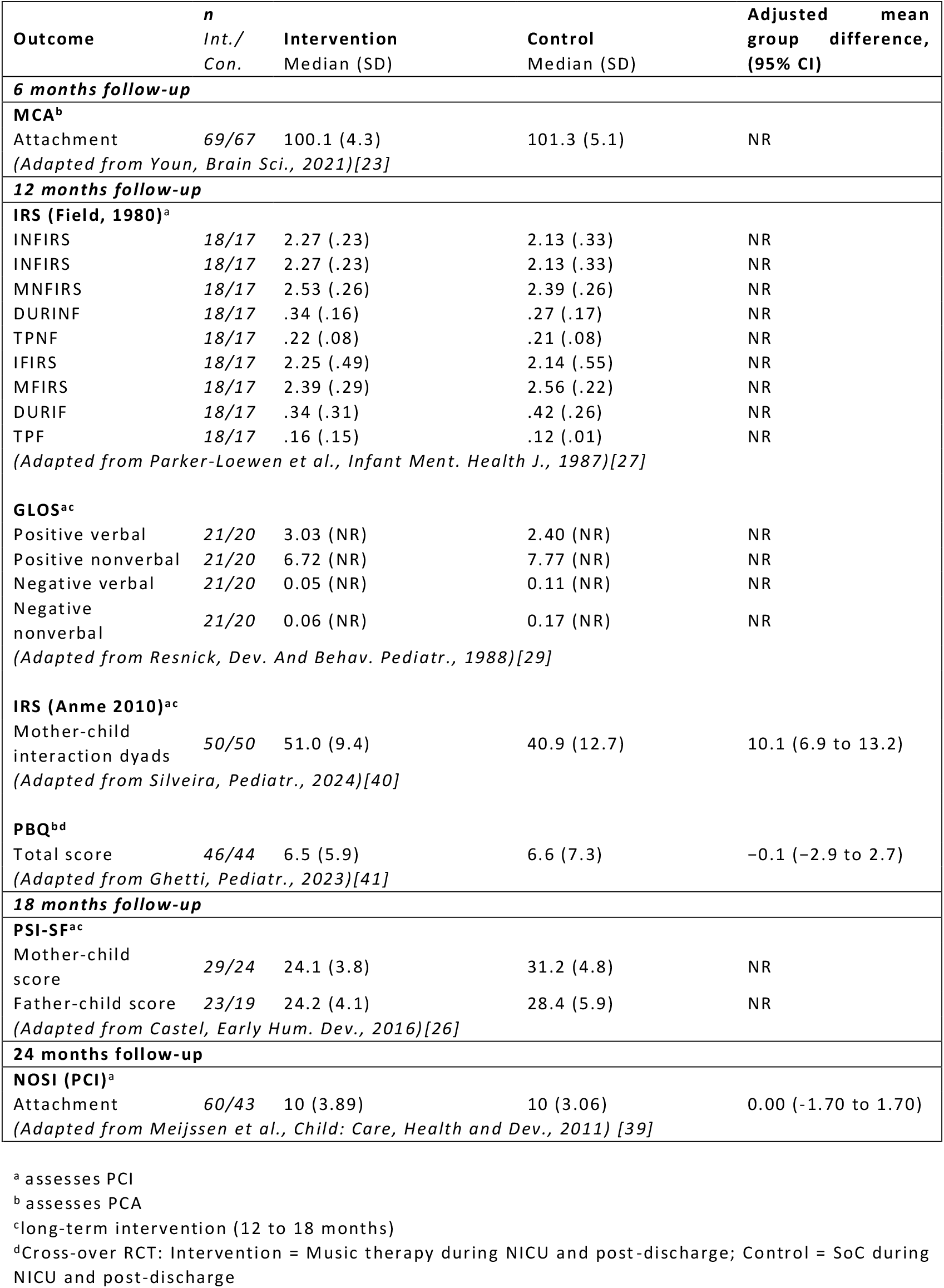
Parent-child interaction reported in different studies.

