## Supplementary material for "Effects of interdisciplinary early developmental intervention programs on behavior, executive functioning and participation in children born preterm: A systematic review with meta-analysis": S5 - Search strategy

### S2 – Characteristics of included studies

| Focus on PCI | Focus on Child Development | Focus on both |
|---|---|---|
| - Parker-Loewen<br>- Castel<br>- Wu<br>- Koldewijn | - Resnick 1988<br>- Youn<br>- IHDP<br>- APIP | - Spittle<br>- Treyvaud<br>- Silveira |

#### S2.1 – Risk of bias

| | | | |
|---|---|---|---|
| D1 | Randomisation process | | |
| D2 | Deviations from intended interventions | + | Low risk |
| D3 | Missing outcome data | ! | Some concerns |
| D4 | Measurement of outcome | - | High risk |
| D5 | Selection of the reported results | | |

#### Primary Outcomes

| Study ID | Assessment Tool | Time of Assessment | Intervention | Comparator | Outcome | D1 | D2 | D3 | D4 | D5 | Overall |
|---|---|---|---|---|---|---|---|---|---|---|---|
| **Meta-analysis** | | | | | | | | | | | |
| **Koldewijn 2009** | MSRS | 6 months | IBAIP | SoC | PCI | - | + | ! | + | ! | - |
| **Treyvaud 2022** | EAS | 24 months | e-Prem | SoC | PCI | + | + | + | + | + | + |
| Parker-Loewen 1987 | IRS (Field 1980) | 2 months | Interaction Coaching | Toys for babies | PCI | ! | + | + | + | ! | ! |
| Castel 2016 | PSI-SF | 3 months | TRT | SoC | PCI | ! | - | + | + | - | - |
| Castel 2016 | PSI-SF | 18 months | TRT | SoC | PCI | ! | - | + | + | - | - |
| Resnick 1988 | GLOS Score | 6 months | CBDI & HBDI | Soc.Serv., PT, OT | PCI | - | - | + | + | ! | - |
| Resnick 1988 | GLOS Score | 12 months | CBDI & HBDI | Soc.Serv., PT, OT | PCI | - | - | + | + | ! | - |
| Silveira 2018 | IRS (Anme 2010) | 12 months | Home-based | SoC | PCI | ! | + | + | - | + | - |
| **Meta-analysis** | | | | | | | | | | | |
| **Koldewijn 2009** | Parent Interview | 5.5 years | IBAIP | SoC | SEN | - | + | - | ! | ! | - |
| **IHDP 1990** | Parent Interview | 8 years | IHDP | SoC | SEN | ! | + | + | ! | ! | ! |
| Spittle 2010 | TOL | 8 years | VIBeS-Plus | SoC | EF | ! | + | + | ! | ! | ! |
| Spittle 2010 | CNT, TEA, D-KEFS | 13 years | VIBeS-Plus | SoC | EF | ! | + | + | ! | ! | ! |

*MSRS – Maternal Sensitivity and Responsivity Scale; EAS – Emotional Availability Scale; IRS – Interaction Rating Scale; PSI-SF – Parental Stress Index – Short Form; GLOS – Greenspan-Lieberman Observation System; TOL – Tower of London; CNT – Contingency Naming Test; TEA-Ch – Test of Everyday Attention for Children; D-KEFS – Delis–Kaplan Executive Function System; IBAIP - Infant Behavioral Assessment and Intervention Program; TRT - Triadic parent-infant Relationship Therapy; CBDI/HBDI – Clinic-based/Home-based Developmental Intervention; IHDP – Infant Health Development Program; VIBeS-Plus - Victorian Infant Brain Studies (plus); SoC – Standard of Care; PT – Physical Therapy; OT – Occupational Therapy*

### Secondary outcomes

| Study ID | Assessment Tool | Time of Assessment | Experimen-tal | Compara-tor | Outcome | D1 | D2 | D3 | D4 | D5 | Overall |
|---|---|---|---|---|---|---|---|---|---|---|---|
| Wu 2014 | CBCL 1,5-5ys | 24 months | CBIP | SoC | Behavior (int./ ext./global score) | + | + | + | ! | ! | ! |
| Wu 2014 | CBCL 1,5-5ys | 24 months | HBIP | SoC | Behavior (int./ ext./global score) | + | + | + | ! | ! | ! |
| Treyvaud 2022 | ITSEA | 24 months | e-Prem | SoC | Behavior (int./ ext. score) | + | + | + | ! | + | ! |
| Youn 2021 | m-ITSEA | 24 months | MITP | SoC | Behavior (int./ ext./global score) | + | + | - | ! | ! | - |
| Koldewijn 2009 | CBCL 1,5-5ys | 24 months | IBAIP | SoC | Behavior (int./ ext./global score) | - | + | - | - | + | - |
| Koldewijn 2009 | CBCL 1,5-5ys | 44 months | IBAIP | SoC | Behavior (int./ ext./global score) | - | + | + | ! | ! | - |
| Koldewijn 2009 | SDQ | 5.5 years | IBAIP | SoC | Behavioral Problem Score | - | + | + | ! | ! | - |
| Koldewijn 2009 | Parent interview | 5.5 years | IBAIP | SoC | Need f. psycholog. support | - | + | - | + | ! | - |
| Koldewijn 2009 | Parent interview | 5.5 years | IBAIP | SoC | Need f. paramedical support | - | + | - | + | ! | - |
| Spittle 2010 | ITSEA, higher social risk | 24 months | VIBeS-Plus | SoC | Behavior (int./ ext.score) | ! | + | + | ! | + | ! |
| Spittle 2010 | ITSEA, lower social risk | 24 months | VIBeS-Plus | SoC | Behavior (int./ ext.score) | ! | + | + | ! | + | ! |
| Spittle 2010 | CBCL; high social risk | 4 years | VIBeS-Plus | SoC | Behavioral Problem Score | ! | + | + | ! | ! | ! |
| Spittle 2010 | CBCL; lower social risk | 4 years | VIBeS-Plus | SoC | Behavioral Problem Score Behavior | ! | + | + | ! | ! | ! |
| Spittle 2010 | SDQ+SSISR | 8 years | VIBeS-Plus | SoC | Total problem score | ! | + | + | ! | ! | ! |
| Spittle 2010 | SDQ | 13 years | VIBeS-Plus | SoC | Total problem score | ! | + | + | ! | ! | ! |
| Castel 2016 | Behavioral SCL | 9 months | TRT | SoC | Behaviour - Total Score | ! | - | + | + | + | - |
| Castel 2016 | Behavioral SCL | 18 months | TRT | SoC | Behaviour - Total Score | ! | - | + | + | + | - |
| IHDP 1990 | CBCL | 3 years | IHDP | SoC | Behavioral Problem Score | ! | + | + | ! | ! | ! |
| APIP 1998 | CBCL | 5 years | Portage | another intervention | Behavioral Problem Score | + | + | - | ! | ! | - |

*CBCL – Child Behavior Checklist; (m-)ITSEA – (modified) Infant and Toddler Social-Emotional Assessment; SDQ – Strength and Difficulties Questionnaire; SSISR - Social Skills Improvement System Rating Scales; SCL – Symptom Checklist; CBIP/HBIP – Clinic-Based/Home-Based Intervention Program; ; IBAIP - Infant Behavioral Assessment and Intervention Program; VIBeS-Plus - Victorian Infant Brain Studies (plus); TRT - Triadic parent-infant Relationship Therapy; IHDP – Infant Health Development Program; SoC – standard of care; int. – internailzing; ext. - externalizing*

#### S2.2 – Studies included in meta-analysis

| | |
|---|---|
| **Study ID** | IHDP 1990 |
| **DOI** | 10.1001/jama.1990.03440220059030 |
| **Title** | **The effectiveness of early intervention: examining risk factors and pathways to enhanced development** |
| **Type of RCT** (parallel/multiple groups) | Parallel group study |
| **Additional study reports available** | Secondary journal pulications (e.g. follow-up study) |
| **Study objective:**<br>To examine … | Child<br>- Behavior<br>- Participation<br>- Cognition<br>- Motor development<br>- Health and need of (para-)medical support<br>Caregiver:<br>- Psychological wellbeing<br>- Behavior<br>- Knowledge about child development<br>Caregiver and Child:<br>- Parent-child interaction |
| **Setting** (single-/multi-centric) | multi-centric |
| **Country in which the study conducted** | United States |
| **Start date** | 07/01/1985 |
| **End date** | 09/10/1985 |
| **Funding source** | grants from:<br>- Robert Wood Johnson Foundation to the Department of Pediatrics, Stanford University, Stanford, Calif<br>- Frank Porter Graham Child Development Center, University of North Carolina at Chapel Hill<br>- the eight participating universities.<br>Additional support:<br>- National Study Office to the Department of Pediatrics, Stanford University, from the Pew Charitable Trusts<br>- Bureau of Maternal and Child Health and Resources Development<br>- National Institute of Child Health and Human Development, Health Resources and Services Administration, Public Health Service, US Dept of Health and Human Services (grant MCJ-060515) |

| | | | | |
|---|---|---|---|---|
| | - Stanford Center for the Study of Families, Children, and Youth. | | | |
| **Trial registration number(s)** | NR | | | |
| **Conflict of interest** | NR | | | |
| **Inclusion criteria** | - GA ≤37 weeks<br>- BW ≤2500 g<br>- born 07/01/1985 - 09/10/1985<br>- Surviving neonatal hospitalization<br>- Unhealthy infants were included if participation in the intervention program was possible<br>- Living within 45 min of one of eight participating centers | | | |
| **Exclusion criteria** | - Residency too far away<br>- GA > 37 weeks<br>- Hospital discharge before or after the designated recruitment period | | | |
| **preterms as...** | Full population | | | |
| **gestational age (weeks)** | preterm (<37 weeks) | | | |
| **birth weight** | LBW (< 2500g) | | | |
| **If multiple group study, define intervention 1, intervention 2, intervention 3** | **IG 1** | Intervention | | |
| | **IG 2** | control | | |
| | **IG 3** | N/A | | |
| **Sociodemographic Characteristics:** | **IG 1** | **IG 2** | **IG 3** | **total** |
| **sociodemographic characterstics of parents** — **education** | NR | NR | N/A | *Mothers*<br>< high school degree: 40%<br>High school diploma: 27%<br>Some college: 20%<br>College degree: 13% |
| **income** | NR | NR | N/A | NR |
| **employment** | NR | NR | N/A | NR |
| **marital status** | NR | NR | N/A | NR |
| **Number of participants allocated** | 377<br><br>Heavier LBW: 142<br>Lighter LBW: 235 | 608<br><br>Heavier LBW: 220<br>Lighter LBW: 388 | N/A | 985<br><br>Heavier LBW: 362<br>Lighter LBW: 623 |
| **baseline population characterstics** | **IG 1** | **IG 2** | **IG 3** | **total** |
| **No. of participants** | 377 | 608 | N/A | 985 |

| | | | | |
|---|---|---|---|---|
| **Mean birthweight (SD) in gramme** | NR | NR | N/A | 1800 (SD not reported) |
| **Mean gestational age (SD) in weeks** | NR | NR | N/A | 33 (SD not reported) |
| **No. of male/female** | NR | NR | N/A | NR |
| **percentage of male/female (in %)** | NR | NR | N/A | NR |
| **multiples** | NR | NR | N/A | NR |
| **Rate of BPD** | NR | NR | N/A | NR |
| **Rate of IVH** | NR | NR | N/A | NR |
| **Rate of PVL/white matter injury** | NR | NR | N/A | NR |
| **Rate of Cerebral Palsy** | NR | NR | N/A | NR |
| **Intervention** | | | | |
| **Specific interventional concepts** | Infant Health Development Program (IHDP) | | | |
| **Description of intervention** | Home visits:<br>- Family support<br>- Education about infant health and development<br>- Curriculum with play activities to promote cognitive, linguistic, and social development<br>- Curriculum to improve parental self-efficiacy and competence<br>Child development centre (CDC):<br>- Individualized curriculum of learning activities<br>- Teacher-child ratio 1:3 (1st year in CDC) or 1:4 (2nd year in CDC)<br>Parent Groups:<br>- Peer support regarding child rearing, health, safety, and other concerns | | | |
| **Aim of intervention:**<br>Improvement of**…** | - | | | |
| **Age of participants at start of intervention (years)** | Post-discharge | | | |
| **Setting** | Home visits<br>Child development centres<br>Parent groups | | | |
| **Number of sessions** | home visits: 104x<br>child development centers: 20h per week for 24 months<br>parent groups: 18x | | | |
| **Frequency of sessions** | home visits:<br>weekly druing the first year, bi-weekly the rest of the intervention<br><br>child developmet centers:<br>20h per week from 12 months on<br><br>parent groups:<br>every other month | | | |
| **Duration of program** | 3 years | | | |
| **Comparator** | | | | |
| **Type of comparator** | SoC | | | |
| **Outcomes** | **Assessment tool** | | | |

| | | | |
|---|---|---|---|
| **Outcomes relevant for our study** | | | |
| **Behavior** | **2-3 years** | Child Behavior Checklist | |
| **Special educational needs** | **5-8 years** | Caregiver and teacher interviews | |
| **Relevant but not eligible outcomes for meta-analysis** | | | Comment |
| **Behaviour (internalizing, externailzing)** | **36 months**<br>**5 years** | Age-appropriate Child Behavior Checklist | Missing data; |
| **Behaviour (internalizing, externailzing)** | **8 years** | Age-appropriate Child Behavior Checklist | no differentiation of intervention and control |
| **Behavioral (problems)** | **18 years** | Behavioral Problem Index | Missing data |
| **Participation (language)** | **5 years**<br>**8 years** | Peabody Picture Vocabulary Test | Too early |
| **Parent-Child Interaction (maternal sensitivity)** | **30 months** | Rake Box Task | Not enough information about measurement method |
| **Maternal supportive behavior towards child** | **30 months** | Rake Box Task | Not poolable |
| **Outcomes *not* relevant for our study** | | | Comment |
| *child* | | | |
| **Growth** | **40 weeks p.c.;**<br>**4/8/12/18/24/30/36 months**<br>**8 years** | Measurements (weight, height, head circumference, BMI) | |
| **Cognition** | **12/24/36 months** | Bayley Scale | |
| **Cognition** | **24 months**<br>**36 months** | Stanford–Binet Intelligence Scale | |
| **Cognition** | **18/36 months**<br>**4/5/8/18 years** | Peabody Picture Vocabulary Test, revised | |
| **Cognition** | **5/8 years** | IQ-Score | |
| **Cogintion** | **5 years** | Wechsler Preschool and Primary Scale of Intelligence | |
| **Cognition** | **8 years** | Wechsler Intelligence Scale for Children | |
| **Cognition** | **8/18 years** | Woodcock-Johnson Tests of Achievement–Revise (Reading, Math) | |
| **Cognition** | **18 years** | Weschler Abbreviated Scale of Intelligence | |
| **Executive Functioning (problem solving)** | **30 months** | Rake Box Task | Too early |

| | | | |
|---|---|---|---|
| **Child temperament** | **12 months** | Infant Characteristics Questionnaire | |
| **Adaptive Social Behavior** | **36 months** | Adaptive Social Behavior Inventory, expressive scale | |
| **Socioemotional behavior** | **24/36 months** | Richman-Graham Behavior Checklist (BCL) | |
| **Socioemotional behavior** | **36 months** | Child Behavior Checklist 2-3 years | |
| **Socioemotional behavior** | **36 months** | Richman-Graham Behavior Checklist (BCL), Child Behavior Checklist (CBCL) | |
| **Risky behavior** | **18 years** | Youth Risk Behavior Surveillance System | Missing data |
| **Youths' justice system involvement** | **18 years** | Self-report, caregiver report | |
| **Motor function** | **12/24/36 months** | Bayley Scale | |
| **Morbidity index** | **36 months** | Maternal report (number of hospitalizations, out-patient surgeries, injuries not resulting in hospitalization or surgery, and different illnesses and conditions) | |
| **Physical health** | **18 years** | self-report and caregiver report | |
| **Annual rate of conditions/hospitali-zations** | **0-5th year** | Maternal report | |
| **Home environment** | **12/36 months<br>18 years** | Age-appropriate version of HOME Inventory | |
| **Home environment** | **5 years** | Adult-child activities scale; home literacy | |
| **Home environment** | **8 years** | Rochester Assessment Package for Schools (RAPS)Parent Report;<br>Conflict subscale of the Family Environment Scale | |
| *caregiver* | | | |
| **Maternal emotional distress** | **40 weeks GA** | General Health Questionnaire | |
| **Maternal depression** | **12 months** | | |
| **Maternal health** | **5/8/18 years** | Medical Outcomes Study General Health Questionnaire – Short-Form | |
| **Parental understanding of child development** | **12/36 months** | Concepts of Development Questionnaire | |
| **Parenting characteristics** | **30 months** | Videotaped mother and child free-play activity; Mother–child problem-solving assessment | |

| | |
|---|---|
| **Study ID** | APIP 1998 |

| | |
|---|---|
| **DOI** | 10.1136/adc.2004.057620 |
| **Title** | **Randomised trial of parental support for families with very preterm children. Avon Premature Infant Project** |
| **Type of RCT** (parallel/multiple groups) | Multiple group study |
| **Additional study reports available** | Secondary journal pulication (e.g. follow-up study) |
| **Study objective:**<br>To examine ... | - neurological development at 2 years CA<br>- cognitive, motor and behavioural development at 5 years CA. |
| **Setting** (single-/multi-centric) | multi-centric |
| **Country in which the study conducted** | United Kingdom |
| **Start date** | xx/12/1990 |
| **End date** | xx/07/1993 |
| **Funding source** | Primary study:<br>five year project grant from Action Research;<br>supplementary funding by Crookes Health Care, Nutricia, and Milupa (UK) Ltd.<br><br>Follow-up study:<br>Action Medical Research Grant to Professor Neil Marlow |
| **Trial registration number(s)** | NR |
| **Conflict of interest** | Primary publication: NR<br>Follow-up: None |
| **Inclusion criteria** | - GA ≤32+6 weeks<br>- Mothers resident in the greater Bristol area<br>- English as the first language used at home |
| **Exclusion criteria** | - English not first language at home<br>- Infants missed due to early discharge policies |
| **preterms as...** | Full population |
| **gestational age (weeks)** | <33 |
| **birth weight** | LBW (<2500g) |
| **If multiple group study, define intervention 1, intervention 2, intervention 3** | **IG 1** Portage |
| | **IG 2** Parent Adviser |
| | **IG 3** Control |

| **Sociodemographic Characteristics:** | | **IG 1** | **IG 2** | **IG 3** | **total** |
|---|---|---|---|---|---|
| **sociodemographic characterstics of parents** | **education** | beyond 16 years, n(%)<br>45 (50) | beyond 16 years, n(%)<br>(47 (53,4) | 55 (66.3) | NR |
| | **income** | NR | NR | NR | NR |
| | **employment** | NR | NR | NR | NR |

| | | | | |
|---|---|---|---|---|
| **marital status** | Lives with both parents n(%) 84 (91.3) | Lives with both parents n(%) 72 (81.8) | Lives with both parents n(%) 72 (88.9) | NR |
| **Number of participants allocated** | 116 | 106 | 106 | 328 |
| **baseline population characterstics** | **IG 1** | **IG 2** | **IG 3** | **total** |
| **No. of participants** | 111 | 99 | 99 | 309 |
| **Median birthweight (IQR) in gramme** | 1560 (1280-1735) | 1331 (1078-1700) | 1420 (1095-1700) | NR |
| **Median gestational age (IQR) in weeks** | 31 (29-32) | 30 (29-31) | 31 (28-32) | NR |
| **No. of male/female** | 68/43 | 50/49 | 59/40 | 177/132 |
| **percentage of male/female (in %)** | 62/38 | 50/50 | 59/41 | 57/43 |
| **multiples** | 24 children | 20 children | 14 children | 58 children |
| **Rate of BPD** | NR | NR | NR | NR |
| **Rate of IVH** | NR | NR | NR | NR |
| **Rate of PVL/white matter injury** | 9 | 6 | 8 | 23 |
| **Rate of Cerebral Palsy** | 12 (9 with severe disability) | 7 (6 with severe disability) | 11 (10 with severe disability) | 30 (diagnosed at 2 years of age) |
| **Intervention** | | | | |
| **Specific interventional concepts** | Portage; Parent Adivser | | | |
| **Description of intervention** | Portage:<br>- Family-centred<br>- Parent training in supporting child's development<br>- Support in accessing social community services → parent adviser scheme<br><br>Parent Adviser:<br>- Supportive counselling model<br>- Seminars, individual and group sessions | | | |
| **Aim of intervention:** Improvement of**...** | Portage: ...children's developmental progress<br><br>Parent Adviser:... access to social community services | | | |
| **Age of participants at start of intervention** | median 11 (10 to 13) days post term | | | |
| **Setting** | Home-visits | | | |
| **Number of sessions** | Portage<br>- Discharge to 6 months post term: 17<br>- 6–12 months post term: 11<br>- 12–18 months post term: 8<br>- 18–24 months post term: 6<br>- Length of visit (minutes): 44<br><br>Parent Adviser:<br>- Discharge to 6 months post term: 16 | | | |

| | |
|---|---|
| | - 6–12 months post term: 12<br>- 12–18 months post term: 8<br>- 18–24 months post term: 6<br>- Length of visit (minutes): 45 minutes |
| **Frequency of sessions** | frequency of visiting was tailored to suit the family:<br><br>first months weekly<br>following year 2-4 weekly<br>monthly by the time disengagement occurred at 2 years |
| **Duration of program** | 2 years |
| **Comparator** | |
| **Type of comparator** | Another type or mode of EI |
| **Outcomes** | **Assessment tool** |
| **Outcomes relevant for our study** | |
| **Behavior 5 years** | Child Behavior Checklist (4-18 years) |
| **Outcomes *not* relevant for our study** | |
| **Neurological outcomes 2 years** | Diagnosis of different grades of neurological disability |
| **Overall mental development 2 years** | Griffiths Mental Developmental Scale |
| **Cognition 5 years** | British Ability Scale – 2nd edition |
| **Motor 5 years** | Movement ABC |

| | |
|---|---|
| **Study ID** | Koldewijn 2009 |
| **DOI** | 10.1016/j.jpeds.2008.07.039 |
| **Title** | **The Infant Behavioral Assessment and Intervention Program for very low birth weight infants at 6 months corrected age** |
| **Type of RCT (parallel/multiple groups)** | Parallel group study |
| **Additional study reports available** | Secondary journal pulication (e.g. follow-up study);<br>Trial registry record without results |
| **Study objective:**<br>**To examine ...** | - developmental and neurobehavioral outcomes at 6 and 24 months CA<br>- ability of self-regulation<br>- sensory processing and daily activities at preschool age<br>- executive functioning, behaviour and cognition at preschool age<br>- cognitive, neuromotor and behavioral development at 5.5 years CA<br>- cognitive and motor development of very preterm infants from 6 months to 5.5 years CA<br>- on mother–infant interaction at 6 months corrected age<br>- maternal psychological distress at 6, 12 and 24 (corrected) months<br>- the maternal attachment representations at 18 months CA<br>- maternal parenting stress as a secondary outcome at 12 and 24 months post term |
| **Setting (single-/multi-centric)** | multi-centric |
| **Country in which the study conducted** | Netherlands |
| **Start date** | xx/01/2004 |
| **End date** | xx/04/2006 |
| **Funding source** | Innovatiefonds Zorgverzekeraars (= Krankenkassen)<br>(project 576, supporting the implementation of the inter-<br>vention program);<br>Zorg Onderzoek (= Forschung im Gesundheitswesen) Nederland<br>(project 62200032, supporting<br>the first author, who wrote the first draft<br>of the manuscript) |
| **Trial registration number(s)** | ISRCTN65503576 |
| **Conflict of interest** | None |
| **Inclusion criteria** | - GA <32 weeks and/or BW <1500 g<br>- Parents lived in Amsterdam |
| **Exclusion criteria** | - Severe congenital abnormalities<br>- Mothers with a documented history of illicit drug use or severe physical or mental illness<br>- Non-Dutch speaking families for whom an interpreter could not be arranged<br>- Participation in another trial on postdischarge management |
| **preterms as...** | Full population |
| **gestational age (weeks)** | <32 weeks |
| **birth weight** | VLBW (< 1500g) |

| If multiple group study, define intervention 1, intervention 2, intervention 3 | | |
|---|---|---|
| | **IG 1** | Intervention |
| | **IG 2** | control |
| | **IG 3** | N/A |

| **Sociodemographic Characteristics:** | | **IG 1** | **IG 2** | **IG 3** | **total** |
|---|---|---|---|---|---|
| **sociodemographic characterstics of parents** | **education** | *Maternal, n (%)*<br>Not high school graduate<br>30 (35)<br><br>High school graduate<br>56 (65)<br><br>*Paternal, n (%)*<br>Not high school graduate<br>33 (40)<br><br>High school graduate<br>49 (60) | *Maternal, n (%)*<br>Not high school graduate<br>36 (41)<br><br>High school graduate<br>52 (59)<br><br>*Paternal, n (%)*<br>Not high school graduate<br>34 (39)<br><br>High school graduate<br>53 (61) | N/A | NR |
| | **income** | NR | NR | N/A | NR |
| | **employment** | Mother with job, n (%)<br>63 (73)<br><br>Father with job, n (%)<br>70 (85) | Mother with job, n (%)<br>53 (61)<br><br>Father with job, n (%)<br>69 (80) | N/A | NR |
| | **marital status** | 2 parents, n (%)<br>70 (81) | 2 parents, n (%)<br>82 (91) | N/A | NR |
| **Number of participants allocated** | | 86 | 90 | N/A | 176 |
| **baseline population characterstics** | | **IG 1** | **IG 2** | **IG 3** | **total** |
| **No. of participants** | | 86 | 90 | N/A | 176 |
| **Mean birthweight (SD) in gramme** | | 1242 (332) | 1306 (318) | N/A | NR |
| **Mean gestational age (SD) in weeks** | | 29.6 (2.2) | 30.0 (2.2) | N/A | NR |
| **No. of male/female** | | 50/36 | 41/49 | N/A | 91/85 |
| **percentage of male/female (in %)** | | 58/36 | 41/49 | N/A | 91/85 |
| **Multiples, n(%)** | | 27 (31) | 26 (29) | N/A | 53 (30) |
| **Rate of BPD, n(%)** | | 34 (40) | 18 (20) | N/A | NR |
| **Rate of IVH** | | Grad 1+2: 15<br>Grad 3: 6<br>17% | Grad 1+2: 9<br>Grad 3: 5<br>16% | N/A | Grad 1+2: 24<br>Grad 3: 11 |
| **Rate of PVL/white matter injury** | | Grad 1: 11<br>Grad 2+3: 1<br>12% | Grad 1: 8<br>Grad 2+3: 2<br>11% | N/A | Grad 1: 19<br>Grad 2+3: 3 |

| | | | | | |
|---|---|---|---|---|---|
| **Rate of Cerebral Palsy** | | NR | NR | N/A | NR |
| **Intervention** | | | | | |
| **Specific interventional concepts** | | IBAIP (Infant Behavioral Assessment and Intervention Program) | | | |
| **Description of intervention** | | - based on "Synactive Model of Newborn Behavioral Organization and Development"<br>- emotional, practical and individual support by physical therapist<br>- improve ability to read and interpret the infant's behavioral cues and co-regulate the environment<br>- consideration of family composition and cultural background | | | |
| **Aim of intervention:** Improvement of... | | - infants' social and environmental interaction<br>- the infant's motivation and autonomy<br>- builds on the strengths of infants and caregivers | | | |
| **Age of participants at start of intervention (years)** | | 0-1y (shortly before discharge) | | | |
| **Setting** | | Started inpatient in NICU;<br>Home-visits | | | |
| **Number of sessions** | | 1 in NICU,<br>6 to 8 home interventions | | | |
| **Frequency of sessions** | | NR | | | |
| **Duration of program** | | Up to 6 months corrected age | | | |
| **Comparator** | | | | | |
| **Type of comparator** | | SoC | | | |
| **Outcomes** | | **Assessment tool** | | | |
| **Outcomes relevant for our study** | | | | | |
| **Behavior** | **6 months** | Infant Behavioral Assessment | | | |
| **Behavior** | **6/24 months** | Behavior Rating Scale | | | |
| **Behavior** | **24/44 months** | Child Behavior Checklist | | | |
| **Behavior** | **5.5 years** | Strength and Difficulties Questionnaire | | | |
| **Use of paramedical support** | **24 months** | Use of pediatric physical therapy and/or occupational therapy and/or speech therapy | | | |
| **Parent-Child Interaction (maternal sensitivity)** | **6 months** | Maternal Sensitivity and Responsivity Scale | | | |
| **Relevant, but *not eligible* outcomes for meta-analysis** | | | | | |
| **Parental attachment** | **18 months** | Working Model of the Child Interview | | | |
| **Parental attachment** | **12 months**<br><br>**24 months** | Nijmeegse Ouderlijke Stress Index – short version<br><br>Nijmeegse Ouderlijke Stress Index | | | |
| **Parent-Child Interaction** | **6 months** | ICEP coding system | | | not poolable |
| **Executive functioning** | **5.5 years** | Visual Motor Integration | | | no overall EF score |
| **Outcomes *not* relevant for our study** | | | | | |

| | | | |
|---|---|---|---|
| **Behavioral inhibition** | **44 months** | Gift Delay Task | |
| **Cognition** | **6/12/24 months** | Bayley Scale 2 (Mental Developmental Index) | |
| **Cognition** | **44 months** | Peabody Picture Vocabulary Test III-NL<br>Miller Assessment for Preschoolers | |
| **Cognition** | **5.5 years** | Wechsler Preschool and Primary Scale of Intelligence, third Dutch version | |
| **Motor** | **6/12/24 months** | Bayley Scale 2 (Psychomotor Developmental Index) | |
| **Motor** | **6/12/24 months**<br>**5.5 years** | Movement Assessment Battery for Children – 2nd edition | |
| **Minor neurologic disfunctions** | **6 months**<br>**5.5 years** | Touwen neurologic examination | |
| **Neurologic** | **24 months** | Not further defined | |
| **Sensory processing** | **44 months** | Sensory Profile – Dutch version | |
| **Participation (daily activities)** | **44 months** | Pediatric Evaluation of Disability Inventory – Dutch version | too early |
| **Executive functioning** | **44 months** | Behavior Rating Inventory of Executive Function-Preschool<br>Visual Motor Integration<br>Visual Attention Task | too early |
| **Maternal psychological distress** | **6/12/24 months** | General Health Questionnaire | |

| | | | | |
|---|---|---|---|---|
| **Study ID** | Treyvaud 2022 | | | |
| **DOI** | 10.1542/peds.2021-055398 | | | |
| **Title** | **Preterm Infant Outcomes at 24 Months After Clinician-Supported Web-Based Intervention** | | | |
| **Type of RCT** (parallel/multiple groups) | Parallel group study | | | |
| **Additional study reports available** | Trial registry record without results | | | |
| **Study objective:**<br>To examine … | - the efficacy over the first year after birth<br>- the child development at 24 months CA.<br>- parental mental health, and the parent-child relationship at 24 months. | | | |
| **Setting** (single-/multi-centric) | single-centric | | | |
| **Country in which the study conducted** | Australia | | | |
| **Start date** | xx/01/2015 | | | |
| **End date** | xx/12/2016 | | | |
| **Funding source** | National Health and Medical Research Council<br>(Centre for Research Excellence in Newborn Medicine 1060733 and 1153176;<br>project grants 1024516, 1028822;<br>Career Development Fellowship 1108714 to Dr Spittle and 1127984 to Dr Lee;<br>Senior Research Fellowship 1081288 to Dr Anderson;<br>Investigator Grant 1176077 to Dr Anderson);<br>Murdoch Children's Research Institute is supported by the Victorian Government's Operational Infrastructure Support Program. | | | |
| **Trial registration number(s)** | ACTRN12614000906651 | | | |
| **Conflict of interest** | None | | | |
| **Inclusion criteria** | - GA <34 weeks<br><br>Admission to NICU at the Royal Women's Hospital in Melbourne, Victoria, between January 2015 to December 2016 | | | |
| **Exclusion criteria** | - Known congenital or genetic abnormality known to adversely affect development<br><br>Non-English speaking parents | | | |
| **preterms as…** | full population | | | |
| **gestational age (weeks)** | <34 | | | |
| **birth weight** | NR | | | |
| **If multiple group study, define intervention 1, intervention 2, intervention 3** | **IG 1** | Intervention | | |
| | **IG 2** | control | | |
| | **IG 3** | N/A | | |
| **Sociodemographic Characteristics:** | **IG 1** | **IG 2** | **IG 3** | **total** |
| **education**<br><br>**sociodemographic characterstics of parents** | **tertiary**<br>33<br>**secondary**<br>8<br>**<12 years**<br>0 | **tertiary**<br>29<br>**secondary**<br>11<br>**<12 years**<br>2 | N/A | NR |

| | | | | |
|---|---|---|---|---|
| **income**<br>**employment** | NR<br>full time<br>36<br>part time<br>3<br>unemployed<br>2 | NR<br>full time<br>37<br>part time<br>2<br>unemployed<br>3 | N/A<br>N/A | NR<br>full time<br>73<br>part time<br>5<br>unemployed<br>5 |
| **Marital/family status** | Family structure<br><br>Two parents<br>39<br>One parent<br>2 | Family structure<br><br>Two parents<br>42<br>One parent<br>0 | N/A | Family structure<br><br>Two parents<br>81<br>One parent<br>2 |
| **Number of participants allocated** | 50 | 53 | N/A | 103 |
| **baseline population characterstics** | **IG 1** | **IG 2** | **IG 3** | **total** |
| **No. of participants** | 50 | 53 | N/A | 103 |
| **Mean birthweight (SD) in gramme** | 1317 (322) | 1269 (447) | N/A | NR |
| **Mean gestational age (SD) in weeks** | 29.2 (2.1) | 28.7 (1.4) | N/A | NR |
| **No. of male/female** | 31/19 | 20/33 | N/A | 51/52 |
| **percentage of male/female (in %)** | 62/38 | 38/62 | N/A | NR |
| **multiples** | 19 | 21 | N/A | 40 |
| **Rate of BPD** | 8/48 | 15/49 | N/A | 23/97 |
| **Rate of IVH** | 9/44 | 12/51 | N/A | 21/95 |
| **Rate of PVL/white matter injury** | NR | NR | N/A | NR |
| **Rate of Cerebral Palsy** | NR | NR | N/A | NR |
| **Intervention** | | | | |
| **Specific interventional concepts** | - e-Prem | | | |
| **Description of intervention** | - 8 age-dependent modules on the e-Prem-Website with 5-10 topics about infant development, parent-infant interaction, and mental health of caregivers<br>- Phone-based clinician support up to 12 months CA | | | |
| **Aim of intervention:**<br>Improvement of… | - infant development<br>- parent-infant relationship<br>- parental mental health | | | |
| **Age of participants at start of intervention (years)** | 0-1 | | | |
| **Setting** | Web- and phone-based intervention at home | | | |
| **Number of sessions** | As needed | | | |
| **Frequency of sessions** | As needed (8 modules; 11 phone calls in average) | | | |
| **Duration of program** | 12 months | | | |
| **Comparator** | | | | |
| **Type of comparator** | SoC | | | |
| **Outcomes** | **Assessment tool** | | | |
| **Outcomes relevant for our study** | | | | |
| **Language** **24 months** | Bayley-III | | | |
| **Behavior** **24 months** | ITSEA | | | |
| **Parent-child interaction** **24 months** | Emotional Availability Scale (EAS) | | | |
| **Outcomes *not* relevant for our study** | | | | |

| | | |
|---|---|---|
| **Cognition** | **24 months** | Bayley-III |
| **Motor** | **24 months** | Bayley-III |
| **Parental wellbeing** | **24 months** | Centre for Epidemiologic Studies Depression Scale (CES-D);<br>Generalized Anxiety Disorder-Questionnaire;<br>Posttraumatic checklist – Diagnostic and Statistical Manual of Mental Disorders, Fifth Edition;<br>Assessment of Quality of Life |

| Study ID | Spittle 2010 |
|---|---|
| **DOI** | 10.1542/peds.2009-3137 |
| **Title** | **Preventive Care at Home for Very Preterm InfantsImproves Infant and Caregiver Outcomes at 2 Years** |
| **Type of RCT** (parallel/multi-groups) | Parallel group study |
| **Additional study reports available** | - Secondary journal pulication (e.g. follow-up study)<br>- Study protocol<br>- Trial registry record without results |
| **Study objective:**<br>To examine … | - the effects of preventive care at home on child development.<br>- primary caregiver mental health at 24 months. |
| **Setting** (single-/multi-centric) | single-centric |
| **Country in which the study conducted** | Australia |
| **Start date** | xx/01/2005 |
| **End date** | xx/01/2007 |
| **Funding source** | National Health and Medical Council (project grant ID 284512 and career development award ID 473840, to Dr Boyd);<br>Cerebral Palsy Foundation (project grant and postdoctoral fellow-ship, to Dr Spittle);<br>Murdoch Childrens Research Institute;<br>Myer Foundation;<br>Allens Arthur Robinson;<br>Thyne Reid Foundation |
| **Trial registration number(s)** | ACTRN12605000492651<br>ACTRN12606000252516 |
| **Conflict of interest** | None |
| **Inclusion criteria** | - GA < 30 weeks<br>- No major congenital brain anomalies associated with poor neurodevelopmental outcome |
| **Exclusion criteria** | - Not living within a 100-km radius of the hospital<br>- Non-English speaking parents |
| **preterms as...** | Full population |
| **gestational age (weeks)** | < 30 |
| **birth weight** | NR |

| **If multiple group study, define intervention 1, intervention 2, intervention 3** | **IG 1** | Intervention |
|---|---|---|
| | **IG 2** | control |
| | **IG 3** | N/A |

| **Sociodemographic Characteristics:** | | **IG 1** | **IG 2** | **IG 3** | **total** |
|---|---|---|---|---|---|
| **sociodemographic characterstics of parents** | **education** | beyond secondary, n (%)<br>21 (40) | beyond secondary, n (%) | N/A | NR |

| | | | | |
|---|---|---|---|---|
| | tertiary educated, n (%)<br>19 (44) | 20 (43)<br><br>tertiary educated, n (%)<br>22 (67) | | |
| **income**<br>**employment** | NR<br>skilled profession, n (%)<br>28 (67)<br><br>full-time employed, n (%)<br>42 (98) | NR<br>skilled profession, n (%)<br>28 (67)<br><br>full-time employed, n (%)<br>42 (98) | N/A<br>N/A | NR<br>NR |
| **marital status** | Two-caregiver family, n (%)<br>42 (98) | Two-caregiver family, n (%)<br>37 (97) | N/A | NR |
| **Number of participants allocated** | 61 | 59 | N/A | 120 |
| **baseline population characterstics** | **IG 1** | **IG 2** | **IG 3** | **total** |
| **No. of participants** | 61 | 59 | N/A | 120 |
| **Mean birthweight (SD) in gramme** | 1029 (287) | 991 (244) | N/A | NR |
| **Mean gestational age (SD) in weeks** | 27.3 (1.6) | 27.4 (1.4) | N/A | NR |
| **No. of male/female** | 34/27 | 27/32 | N/A | 61/59 |
| **percentage of male/female (in %)** | 56/44 | 46/54 | N/A | 51/49 |
| **multiples** | 20 | 19 | N/A | 39 |
| **Rate of BPD** | NR | NR | N/A | NR |
| **Rate of IVH** | 3 | 3 | N/A | 6 |
| **Rate of PVL/white matter injury** | NR | NR | N/A | NR |
| **Rate of Cerebral Palsy** | NR | NR | N/A | NR |
| **Intervention** | | | | |
| **Specific interventional concepts** | VIBeS Plus | | | |
| **Description of intervention** | - Problem-based learning approach using home-visits in the first year of life<br>- education about infant self-regulation by a psychologist<br>- teaching techniques to improve infant posture, strength and coordination by a physiotherapist | | | |
| **Aim of intervention:**<br>Improvement of... | - parent-infant interaction<br>- caregivers' mental health<br>- infants' musculosceletal development | | | |
| **Age of participants at start of intervention (years)** | 0-1 (recruitment at term equivalet age) | | | |
| **Setting** | Home-based | | | |
| **Number of sessions** | 9 | | | |
| **Frequency of sessions** | At 2 weeks, 4 weeks and at 2, 3, 4, 6, 8, 9, and 11 months' corrected age. | | | |
| **Duration of program** | 1 year | | | |

| Comparator | | |
|---|---|---|
| **Type of comparator** | | SoC |
| **Outcomes** | | **Assessment tool** |
| **Outcomes relevant for our study** | | |
| **Behaviour** | **24 months**<br>**4 years**<br><br>**8 years**<br><br><br>**13 years** | ITSEA<br>Behavior Assessment System for Children – Preschool version<br>Child Behaviour Checklist<br>Social Skills Improvements Rating Scale<br>Strengths and Difficulties Questionnaire<br>Test of Everyday Attention – Children<br>Behavior Rating Inventory of Executive Function, 2nd Edition<br>Strengths and Difficulties Questionnaire<br>Test of Everyday Attention – Children |
| **Language** | **24 months**<br>**4/8 years** | Bayley-III (too early)<br>Differential Abilities Scale, 2nd Edition – verbal composite score |
| **Executive functioning** | **8 years** | Tower Of London (spatial planning, behavioral inhibition) |
| **Executive function** | **13 years** | CNT arrow switching<br>D-KEFS Tower Test<br>Working Memory Test Battery for Children<br>Behavior Rating Inventory of Executive Function, 2nd Edition |
| **Outcomes *not* relevant for our study** | | |
| **Motor** | **24 months**<br>**4/8/13 years** | Bayley-III<br>Movement Assessment Battery for Children, 2nd Edition |
| **Cognition** | **24 months**<br>**4/8 years**<br>**13 years** | Bayley-III<br>Differential Ability Scale, 2nd Edition<br>Wechsler Abbreviated Scale of Intelligence, 2nd Edition<br>WRAT5 Reading, Spelling, Mathematical Computation |
| **Caregiver mental health** | **2/4/8/13 years** | Hospital Anxiety and Depression Scale (HADS) |

| | | |
|---|---|---|
| **Study ID** | Wu 2014 | |
| **DOI** | 10.1016/j.ridd.2014.06.009 | |
| **Title** | **A randomized controlled trial of clinic-based and home-based interventions in comparison with usual care for preterm infants: Effects and mediators** | |
| **Type of RCT** (parallel/multiple groups) | Multiple group study | |
| **Additional study reports available** | - Secondary study publication<br>- Trial registry record | |
| **Study objective:**<br>To examine ... | - developmental and behavioral outcomes<br>- the costs and effectiveness of the UCP, the CBIP, and the HBIP for VLBW preterm infants<br>- emotion regulation to stress | |
| **Setting** (single-/multi-centric) | multi-centric | |
| **Country in which the study conducted** | Taiwan | |
| **Start date** | xx/01/2006 | |
| **End date** | xx/10/2010 | |
| **Funding source** | - 2 grants from the National Health Research Institute (NHRI-EX98-9519PI, NHRI-EX102-10106PI)<br>- grant from the National Science Council in Taiwan (NSC98-2314-B-002-010-MY3) | |
| **Trial registration number(s)** | NCT00173108<br>NCT01952093 | |
| **Conflict of interest** | None | |
| **Inclusion criteria** | - BW below 1,501 gm<br>- GA under 37 weeks<br>- Admission to the NICU within the first 7 days of life<br>- Physiologically stable at PCA 36 weeks as diagnosed by attending physician<br>- Hospital discharge prior to PCA 40 weeks<br>- Absence of congenital anomalies and/or severe neonatal diseases<br>- Singleton or the first child of twin/multiple birth<br>- Family residence in the greater Taipei area | |
| **Exclusion criteria** | - Major neurologic abnormalities<br>- Necrotizing enterocolitis with colostomy<br>- Severe cardiopulmonary disease requiring daily oxygen use at hospital discharge | |
| **preterms as...** | Full population | |
| **gestational age (weeks)** | preterm (<37 weeks) | |
| **birth weight** | VLBW (< 1500g) | |
| **If multiple group study, define intervention 1, intervention 2, intervention 3** | **IG 1** | Clinic-based Intervention Program (CBIP) |
| | **IG 2** | Home-based Intervention Program (HBIP) |

| | | IG 3 | Usual Care | | |
|---|---|---|---|---|---|
| **Sociodemographic Characteristics:** | | **IG 1** | **IG 2** | **IG 3** | **total** |
| **sociodemographic characterstics of parents** | **education** | **Maternal education, n (%)**<br>College or above<br>42 (78)<br><br>High school<br>9 (17)<br><br>Below high school<br>3 (5)<br><br>**Paternal education, n (%)**<br>College or above<br>40 (74)<br><br>High school<br>11 (20)<br><br>Below high school<br>3 (6) | **Maternal education, n (%)**<br>College or above<br>34 (61)<br><br>High school<br>20 (36)<br><br>Below high school<br>2 (3)<br><br>**Paternal education, n (%)**<br>College or above<br>34 (61)<br><br>High school<br>19 (34)<br><br>Below high school<br>3 (5) | **Maternal education, n (%)**<br>College or above<br>39 (76)<br><br>High school<br>12 (24)<br><br>Below high school<br>0 (0)<br><br>**Paternal education, n (%)**<br>College or above<br>29 (58)<br><br>High school<br>18 (36)<br><br>Below high school<br>3 (6) | NR |
| | **income** | NR | NR | NR | NR |
| | **employment** | **Maternal occupation, n (%)**<br>Professional<br>19 (35)<br>Technician<br>15 (28)<br>Labor or housewife<br>20 (37)<br><br>**Paternal occupation, n (%)**<br>Professional<br>30 (56)<br>Technician<br>21 (39)<br>Labor or | **Maternal occupation, n (%)**<br>Professional<br>13 (23)<br>Technician<br>16 (29)<br>Labor or housewife<br>27 (48)<br><br>**Paternal occupation, n (%)**<br>Professional<br>20 (36)<br>Technician<br>27 (48)<br>Labor or | N/A | NR |

| | | | | |
|---|---|---|---|---|
| | unemployed<br>3 (5) | unemployed<br>9 (16) | | |
| **Marital/family status** | NR | NR | N/A | NR |
| **Number of participants allocated** | 57 | 63 | 58 | 178 |
| **baseline population characterstics** | **IG 1** | **IG 2** | **IG 3** | **total** |
| **No. of participants** | 54 | 56 | 51 | 161 |
| **Mean birthweight (SD) in gramme** | 1179 (228) | 1149 (283) | 1091 (268) | NR |
| **Mean gestational age (SD) in weeks** | 30.0 (2.6) | 29.9 (3.2) | 29.3 (2.7) | NR |
| **No. of male/female** | 29/25 | 31/25 | 24/27 | 84/77 |
| **percentage of male/female (in %)** | 54/46 | 55/45 | 47/53 | 52.2/47.8 |
| **multiples** | NR | NR | NR | NR |
| **Rate of BPD** | NR | NR | NR | NR |
| **Rate of IVH** | 0 | 0 | 0 | 0 |
| **Rate of PVL/white matter injury** | 0 | 0 | 0 | 0 |
| **Rate of Cerebral Palsy, n(%)** | 0 | 1 (2) | 3 (7) | 4 (2) |

| | | | |
|---|---|---|---|
| **Intervention** | | | |
| **Specific interventional concepts** | | CBIP, HBIP | |
| **Description of intervention** | | in-hospital:<br>- Based on Synactive Theory[1] and Family-Centered Care[2]<br>- Applied by a nurse and a physical therapist<br>after-discharge:<br>- Based on the Biosocial Systems Theory[3]<br>- 8 sessions<br>- Either at the hospital (CBIP), or at home (HBIP)<br>- Applied by a physical therapist | |
| **Aim of intervention:**<br>Improvement of… | | - Handling of the newborn<br>- neurobiological health of infant and caregivers<br>- parent-child interaction | |
| **Age of participants at start of intervention (years)** | | NR | |
| **Setting** | | NR | |
| **Number of sessions** | | NR | |
| **Frequency of sessions** | | NR | |
| **Duration of program** | | NR | |
| **Comparator** | | | |
| **Type of comparator** | | SoC | |
| **Outcomes** | | **Assessment tool** | |
| **Outcomes relevant for our study** | | | |
| **Behavior** | **24 months** | CBCL 1.5-5 | |
| **Outcomes *not* relevant for our study** | | | |
| **Parent-child interaction** | **12/18/24 months** | Toy-behind-barrier procedure,<br>Free-play procedure | not poolable |
| **Participation** | **24 months** | Bayley-III | too early |
| **Motor** | **24 months** | Bayley-III | |

[1] Als et al., 1986
[2] Dunn et al., 2006
[3] Ramey et al., 1984

| | | |
|---|---|---|
| **Cognition** | **24 months** | Bayley-III |
| **Neurosensory impairments** | | Determined by pediatric neurologists, ophthalmologists, otolaryngologists |

| | | | | | |
|---|---|---|---|---|---|
| **Study ID** | | Youn 2021 | | | |
| **DOI** | | 10.3390/brainsci11050575 | | | |
| **Title** | | **Preventive Intervention Program on the Outcomes of Very Preterm Infants and Caregivers: A Multicenter Randomized Controlled Trial** | | | |
| **Type of RCT** (parallel/multiple groups) | | Parallel group study | | | |
| **Additional study reports available** | | Trial registry record without results | | | |
| **Study objective:**<br>To examine ... | | - neurodevelopmental and behavioral outcomes<br>- parent-infant interaction<br>- maternal wellbeing | | | |
| **Setting** | | multi-centric | | | |
| **Country in which the study conducted** | | Republic of Korea | | | |
| **Start date** | | xx/03/2015 | | | |
| **End date** | | xx/02/2019 | | | |
| **Funding source** | | Korea Health Technology R&D Project through the Korea Health Industry Development Institute (KHIDI), funded by the Ministry of Public Health & Welfare, Republic of Korea (HI14C3451) | | | |
| **Trial registration number(s)** | | NCT02415530 | | | |
| **Conflict of interest** | | None | | | |
| **Inclusion criteria** | | GA ≤ 30 weeks or BW ≤ 1500 g | | | |
| **Exclusion criteria** | | Congenital neuromuscular diseases, cardiac anomalies, or chromosomal anomalies | | | |
| **preterms as...** | | full population | | | |
| **gestational age (weeks)** | | </= 30 weeks | | | |
| **birth weight** | | VLBW (< 1500g) | | | |
| **Definition of intervention 1, intervention 2, intervention 3** | | **IG 1** | Intervention | | |
| | | **IG 2** | Control | | |
| | | **IG 3** | N/A | | |
| **Sociodemographic Characteristics:** | | **IG 1** | **IG 2** | **IG 3** | **total** |
| **sociodemographic characterstics of parents** | **education** | College education or above<br><br>Father:<br>59 (85.5)<br><br>Mother:<br>IG 59 (85.5) | College education or above<br><br>Father:<br>CG 63 (94)<br><br>Mother:)<br>CG 61 (91) | N/A | NR |
| | **income** | in thousand won:<br><br><2000<br>2 (2.9%)<br><br>2000-4000<br>30 (43.5%)<br><br>>4000<br>37 (53.6%) | in thousand won:<br><br><2000<br>3 (4.5%)<br><br>2000-4000<br>CG 30 (44.8%)<br><br>>4000<br>34 (50.8%) | N/A | NR |

| | | | | |
|---|---|---|---|---|
| **employment** | salaried employee<br>IG 19 (27.5)<br><br>public officials<br>IG 1 (1.5)<br><br>self-employed<br>IG 1 (1.5)<br><br>professional<br>IG 9 (13)<br><br>others<br>IG 39 (56.5) | salaried employee<br>CG 14 (20.9)<br><br>public officials<br>CG 0 (0)<br><br>self-employed<br>CG 3 (4.5)<br><br>professional<br>CG 6 (9)<br><br>others<br>CG 44 (65.7) | N/A | NR |
| **Marital/family status** | Marriage:<br>IG 69 (100%) | Marriage:<br>CG 66 (98.5%) | N/A | NR |
| **Number of participants allocated** | 69 | 69 | N/A | 138 |
| **baseline population characterstics** | **IG 1** | **IG 2** | **IG 3** | **total** |
| **No. of participants** | 69 | 67 | N/A | 136 |
| **Mean birthweight (SD) in gramme** | 1145.5 (344.5) | 1188.9 (340.6) | N/A | NR |
| **Mean gestational age (SD) in weeks** | 29 (2.6) | 29 (2.5) | N/A | NR |
| **No. of male/female** | 31/38 | 36/31 | N/A | 67/69 |
| **percentage of male/female (in %)** | 43.3/56.7 | 55.1/44.9 | N/A | 49.3/50.7 |
| **multiples** | 26 (38.8%) | 31 (44.9%) | N/A | 57 (42.3%) |
| **Rate of BPD** | 18 (26.1%) | 21 (31.8%) | N/A | 39 (28.7%) |
| **Rate of IVH** | 26 (37.7%) low-grade IVH,<br><br>9 (13%) severe injury | 21 (31.3%) low-grade IVH,<br>13 (19.4%) severe injury | N/A | 47 (34.6%) |
| **Rate of PVL/white matter injury** | NR | NR | N/A | NR |
| **Rate of Cerebral Palsy** | NR | NR | N/A | NR |
| **Intervention** | | | | |
| **Specific interventional concepts** | - Mother–Infant Transaction Program (MITP) based on IHDP | | | |
| **Description of intervention** | - home visits by experiences NICU nurses<br>- group physiotherapy by a physiotherapist specialized in infant neurodevelopment and a pediatric physiotherapist | | | |
| **Aim of intervention:**<br>Improvement of… | - infant's growth and neurodevelopment<br>- infants' gross motor development<br>- infants' sensory stimulation<br>- parent-child attachement<br>- reading of infants' behavioral cues | | | |
| **Age of participants at start of intervention (years)** | 0-1 | | | |
| **Setting** | home-visits; community-centre or child-care centre | | | |
| **Number of sessions** | up to 15 | | | |
| **Frequency of sessions** | - 4 home visits 5 days, 2 weeks, and 1 month after discharge, and at 2 months CA<br>- up to 12 group interventions of physical therapy before 6 months CA | | | |
| **Duration of program** | from birth to 6 months corrected age (CA) | | | |
| **Comparator** | | | | |

| | | | |
|---|---|---|---|
| **Type of comparator** | | SoC | |
| **Outcomes** | | **Assessment Tool** | |
| **Outcomes relevant for our study** | | | |
| **Behavior** | **24 months** | Modified Infant and Toddler Social and Emotional Assessment (m-ITSEA) | |
| **Parent-child interaction** | **2/6 months** | Mother-to-Child Attachment (MCA) score | |
| **Outcomes *not* relevant for our study** | | | |
| **Behavior (temperament)** | **10 months** | Infant Characteristics Questionnaire (ICQ) | |
| **Participation (language)** | **10 months** | Bayley-III | too early |
| **Cognition** | **24 months** | Bayley-III<br>Korean Developmental Screening Test (K-DST) | |
| **Motor** | **24 months** | Bayley-III<br>Korean Developmental ScreeningTest (K-DST) | |
| **Parental well-being** | **2/6 months** | Center for Epidemiologic Studies Depression Scale (CES-D) | |

#### S2.3 – Studies with narrative outcomes

| | |
|---|---|
| **Study ID** | Parker-Loewen 1987 |
| **DOI** | 10.1002/1097-0355(198723)8:3<277::AID-IMHJ2280080310>3.0.CO;2-X |
| **Title** | **Effects of short-term interaction coaching with mothers of preterm infants.** |
| **Type of RCT** (parallel/multiple groups) | Parallel group study |
| **Additional study reports available** | None identified |
| **Study objective:**<br>To examine … | …the effects of eight 40-minute interaction coaching sessions on the mother- infant interaction patterns of mother-preterm infant dyads. |
| **Setting** (single-/multi-centric) | multi-centric |
| **Country in which the study conducted** | Canada |
| **Start date** | xx/04/1983 |
| **End date** | xx/03/1984 |
| **Funding source** | Health and Welfare Canada |
| **Trial registration number(s)** | NR |
| **Conflict of interest** | NR |
| **Inclusion criteria** | - GA < 37 weeks<br>- BW 1000-2000 g<br>- Admission to three major hospitals in Calgary, Alberta, between 04/1983 and 03/1984 |
| **Exclusion criteria** | NR |
| **preterms as…** | Full population |
| **gestational age (weeks)** | preterm (<37 weeks) |
| **birth weight** | 1000-2000g |

| | | |
|---|---|---|
| **If multiple group study, define intervention 1, intervention 2, intervention 3** | **IG 1** | Intervention |
| | **IG 2** | control |
| | **IG 3** | N/A |

| **Sociodemographic Characteristics:** | | **IG 1** | **IG 2** | **IG 3** | **total** |
|---|---|---|---|---|---|
| **sociodemographic charactestics of parents** | **education**<br><br>from 4 (university graduate) to 1 (less than high school graduate | Mother<br>2.50 (1.10) | Mother:<br>2.53 (1.07) | N/A | Mother:<br>2.51 (1.07)<br><br>primarily high school graduates |
| | **income** | NR | NR | N/A | NR |
| | **employment** | NR | NR | N/A | NR |

| | | | | |
|---|---|---|---|---|
| **marital status** | NR | NR | N/A | Mothers were primarily married |
| **Number of participants allocated** | 18 | 17 | N/A | 35 |
| **baseline population characterstics** | **IG 1** | **IG 2** | **IG 3** | **total** |
| **No. of participants** | 18 | 17 | N/A | 35 |
| **Mean birthweight (SD) in gramme** | 1596.94 (293.38) | 1749.94 (248.72) | N/A | 1671.14 (279.55) |
| **Mean gestational age (SD) in weeks** | 32.39 (2.45) | 32.59 (1.87) | N/A | 32.49 (2.16) |
| **No. of male/female** | NR | NR | N/A | NR |
| **percentage of male/female (in %)** | NR | NR | N/A | NR |
| **multiples** | NR | NR | N/A | NR |
| **Rate of BPD** | NR | NR | N/A | NR |
| **Rate of IVH** | NR | NR | N/A | NR |
| **Rate of PVL/white matter injury** | NR | NR | N/A | NR |
| **Rate of Cerebral Palsy** | NR | NR | N/A | NR |
| **Intervention** | | | | |
| **Specific interventional concepts** | None | | | |
| **Description of intervention** | - 40min training sessions<br>- Video-taped “playing”/interaction of mother and child<br>- 10min normal interaction with child, 20 minutes interaction with earpiece microphone, 10min free interaction time again<br>- Trainer offered advice regarding the interaction via earpiece | | | |
| **Aim of intervention:**<br>Improvement of**…** | - Mothers’ responsiveness and sensitivity to child cues<br>- Satisfaction in being a parent<br>- Knowledge of infant development | | | |
| **Age of participants at start of intervention (years)** | 0-1<br><br>IG: 12.78 (1.59) weeks<br>CG: 13.00 (2.21) weeks<br>total: 12.89 (1.89) weeks | | | |
| **Setting** | outpatient | | | |
| **Number of sessions** | 8 | | | |
| **Frequency of sessions** | NR | | | |
| **Duration of program** | 12-15 weeks | | | |
| **Comparator** | | | | |
| **Type of comparator** | SoC | | | |
| **Outcomes** | **Assessment tool** | | | |
| **Outcomes relevant for our study but *without* eligible assessment tool for meta-analysis** | | | | |
| **Parent-child interaction** **prior to/immediately after/2 months after treatment** | adapted version of the Interaction Rating Scale (IRS; Field, 1980) | | ≠ Silveira 2016 | |

| | | | | | |
|---|---|---|---|---|---|
| **Study ID** | Resnick 1988 | | | | |
| **DOI** | NR | | | | |
| **Title** | **Developmental intervention program for high-risk premature infants: effects on development and parent-infant interactions** | | | | |
| **Type of RCT (parallel/multiple groups)** | Parallel group study | | | | |
| **Additional study reports available** | None identified | | | | |
| **Study objective:**<br>**To examine ...** | - the development of high-risk, preterm infants.<br>- the quality of communication between infants and their caregivers. | | | | |
| **Setting (single-/multi-centric)** | single-centric | | | | |
| **Country in which the study conducted** | United States | | | | |
| **Start date** | NR | | | | |
| **End date** | NR | | | | |
| **Funding source** | Jessie Ball Dupont Foundation, Nationale Foundation of the March of Dimes, State of Florida's Children's Medical Services, Developmental Services, and Developmental Disabilities Council | | | | |
| **Trial registration number(s)** | NR | | | | |
| **Conflict of interest** | NR | | | | |
| **Inclusion criteria** | - BW < 1800g<br>- Admission to NICU within 24h of birth<br>- Living within the state's Health and Rehabilitative Services District III | | | | |
| **Exclusion criteria** | NR | | | | |
| **preterms as...** | Full population | | | | |
| **gestational age (weeks)** | Preterm (<37 weeks) | | | | |
| **birth weight** | < 1800g | | | | |
| **If multiple group study, define intervention 1, intervention 2, intervention 3** | **IG 1** | Intervention | | | |
| | **IG 2** | control | | | |
| | **IG 3** | N/A | | | |
| **Sociodemographic Characteristics:** | | **IG 1** | **IG 2** | **IG 3** | **total** |
| **sociodemographic characterstics of parents** | **education** | Mothers education: 11.4 (3.4)y<br><br>Fathers education: 10.7 (4.7)y | Mothers education: 12.4 (1.8)y<br><br>Fathers education: 10.3 (5.1)y | N/A | NR |
| | **income** | NR | NR | N/A | NR |
| | **employment** | NR | NR | N/A | NR |
| | **marital status** | NR | NR | N/A | NR |
| **Number of participants allocated** | | 21 | 20 | N/A | 41 |
| **baseline population characterstics** | | **IG 1** | **IG 2** | **IG 3** | **total** |
| **No. of participants** | | 21 | 20 | N/A | 41 |

| | | | | |
|---|---|---|---|---|
| **Mean birthweight (SD) in gramme** | 1427 (278) | 1321 (311) | N/A | NR |
| **Mean gestational age (SD) in weeks** | 31.5 (2.6) | 31.6 (2.8) | N/A | NR |
| **No. of male/female** | 11/10 | 8/12 | N/A | 19/22 |
| **percentage of male/female (in %)** | 52.4/47.6 | 40/60 | N/A | 46.3/53.7 |
| **multiples** | 1 | 0 | N/A | 1 |
| **Rate of BPD** | NR | NR | N/A | NR |
| **Rate of IVH** | NR | NR | N/A | NR |
| **Rate of PVL/white matter injury** | NR | NR | N/A | NR |
| **Rate of Cerebral Palsy** | NR | NR | N/A | NR |

| | |
|---|---|
| **Intervention** | |
| **Specific interventional concepts** | None |
| **Description of intervention** | NICU:<br>- two developmental interventions per day<br>- applied by postmaster's level graduate students specializing in early childhood development<br>home-visits:<br>- weekly by pediatric nurse practitioner until the infant reached adjusted birth date<br>- thereafter by an early childhood development specialist until infant reached adjusted age of 12 months<br>- curriculum of 160 activities |
| **Aim of intervention:**<br>Improvement of**...** | - language<br>- social skill<br>- cognitive development and spatial concept<br>- muscular development: small motor exercises<br>- muscular development: large motor exercises<br>- parenting activities |
| **Age of participants at start of intervention (years)** | 0-1 |
| **Setting** | started inpatient in NICU;<br>home-visits |
| **Number of sessions** | >24 |
| **Frequency of sessions** | NICU: min. 2/day home-based:<br>- until adjusted birth date: 1/week<br>- until CA 12 months: 2/month |
| **Duration of program** | from birth until 12 months' adjusted age |
| **Comparator** | |
| **Type of comparator** | SoC |
| **Outcomes** | **Assessment tool** |
| **Outcomes relevant for our study but *without* eligible assessment tool for meta-analysis** | |
| **Parent-child interaction** **6 months** **1 year** | Greenspan-Lieberman Observation Scale |
| **Outcomes *not* relevant for our study** | |
| **Cognition** **6 months** **1 year** | Bayley MDI |
| **Motor** **6 months** **1 year** | Bayley PDI |

| | |
|---|---|
| **Study ID** | Castel 2016 |
| **DOI** | 10.1016/j.earlhumdev.2016.05.007 |
| **Title** | **Effects of an intervention program on maternal and paternal parenting stress after preterm birth: A randomized trial** |
| **Type of RCT** (parallel/multiple groups) | Parallel group study |
| **Additional study reports available** | Trial registry record without results |
| **Study objective:**<br>To examine ... | - the effect on parenting stress and parental mental health.<br>- the effect on preterm infant development in the motor, language, social, behavioral and emotional domains. |
| **Setting** (single-/multi-centric) | single-centric |
| **Country in which the study conducted** | France |
| **Start date** | 01/06/2006 |
| **End date** | 30/08/2008 |
| **Funding source** | Hospital Program for Clinical Research (DGS: 2006/0215);<br>Wyeth Foundation |
| **Trial registration number(s)** | NCT02394444 |
| **Conflict of interest** | None |
| **Inclusion criteria** | Infants:<br>- GA 28+0 - 35+6<br>- no congenital anomalies o<br>- any other foreseeable disabilities during the neonatal period<br>- Siblings were not excluded<br>Parents:<br>- French speakers<br>- Age > 18 years<br>- without known psychiatric history<br>- Residing within a 50-km radius of the hospital |
| **Exclusion criteria** | - Congenital anomalies<br>- Any other foreseeable disabilities during the neonatal period |
| **preterms as...** | Full population |
| **gestational age (weeks)** | 28+0 - <35+6 weeks |
| **birth weight** | NR |

| | | |
|---|---|---|
| **If multiple group study, define intervention 1, intervention 2, intervention 3** | **IG 1** | Intervention |
| | **IG 2** | control |
| | **IG 3** | N/A |

| **Sociodemographic Characteristics:** | **IG 1** | **IG 2** | **IG 3** | **total** |
|---|---|---|---|---|
| **education** | <A level: | <A level: | N/A | NR |

| | | | | |
|---|---|---|---|---|
| **sociodemographic characterstics of parents** | 11 (33.3%)<br><br>A level-Bachelor: 16 (48.5%)<br><br>>Bachelor: 6 (18.2%) | 7 (21.9%)<br><br>A level-Bachelor: 15 (46.9%)<br><br>>Bachelor: 10 (31.3%) | | |
| **income** | NR | NR | N/A | NR |
| **employment** | employed: 24 (72.7%) | employed: 28 (87.5%) | N/A | NR |
| **marital status** | Married: 10 (30.3%) | Married: 16 (50.0%) | N/A | NR |
| **Number of participants allocated** | 40 | 35 | N/A | 75 |
| **baseline population characterstics** | **IG 1** | **IG 2** | **IG 3** | **total** |
| **No. of participants** | 40 | 35 | N/A | 75 |
| **Mean birthweight (SD) in gramme** | 1689 (469) | 1915 (459) | N/A | NR |
| **Mean gestational age (SD) in weeks** | 31.7 (2.7) | 32.5 (2.0) | N/A | NR |
| **No. of male/female** | 15/25 | 18/16 | N/A | 33/41 |
| **percentage of male/female (in %)** | 37.5/62.5 | 52.9/47.1 | N/A | 44/56 |
| **multiples** | 7 | 3 | N/A | 10 |
| **Rate of BPD** | NR | NR | N/A | NR |
| **Rate of IVH** | NR | NR | N/A | NR |
| **Rate of PVL/white matter injury** | NR | NR | N/A | NR |
| **Rate of Cerebral Palsy** | NR | NR | N/A | NR |
| **Intervention** | | | | |
| **Specific interventional concepts** | Triadic parent-infant Relationship Therapy (TRT) | | | |
| **Description of intervention** | - Conducted by a psychologist<br>- Promoting PCI (mother-father-infant triad)<br>- Parent education about child development | | | |
| **Aim of intervention:** Improvement of**...** | - Reduce parental stress<br>- infant's cognitive, motor, socio-emotional and behavioral development | | | |
| **Age of participants at start of intervention (years)** | 0-1 | | | |
| **Setting** | Outpatient;<br>Home-visits | | | |
| **Number of sessions** | 22 | | | |
| **Frequency of sessions** | -home visits: twice per month during the first four months<br>-monthly consultations in the neonatology ward (5-18 months CA) | | | |
| **Duration of program** | up to 18 months CA | | | |
| **Comparator** | | | | |
| **Type of comparator** | SoC | | | |
| **Outcomes** | **Assessment tool** | | | |
| **Outcomes relevant for our study but *without* eligible assessment tool for meta-analysis** | | | | |
| **PCI 3/18 months** | Parental Stress Index (short form) | | Not poolable | |

| | | | |
|---|---|---|---|
| **Behaviour** | | Neonatal Behavioral Assessment Scale (NBAS) | Not poolable |
| **Behaviour** | **9 months**<br>**18 months** | Infant Behavioral Symptom Check-List | Not externalizing/ internalizing |
| **Participation (language, socialization)** | **3 months**<br>**9 months**<br>**18 months** | Brunet-Lézine Revised test (BLR) | Too early (<4y CA) |
| **Outcomes *not* relevant for our study** | | | |
| **Caregiver mental health** | **Baseline**<br>**3 months**<br>**9 months**<br>**18 months** | Edinburgh Postnatal Depression Scale (EPDS) | |
| **Caregiver mental health** | **Baseline**<br>**18 months** | Perinatal Post-Traumatic Stress Disorder (PTSD) Scale | |
| **Global Infant Development** | **3 months**<br>**9 months**<br>**18 months** | Brunet-Lézine Revised test (BLR) – Global /Motor/Coordination Score | |

| | |
|---|---|
| **Study ID** | Silveira 2016 |
| **DOI** | 10.1001/jamanetworkopen.2024.2189 |
| **Title** | **Early Intervention Program for Preterm Infants and Their Parents: establishing the Impact at 18 Months Corrected Age** |
| **Type of RCT (parallel/multiple groups)** | Parallel group study |
| **Additional study reports available** | Secondary journal publication, study protocol, trial registry record without results |
| **Study objective:**<br>**To examine ...** | - body composition of preterm infants<br>- neurodevelopmental outcomes of preterm infants |
| **Setting (single-/multi-centric)** | single-centric |
| **Country in which the study conducted** | Brazil |
| **Start date** | 01/01/2016 |
| **End date** | 31/05/2022 |
| **Funding source** | grant OPP1142172 from the Bill & Melinda Gates Foundation |
| **Trial registration number(s)** | NCT02835612 |
| **Conflict of interest** | None |
| **Inclusion criteria** | - GA < 32 weeks or BW < 1500g<br>- Residence within 40 km of the birth hospital |
| **Exclusion criteria** | - major congenital malformations<br>- metabolic conditions<br>- congenital infections<br>- autoimmune conditions |
| **preterms as...** | Full population |
| **gestational age (weeks)** | < 32 |
| **birth weight** | < 1500g (VLBW) |

| | | |
|---|---|---|
| **If multiple group study, define intervention 1, intervention 2, intervention 3** | **IG 1** | Intervention |
| | **IG 2** | control |
| | **IG 3** | N/A |

| **Sociodemographic Characteristics:** | **IG 1** | **IG 2** | **IG 3** | **total** |
|---|---|---|---|---|
| **education**<br><br>**sociodemographic characterstics of parents** | **Maternal educational level, No. (%)**<br>Incomplete middle school<br>6 (12)<br><br>Complete middle school<br>6 (12)<br><br>Incomplete high school<br>5 (10)<br><br>High school degree<br>21 (42) | **Maternal educational level, No. (%)**<br>Incomplete middle school<br>7 (14)<br><br>Complete middle school<br>13 (26)<br><br>Incomplete high school<br>4 (8)<br><br>High school degree<br>16 (32) | N/A | NR |

| | | | | | |
|---|---|---|---|---|---|
| | | Incomplete undergraduate<br>7 (14)<br><br>Undergraduate degree<br>5 (10)<br><br>**Paternal educational level, No. (%)**<br>Incomplete middle school<br>9 (18)<br><br>Complete middle school<br>7 (14)<br><br>Incomplete high school<br>5 (10)<br><br>High school degree<br>20 (40)<br><br>Incomplete undergraduate<br>2 (4)<br><br>Undergraduate degree<br>6 (12)<br><br>Graduate degree<br>1 (2) | Incomplete undergraduate<br>6 (12)<br><br>Undergraduate degree<br>2 (4)<br><br>**Paternal educational level, No. (%)**<br>Incomplete middle school<br>8 (16)<br><br>Complete middle school<br>14 (28)<br><br>Incomplete high school<br>5 (10)<br><br>High school degree<br>22 (44)<br><br>Incomplete undergraduate<br>0<br><br>Undergraduate degree<br>3 (6)<br><br>Graduate degree<br>0 | | |
| | **income** | Monthly (US-Dollar)<br>436 (280) | Monthly (US-Dollar)<br>419 (313) | N/A | NR |
| | **employment** | NR | NR | N/A | NR |
| | **marital status** | **Parents live together, No. (%)**<br><br>Yes<br>39 (78)<br><br>No<br>11 (22) | **Parents live together, No. (%)**<br><br>Yes<br>43 (86)<br><br>No<br>7 (14 | N/A | NR |

| Number of participants allocated | NR | NR | N/A | NR |
|---|---|---|---|---|
| **baseline population characterstics** | **IG 1** | **IG 2** | **IG 3** | **total** |
| **No. of participants** | 50 | 50 | N/A | 100 |
| **Mean birthweight (SD) in gramme** | 1115 (286) | 1077 (318) | N/A | 1096 (300) |
| **Mean gestational age (SD) in weeks** | 28.3 (2.3) | 28.5 (2.2) | N/A | 28.4 (2.2) |
| **No. of male/female** | 29/21 | 28/22 | N/A | 57/43 |
| **percentage of male/female (in %)** | 58/42 | 56/44 | N/A | NR |
| **multiples** | NR | NR | N/A | NR |
| **Rate of BPD** | NR | NR | N/A | 32 |
| **Rate of IVH** | NR | NR | N/A | NR |
| **Rate of PVL/white matter injury** | NR | NR | N/A | 35 |
| **Rate of Cerebral Palsy** | NR | NR | N/A | NR |
| **Intervention** | | | | |
| **Specific interventional concepts** | - | | | |
| **Description of intervention** | - NICU: tactile-kinesthetic stimulation and kangaroo care<br>- Post-discharge:<br>- Ten at-home professional stimulation sessions<br>- Ten home visits (multidisciplinary team orientation, monthly stimulus book, and monthly toy kit)<br>- monthly standard of care at follow-up clinic | | | |
| **Aim of intervention:** | Improvement of**...**<br>- neurodevelopment<br>- parent-child relationship<br>Investigation of...<br>- body composition<br>- neonatal growth<br>- biochemical charateristics | | | |
| **Age of participants at start of intervention** | 7 days postnatal | | | |
| **Setting** | Started in NICU;<br>Outpatient;<br>Home-visits | | | |
| **Number of sessions** | 10 at-home professional stimulation sessions;<br>10 home visits | | | |
| **Frequency of sessions** | NR | | | |
| **Duration of program** | 18 months | | | |
| **Comparator** | | | | |
| **Type of comparator** | NICU: kangaroo care<br>post-discharge: routine visits to the follow-up clinic | | | |
| **Outcomes** | **Assessment tool** | | | |
| **Outcomes relevant for our study** | | | | |
| **Parent-Child Interaction** **12 months** | Interaction Rating Scale | | | ≠ Parker-Loewen 1987 |
| **Outcomes *not* relevant for our study** | | | | |
| **Body Composition** **5 years** | bioelectrical impedanceanalyses,<br>physical activity and feeding practices questionnaires | | | |
| **Cognition** **4/8/12/18 months** | BSID-III | | | |
| **Motor Outcomes** **4/8/12/18 months** | Alberta Infant Motor Scale<br>BSID-III Motor Scale | | | |
| **Participation (language)** **4/8/12/18 months** | BSID-III Language Scale | | | Too early |

### S2.4 – Studies without eligible outcomes

***Table 1.2: Characteristics of included studies, that were not eligible for metanalysis***

| | | |
|---|---|---|
| **Study ID** | Sinha 2021 | |
| **DOI** | 10.4269/ajtmh.21-0877 | |
| **Title** | **Effect of Community-Initiated Kangaroo Mother Care on Fecal Biomarkers of Gut Function inLow Birth Weight Infants in North India: A Randomized Clinical Trial** | |
| **Type of RCT** (parallel/multiple group) | Parallel group study | |
| **Additional study reports available** | None identified | |
| **Study objective:**<br>To examine … | - individual fecal biomarkers<br>- enteric enteropathy | |
| **Setting** (single-/multi-centric) | multi-centric | |
| **Country in which the study conducted** | India | |
| **Start date** | xx/05/2017 | |
| **End date** | xx/10/2017 | |
| **Funding source** | Science and Engineering Research Board (SERB), a statutory body of the Department of Science and Technology, Government of India (File No. EMR/2017/003414);<br>Centre for Intervention Science in Maternal and Child Health (CISMAC; project number 223269), funded by the Research Council of Norway through its Centres of Excellence scheme and the University of Bergen (UiB), Norway | |
| **Trial registration number(s)** | ClinicalTrials.gov NCT02653534<br>Clinical trials registry-India CTRI/2017/04/008430 | |
| **Conflict of interest** | None | |
| **Inclusion criteria** | - BW 1,500–2,250 g<br>- Screening within 72 hours of birth<br>- One eligible child per household | |
| **Exclusion criteria** | - Kangaroo Mother Care already initiated in a birth facility<br>- Infants unable to feed<br>- Breathing problems<br>- Gross congenital malformations<br>- Less than normal movements<br>- Mothers not living with their babies or intending to move away over the next 6 months<br>- Twins and triplets | |
| **preterms as...** | subgroup | |
| **gestational age (weeks)** | preterm (<37 weeks) | |
| **birth weight** | 1500-2250g | |
| **If multiple group study, define intervention 1, intervention 2, intervention 3** | **IG 1** | Intervention |
| | **IG 2** | Control |

| | | | | |
|---|---|---|---|---|
| | IG 3 | N/A | | |
| **Sociodemographic Characteristics:** | **IG 1** | **IG 2** | **IG 3** | **total** |
| **sociodemographic characterstics of parents** — **education** | **Maternal education**<br>years of schooling, n(SD)<br>5.7 (5.2) | **Maternal education**<br>years of schooling, n(SD)<br>7.1 (5.3) | N/A | NR |
| **income** | **Wealth Quintiles**<br>Least poor<br>16<br>Less poor<br>27<br>Poor<br>26<br>Very poor<br>13<br>Most poor<br>18 | **Wealth Quintiles**<br>Least poor<br>19<br>Less poor<br>25<br>Poor<br>17<br>Very poor<br>25<br>Most poor<br>14 | N/A | NR |
| **employment** | NR | NR | N/A | NR |
| **marital status** | NR | NR | N/A | NR |
| **Number of participants allocated** | 100 | 100 | N/A | 200 |
| **baseline population characterstics** | **IG 1** | **IG 2** | **IG 3** | **total** |
| **No. of participants** | 100 | 100 | N/A | 200 |
| **Mean birthweight (SD) in gramme** | 2086.2 (139.1) | 2094.5 (162.1) | N/A | NR |
| **Mean gestational age (SD) in weeks** | 35.9 (1.6) | 35.9 (1.9) | N/A | NR |
| **No. of male/female** | 41/59 | 48/52 | N/A | 89/111 |
| **percentage of male/female (in %)** | 41/59 | 48/52 | N/A | 44.5/55.5 |
| **multiples** | 0 | 0 | N/A | 0 |
| **Rate of BPD** | NR | NR | N/A | NR |
| **Rate of IVH** | NR | NR | N/A | NR |
| **Rate of PVL/white matter injury** | NR | NR | N/A | NR |
| **Rate of Cerebral Palsy** | NR | NR | N/A | NR |
| **Intervention** | | | | |
| **Specific interventional concepts** | Community-initiated Kangaroo Mother Care | | | |
| **Description of intervention** | - Home visits<br>- Promotion of skin to skin care (SSC) and exclusive breastfeeding | | | |
| **Aim of intervention:**<br>Improvement of... | - Gut inflammation and permeability by prolonged SSC | | | |
| **Age of participants at start of intervention (years)** | 0-1; median (IQR) age 27.5 (12.5–38.5) hours. | | | |
| **Setting** | Home-visits | | | |
| **Number of sessions** | At least 9 | | | |
| **Frequency of sessions** | on 1, 2, 3, 5, 7, 10, 14, 21, and 28 days after birth | | | |
| **Duration of program** | Visits continued till 28 days | | | |
| **Comparator** | | | | |

| | |
|---|---|
| **Type of comparator** | SoC (home-based postnatal care visits by ASHAs as implemented through the health system) |
| **Outcomes** | **Assessment tool** |
| **Outcomes *not* relevant for our study** | |
| **intestinal health** **1 month** | Fecal biomarkers (Neopterin, Myeloperoxidase, Alpha-1-Antitrypsin) |

| | |
|---|---|
| **Study ID** | Sgandurra 2017 |
| **DOI** | 10.1371/journal.pone.0173521 |
| **Title** | **A randomized clinical trial in preterm infants on the effects of a home-based early intervention with the 'CareToy System'** |
| **Type of RCT** (parallel/multiple groups) | Parallel group study |
| **Additional study reports available** | - Secondary journal pulication (e.g. follow-up study)<br>- Study protocol<br>- Trial registry record without results |
| **Study objective:**<br>To examine ... | - Early motor development<br>- Visual development<br>- Parenting stress<br>- Parent-child interaction |
| **Setting** (single-/multi-centric) | multi-centric |
| **Country in which the study conducted** | Italy<br>Denmark |
| **Start date** | xx/07/2013 |
| **End date** | xx/12/2015 |
| **Funding source** | European Union under the Seventh Framework Program, Grant ICT-2011.5.1-287932 |
| **Trial registration number(s)** | NCT01990183 |
| **Conflict of interest** | None |
| **Inclusion criteria** | - GA 28+0 - 32+6<br>- Children with 3–9 months of CA who had achieved a predefined cut-off score in gross motor ability derived from ASQ-3 |
| **Exclusion criteria** | - BW < 10th percentile<br>- Brain damage i.e. intra-ventricular haemorrhage < grade 1, any degree of periventricular leukomalacia, or brain malformation<br>- Any form of seizure or epilepsy<br>- Severe sensory deficits (blindness, deafness)<br>- Severe non-neurological malformations<br>- Participation in other experimental rehabilitation studies |
| **preterms as...** | Full population |
| **gestational age (weeks)** | 28+0 - 32+6 |
| **birth weight** | NR |

| | | |
|---|---|---|
| **If multiple group study, define intervention 1, intervention 2, intervention 3** | **IG 1** | Intervention |
| | **IG 2** | control |
| | **IG 3** | N/A |

| **Sociodemographic Characteristics:** | **IG 1** | **IG 2** | **IG 3** | **total** |
|---|---|---|---|---|

| | | | | | |
|---|---|---|---|---|---|
| sociodemographic characterstics of parents | education<br>income<br>employment<br>marital status | NR<br>NR<br>NR<br>no. of married 17 | NR<br>NR<br>NR<br>no. of married<br>CG 13 | N/A<br>N/A<br>N/A<br>N/A | NR<br>NR<br>NR<br>no. of married 30 |
| **Number of participants allocated** | | 19 | 22 | N/A | 41 |
| **baseline population characterstics** | | **IG 1** | **IG 2** | **IG 3** | **total** |
| **No. of participants** | | 19 | 22 | N/A | 41 |
| **Mean birthweight (SD) in gramme** | | 1368.3 (330.3) | 1459.6 (275.6) | N/A | NR |
| **Mean gestational age (SD) in weeks** | | 30.7 (1.4) | 30.82 (1.1) | N/A | NR |
| **No. of male/female** | | 8/11 | 11/11 | N/A | 19/22 |
| **percentage of male/female (in %)** | | 42.11/57.89 | 50/50 | N/A | 46.34/53.66 |
| **multiples** | | 8 | 10 | N/A | 18 |
| **Rate of BPD** | | NR | NR | N/A | NR |
| **Rate of IVH** | | 0 | 0 | N/A | 0 |
| **Rate of PVL/white matter injury** | | 0 | 0 | N/A | 0 |
| **Rate of Cerebral Palsy** | | NR | NR | N/A | NR |
| **Intervention** | | | | | |
| **Specific interventional concepts** | | CareToy | | | |
| **Description of intervention** | | - Family-centred, goal-directed program using a common baby gym (instrumented hanging toys; vision module, sensorized mat)<br>- Remote session planning according to infants' needs<br>- Phase 1: one week habituation to system<br>- Phase 2: three weeks customized training | | | |
| **Aim of intervention:**<br>Improvement of... | | - Visual and motor development<br>- Parent-child interaction and therefor reduce parenting stress | | | |
| **Age of participants at start of intervention (years)** | | 0-1 | | | |
| **Setting** | | Online/at home (reports are automatically sent to rehabilitative staff to customize next sessions) | | | |
| **Number of sessions** | | 28 | | | |
| **Frequency of sessions** | | Daily | | | |
| **Duration of program** | | 4 weeks | | | |
| **Comparator** | | | | | |
| **Type of comparator** | | SoC | | | |
| **Outcomes** | | **Assessment tool** | | | |
| **Outcomes relevant for our study** | | | | | |
| **Parent-child interaction** | after intervention (a.i.) | Parenting Stress Index, Short form | | | |
| **Outcomes *not* relevant for our study** | | | | | |
| **Caregiver mental health** | (a.i.) | Parenting Stress Index, Short form | | | |
| **Motor** | (a.i.) | InfantMotor Profile (IMP)<br>Alberta Infant Motor Scale (AIMS) | | | |
| **Vision** | (a.i.) | Teller Acuity Cards | | | |

| | | | | | |
|---|---|---|---|---|---|
| **Study ID** | | Resnick 1987 | | | |
| **DOI** | | NR | | | |
| **Title** | | **Developmental intervention for low birth weight infants: improved early development outcome** | | | |
| **Type of RCT** (parallel/multiple groups) | | Parallel group study | | | |
| **Additional study reports available** | | None identified | | | |
| **Study objective:**<br>To examine … | | - mental and physical development | | | |
| **Setting** (single-/multi-centric) | | single-centric | | | |
| **Country in which the study conducted** | | United States | | | |
| **Start date** | | 26/01/1979 | | | |
| **End date** | | 21/09/1981 | | | |
| **Funding source** | | Jessie Ball Dupont Foundation, National Foundation March of Dimes, Developmental Disability Council, State of Florida, Developmental Services Program, State of Florida, and Children's Medical Services, Department of Health and Rehabilitative Services, State of Florida | | | |
| **Trial registration number(s)** | | NR | | | |
| **Conflict of interest** | | NR | | | |
| **Inclusion criteria** | | - BW 500 - 1,800 g<br>- Admission to the University of Florida's Regional Neonatal Intensive Care Center at Shands Teaching Hospital, Gainesville<br>- between 26/01/1979 - 21/09/1981<br>- Survival of first 24h of life | | | |
| **Exclusion criteria** | | NR | | | |
| **preterms as…** | | Full population | | | |
| **gestational age (weeks)** | | NR | | | |
| **birth weight** | | 500-1800g | | | |
| **If multiple group study, define intervention 1, intervention 2, intervention 3** | | **IG 1** | Intervention | | |
| | | **IG 2** | control | | |
| | | **IG 3** | N/A | | |
| **Sociodemographic Characteristics:** | | **IG 1** | **IG 2** | **IG 3** | **total** |
| **sociodemographic characterstics of parents** | **education** | Maternal education (mean years) 11.9 | Maternal education (mean years) 12.1 | N/A | NR |
| | **income** | NR | NR | N/A | mean per capita income approx. $6,000 |
| | **employment** | NR | NR | N/A | NR |
| | **marital status** | NR | NR | N/A | unmarried: 45% |
| **Number of participants allocated** | | 124 | 11 | N/A | 255 |
| **baseline population characterstics** | | **IG 1** | **IG 2** | **IG 3** | **total** |

| | | | | |
|---|---|---|---|---|
| **No. of participants** | 107 | 114 | N/A | 221 |
| **Mean birthweight (SD) in gramme** | 1411 (275) | 1417 (308) | N/A | NR |
| **Mean gestational age (SD) in weeks** | 31.5 (1.1) | 31.6 (2.8) | N/A | NR |
| **No. of male/female** | 52/55 | 44/70 | N/A | 96/125 |
| **percentage of male/female (in %)** | 49/51 | 39/61 | N/A | 43/57 |
| **multiples** | NR | NR | N/A | NR |
| **Rate of BPD** | "and/or respiratory distress syndrome" 43% | "and/or respiratory distress syndrome" 45% | N/A | NR |
| **Rate of IVH** | 25% | 24% | N/A | NR |
| **Rate of PVL/white matter injury** | NR | NR | N/A | NR |
| **Rate of Cerebral Palsy** | NR | NR | N/A | NR |
| **Intervention** | | | | |
| **Specific interventional concepts** | None | | | |
| **Description of intervention** | - Interdisciplinary team including psychology, nursing, occupational therapy, and early childhood education<br>NICU intervention:<br>- Vestibular and visual stimulation<br>- Two 20-minute-sessions/day with 25 exercises aiming at visual, auditory, tactile, kinesthetic,and personal-social development<br>- Services for caregivers (crisis counceling, parenting education, communication skills training)<br>Home-based:<br>- Pediatric nurse supporting family by home-visits, clinic visits, and phone-calls<br>- Early childhood development specialist providing developmental care for infants and support for parents<br>- Curriculum with exercises for ages 1 month to 2 years | | | |
| **Aim of intervention:**<br>Improvement of... | In infants:<br>- personal-social development<br>- visual development<br>- auditory development<br>- language<br>- memory<br>- perceptual motor skills<br>- physical development<br>in caregivers:<br>- bonding process<br>- personal-social development<br>- knowledge about child health care services | | | |
| **Age of participants at start of intervention (years)** | 0-1 | | | |
| **Setting** | Started inpatient;<br>Out-patient;<br>Home-visits;<br>Telephone calls | | | |
| **Number of sessions** | NR | | | |
| **Frequency of sessions** | daily in the NICU;<br>twice-monthly home visits until the child was 2 years old | | | |
| **Duration of program** | Birth to 2 years CA | | | |
| **Comparator** | | | | |

| **Type of comparator** | | SoC |
|---|---|---|
| **Outcomes** | | **Assessment tool** |
| **Outcomes *not* relevant for our study** | | |
| **Cognition** | **12/24 months** | Bayley - MDI |
| **Motor** | **12/24 months** | Bayley - PDI |

| | |
|---|---|
| **Study ID** | Mazumder 2019 |
| **DOI** | 10.1016/S0140-6736(19)32223-8 |
| **Title** | **Effect of community-initiated kangaroo mother care on survival of infants with low birthweight: a randomised controlled trial** |
| **Type of RCT** (parallel/multiple groups) | Parallel group study |
| **Additional study reports available** | Study protocol;<br>Trial registry record without results |
| **Study objective:**<br>To examine … | - the effect on the mortality from enrolment to age 28 days, and from enrolment to age 180 days (ie, 6 months). |
| **Setting** (single-/multi-centric) | multi-centric |
| **Country in which the study conducted** | India |
| **Start date** | xx/04/1983 |
| **End date** | xx/03/1984 |
| **Funding source** | Research Council of Norway through its Centers of Excellence Scheme (223269);<br>University of Bergen through funding to the Centre for Intervention Science in Maternal and Child Health |
| **Trial registration number(s)** | NCT02653534<br>NCT02631343<br>CTRI/2016/02/006653 |
| **Conflict of interest** | None |
| **Inclusion criteria** | BW 1500–2250 g<br>- Screening within 3 days of delivery<br>Infants born at home:<br>- Enrollment until 72h after birth<br>Infants born in health facilities:<br>- no initiation of Kangaroo Mother Care in the facility<br>BW 1500–1800g:<br>- Referral for hospital care (instead of intervention)<br>- Invitation to participate if the families refused to take the baby to the hospital or if the baby was taken to hospital but was either not admitted or admitted and discharged before they became 72h old |
| **Exclusion criteria** | Infants…<br>- not weighed within 72 h of birth<br>- unable to feed<br>- had difficulty in breathing<br>- had less than normal movements<br>- had major congenital malformations<br>- kangaroo mother care initiated in hospital |

| | | | | | |
|---|---|---|---|---|---|
| | - mothers did not intend to stay in the study area for the next 6 months or did not consent to participate | | | | |
| **preterms as...** | subgroup | | | | |
| **gestational age (weeks)** | preterm (<37 weeks) | | | | |
| **birth weight** | 1500-2250g | | | | |
| **If multiple group study, define intervention 1, intervention 2, intervention 3** | **IG 1** | Intervention | | | |
| | **IG 2** | control | | | |
| | **IG 3** | N/A | | | |
| **Sociodemographic Characteristics:** | **IG 1** | | **IG 2** | **IG 3** | **total** |
| **sociodemographic characterstics of parents** — **education** | Mother's schooling years 6·0 (5·2)<br><br>Mother never been to school 1625 (36·3%) | | Mother's schooling years 6·2 (5·2)<br><br>Mother never been to school 1341 (34·2%) | N/A | NR |
| **income** | Family wealth quintile<br>Least poor 891 (19·9%)<br><br>Less poor 917 (20·5%)<br><br>Poor 891 (19·9%)<br><br>Very poor 904 (20·2%)<br><br>Most poor 871 (19·5%)<br><br>Family below poverty line 909 (20·3%) | | Family wealth quintile<br>Least poor 794 (20·2%)<br><br>Less poor 766 (19·5%)<br><br>Poor 783 (20·0%)<br><br>Very poor 768 (19·6%)<br><br>Most poor 811 (20·7%)<br><br>Family below poverty line 796 (20·3%) | N/A | NR |
| **employment** | Mother working outside of home 226 (5·0%) | | Mother working outside of home 223 (5·7%) | N/A | NR |
| **marital status** | NR | | NR | N/A | NR |

| Number of participants allocated | 4480 | 3922 | N/A | 8402 |
|---|---|---|---|---|
| **baseline population characterstics** | **IG 1** | **IG 2** | **IG 3** | **total** |
| **No. of participants** | 4480 | 3922 | N/A | 8402 |
| **Mean birthweight (SD) in gramme** | Preterm and SGA<br>857 (19·4)<br><br>Preterm and AGA<br>1726 (39·1)<br><br>Term and SGA<br>1386 (41·6) | Preterm and SGA<br>746 (19·4)<br><br>Preterm and AGA<br>1476 (38·3)<br><br>Term and SGA<br>1628 (42·3) | N/A | NR |
| **Mean gestational age (SD) in weeks** | 36·1 (1·8) | 36·1 (1·8) | N/A | NR |
| **No. of male/female** | 1907/2573 | 1741/2181 | N/A | 3648/4754 |
| **percentage of male/female (in %)** | 42.6/57.4 | 44.4/55.6 | N/A | 43.4/56.6 |
| **multiples** (twin pairs) | 72 (1,6%) | 63 (1.6%) | N/A | 135 |
| **Rate of BPD** | NR | NR | N/A | NR |
| **Rate of IVH** | NR | NR | N/A | NR |
| **Rate of PVL/white matter injury** | NR | NR | N/A | NR |
| **Rate of Cerebral Palsy** | NR | NR | N/A | NR |
| **Intervention** | | | | |
| **Specific interventional concepts** | Community-initated Kangaroo Mother Care | | | |
| **Description of intervention** | - Home visits<br>- Promotion of skin-to-skin care (SSC) by mother and other family members<br>- exclusive breastfeeding | | | |
| **Aim of intervention:**<br>Improvement of... | - the mortality of babies | | | |
| **Age of participants at start of intervention (years)** | At day of birth | | | |
| **Setting** | Home-visits | | | |
| **Number of sessions** | 9 | | | |
| **Frequency of sessions** | days 1–3, 5, 7, 10, 14, 21, and 28 of life | | | |
| **Duration of program** | ≤28 days of age | | | |
| **Comparator** | | | | |
| **Type of comparator** | SoC | | | |
| **Outcomes** | **Assessment tool** | | | |
| **Outcomes *not* relevant for our study** | | | | |
| **Mortality ≤180 days** | | | | |
| **Morbidity ≤28 days ≤90 days ≤180 days** | serious bacterial infection or localised infection;<br><br>diarrhoea, with or without symptoms of dysentery or dehydration;<br>pneumonia, or severe pneumonia;<br>hospitalisation and care seeking for these illnesses | | | |

| | | | | | |
|---|---|---|---|---|---|
| **Study ID** | Ma 2015 | | | | |
| **DOI** | 10.1016/j.braindev.2014.07.002. | | | | |
| **Title** | **Effect of early intervention on premature infants' general movements** | | | | |
| **Type of RCT** (parallel/multiple groups) | Parallel group study | | | | |
| **Additional study reports available** | None identified | | | | |
| **Study objective:**<br>To examine ... | - the characteristics of infants' general movements (GMs) and the effect of early intervention on their GMs. | | | | |
| **Setting** (single-/multi-centric) | NR | | | | |
| **Country in which the study conducted** | China | | | | |
| **Start date** | xx/09/2011 | | | | |
| **End date** | xx/12/2012 | | | | |
| **Funding source** | Supported by Science and Technology Star Program from Department of Science & Technology of Jinan in Shandong Province (20120140);<br>Science and Technology Program from Binzhou Medical College (BY2013KJ31);<br>Research Foundation for Outstanding Young Scientists in Shandong Province (BS2013YY011) | | | | |
| **Trial registration number(s)** | NR | | | | |
| **Conflict of interest** | NR | | | | |
| **Inclusion criteria** | - GA < 37 weeks<br>- Admission to NICU | | | | |
| **Exclusion criteria** | - Infection of central nervous system<br>- Genetic and metabolic disease<br>- Chromosomal disease<br>- Congenital abnormality<br>- Brain malformation and tumors of the central nervous system | | | | |
| **preterms as...** | Full population | | | | |
| **gestational age (weeks)** | preterm (<37 weeks) | | | | |
| **birth weight** | NR | | | | |
| **If multiple group study, define intervention 1, intervention 2, intervention 3** | **IG 1** | Intervention | | | |
| | **IG 2** | control | | | |
| | **IG 3** | N/A | | | |
| **Sociodemographic Characteristics:** | | **IG 1** | **IG 2** | **IG 3** | **total** |
| **sociodemographic charactersitics of parents** | **education** | NR | NR | N/A | NR |
| | **income** | NR | NR | N/A | NR |
| | **employment** | NR | NR | N/A | NR |
| | **marital status** | NR | NR | N/A | NR |
| **Number of participants allocated** | | 145 | 140 | N/A | 285 |
| **baseline population charactersitics** | | **IG 1** | **IG 2** | **IG 3** | **total** |
| **No. of participants** | | 145 | 140 | N/A | 285 |
| **Mean birthweight (SD) in gramme** | | 2640 (610) | 2520 (560) | N/A | NR |

| | | | | |
|---|---|---|---|---|
| **Mean gestational age (SD) in weeks** | 33.93 (2.61) | 33.49 (1.89) | N/A | NR |
| **No. of male/female** | 74/71 | 75/65 | N/A | 149/136 |
| **percentage of male/female (in %)** | 51.0/49.0 | 53.5/46.5 | N/A | 52.3/47.5 |
| **multiples** | NR | NR | N/A | NR |
| **Rate of BPD** | NR | NR | N/A | NR |
| **Rate of IVH** | Grade 1–2: 3.4%, Grade 3–4: 1.3% | Grade 1–2: 4.2%, Grade 3–4: 0% | N/A | 13 |
| **Rate of PVL/white matter injury** | 7.6% | 6.2% | N/A | 20 |
| **Rate of Cerebral Palsy** | NR | NR | N/A | NR |
| **Intervention** | | | | |
| **Specific interventional concepts** | - | | | |
| **Description of intervention** | Hospital intervention:<br>- auditory stimulation<br>- visual stimulation<br>- tactile stimulation<br>post-discharge:<br>- neurodevelopmental follow-up by professional doctor<br>- parental training in the intervention methods | | | |
| **Aim of intervention:** | - auditory stimulation<br>- visual stimulation<br>- tactile stimulation<br>- vestibular motion<br>- stimulation<br>- pediatric gymnastic<br>- hydrotherapy | | | |
| **Age of participants at start of intervention (years)** | 3 days | | | |
| **Setting** | Started inpatient;<br>Outpatient | | | |
| **Number of sessions** | NR | | | |
| **Frequency of sessions** | hospital itnervention: 1-2 a day<br>family intervention: once a week<br>intervention apllied by parents: once a day | | | |
| **Duration of program** | ≤ 54 weeks of GA | | | |
| **Comparator** | | | | |
| **Type of comparator** | SoC | | | |
| **Outcomes** | **Assessment tool** | | | |
| **Outcomes *not* relevant for our study** | | | | |
| **Motor 14 weeks** | Frequency of writhing and fidgety movements and comparison between different GA-groups and BW-Groups | | | |

| | | | | | |
|---|---|---|---|---|---|
| **Study ID** | Lecuona 2017 | | | | |
| **DOI** | 10.7196/SAMJ.2017.v107i11.12393 | | | | |
| **Title** | **Sensory integration intervention and the development of the premature infant: A controlled trial** | | | | |
| **Type of RCT** (parallel/multiple groups) | Parallel group study | | | | |
| **Additional study reports available** | None identified | | | | |
| **Study objective:**<br>To examine … | …the effect on the development of ELBW to VLBW premature infants from low socioeconomic settings. | | | | |
| **Setting** (single-/multi-centric) | single-centric | | | | |
| **Country in which the study conducted** | South Africa | | | | |
| **Start date** | NR | | | | |
| **End date** | NR | | | | |
| **Funding source** | South African Institute for Sensory Integration (SAISI) Research Committee of the School for Allied Health Professions of the University of the Free State (UFS) | | | | |
| **Trial registration number(s)** | NR | | | | |
| **Conflict of interest** | None | | | | |
| **Inclusion criteria** | - GA 26 - 36 weeks<br>- BW 750 - 1,499 g<br>- Medically stable<br>-<br>- Corrected age 4 - 10 months? | | | | |
| **Exclusion criteria** | - Previous occupational therapy or sensory integration intervention<br>- Additional conditions/neurological abnormalities | | | | |
| **preterms as…** | Full population | | | | |
| **gestational age (weeks)** | 26-36 | | | | |
| **birth weight** | 750-1499g | | | | |
| **If multiple group study, define intervention 1, intervention 2, intervention 3** | **IG 1** | Intervention | | | |
| | **IG 2** | control | | | |
| | **IG 3** | N/A | | | |
| **Sociodemographic Characteristics:** | **IG 1** | | **IG 2** | **IG 3** | **total** |
| **sociodemographic characterstics of parents** — **education** | Mother<br>50% Matric<br>50% Other<br><br>Father<br>58.3% Matric<br>41.7% Other | | Mother<br>58.3% Matric<br>41.7% Other<br><br>Father<br>75% Matric<br>25% Other | N/A | NR |
| **income** | NR | | NR | N/A | NR |
| **employment** | *Mother*<br>Unemployed<br>83.3% | | *Mother* | N/A | NR |

| | | | | |
|---|---|---|---|---|
| | Unqualified/casual 8.3%<br>Professional 8.3%<br><br>*Father*<br>Unemployed 0%<br>Unqualified/casual 75%<br>Professional 25% | Unemployed 58.3%<br>Unqualified/casual 0%<br>Professional 41.7%<br><br>*Father*<br>Unemployed 8.3%<br>Unqualified/casual 83.3%<br>Professional 8.3% | | |
| **marital status** | Married/living together 83.3%<br><br>Single 16.7% | Married/living together 58.3%<br><br>Single 41.7% | N/A | NR |
| **Number of participants allocated** | 12 | 12 | N/A | 24 |
| **baseline population characterstics** | **IG 1** | **IG 2** | **IG 3** | **total** |
| **No. of participants** | 12 | 12 | N/A | 24 |
| **Mean birthweight (SD) in gramme** | 1,098 g (SD not reported) | 1,204 g (SD not reported) | N/A | NR |
| **Mean gestational age (SD) in weeks** | 30.3 weeks (SD not reported) | 30.7 weeks (SD not reported) | N/A | NR |
| **No. of male/female** | 8/4 | 6/6 | N/A | 14/10 |
| **percentage of male/female (in %)** | 66.7/33.3 | 50/50 | N/A | 58/42 |
| **multiples** | NR | NR | N/A | NR |
| **Rate of BPD** | NR | NR | N/A | NR |
| **Rate of IVH** | NR | NR | N/A | NR |
| **Rate of PVL/white matter injury** | NR | NR | N/A | NR |
| **Rate of Cerebral Palsy** | NR | NR | N/A | NR |
| **Intervention** | | | | |
| **Specific interventional concepts** | Ayres Sensory Integration (ASI) | | | |
| **Description of intervention** | - weekly 45-minute ASI intervention sessions applied by occupational therapist<br>- two appropriate play/handling recommendations demonstrated to caregivers | | | |
| **Aim of intervention:**<br>Improvement of**...** | - understanding infants' expression and developmental progress.<br>- infants' sensory environment.<br>- sensory processing and self-regulation and/or their behavioural expression in terms of arousal, attention, affect and action. | | | |
| **Age of participants at start of intervention (years)** | 4-10 months corrected age | | | |
| **Setting** | NR | | | |
| **Number of sessions** | 10 | | | |
| **Frequency of sessions** | weekly | | | |
| **Duration of program** | 10 weeks | | | |

| **Comparator** | | | |
|---|---|---|---|
| **Type of comparator** | | SoC | |
| **Outcomes** | | **Assessment tool** | |
| **Outcomes *not* relevant for our study** | | | Comment |
| **Participation (language)** | **Before/after intervention** | Bayley-III (expressive/receptive communication) | Too early (<4y CA) |
| **Behavior** | | Bayley-III (social-emotional, adaptive) | Not externalizing/ internalizing |
| **Cognition** | | Bayley-III (cognitive scale) | |
| **Motor** | | Bayley-III (fine/gross motor) | |
| **Sensory processing** | | Infant/Toddler Sensory Profile;<br>Test of Sensory Functions in Infants | |

| | |
|---|---|
| **Study ID** | Landry 2006 |
| **DOI** | 10.1037/0012-1649.42.4.627 |
| **Title** | **Responsive parenting: establishing early foundations for social, communication, and independent problem-solving skills** |
| **Type of RCT** (parallel/multiple groups) | Parallel group study |
| **Additional study reports available** | None identified |
| **Study objective:**<br>To examine … | - the effect of maternal responsiveness on infant skills<br>- the effect of different aspects of responsiveness<br>- the effect in different BW-groups |
| **Setting** (single-/multi-centric) | multi-centric |
| **Country in which the study conducted** | United states |
| **Start date** | NR |
| **End date** | NR |
| **Funding source** | National Institutes of Health Grant HD36099 |
| **Trial registration number(s)** | NR |
| **Conflict of interest** | NR |
| **Inclusion criteria** | VLBW (< 1500g) OR born at term equivalent age |
| **Exclusion criteria** | NR |
| **preterms as...** | subgroup |
| **gestational age (weeks)** | NR |
| **birth weight** | VLBW (< 1500g) |

| | | |
|---|---|---|
| **If multiple group study, define intervention 1, intervention 2, intervention 3** | **IG 1** | Intervention |
| | **IG 2** | control |
| | **IG 3** | N/A |

| **Sociodemographic Characteristics:** | | **IG 1** | **IG 2** | **IG 3** | **total** |
|---|---|---|---|---|---|
| **sociodemographic characterstics of parents** | **education** | NR | NR | N/A | Mothers 12.6 years |
| | **income** | NR | NR | N/A | NR |
| | **employment** | NR | NR | N/A | NR |
| | **marital status** | One parent families: 62%<br><br>Two parent families: 38% | One parent families: 55%<br><br>Two parent families: 45% | N/A | NR |
| **Number of participants allocated** | | 133 | 131 | N/A | 264 |

| **baseline population characterstics** | **IG 1** | **IG 2** | **IG 3** | **total** |
|---|---|---|---|---|
| **No. of participants** | 133 | 131 | N/A | 164 |
| **Mean birthweight (SD) in gramme**<br>high risk VLBW–low risk VLBW–term | 833–1,256–3,385 | 918–1,412–3,332 | N/A | NR |
| **Mean gestational age (SD) in weeks**<br>high risk VLBW–low risk VLBW–term | 27–30–40 | 27–31–40 | N/A | NR |

| | | | | |
|---|---|---|---|---|
| **No. of male/female** | 60/73 | 68/63 | N/A | 128/136 |
| **percentage of male/female (in %)** | 45/55 | 52/48 | N/A | 48/52 |
| **multiples** | NR | NR | N/A | NR |
| **Rate of BPD** | NR | NR | N/A | NR |
| **Rate of IVH** | NR | NR | N/A | NR |
| **Rate of PVL/white matter injury** | NR | NR | N/A | NR |
| **Rate of Cerebral Palsy** | NR | NR | N/A | NR |
| **Intervention** | | | | |
| **Specific interventional concepts** | Playing And Learning Strategies (PALS) | | | |
| **Description of intervention** | - videotaped examples of problem solving activities<br>- caregiver teaching using videos of their own behavior<br>- emotional-affective support for parents | | | |
| **Aim of intervention:**<br>Improvement of... | - contingent responding<br>- support for infant foci of attention<br>- language input. | | | |
| **Age of participants at start of intervention** | 6-10 months | | | |
| **Setting** | Home-visits | | | |
| **Number of sessions** | 10 | | | |
| **Frequency of sessions** | Weekly | | | |
| **Duration of program** | 10 weeks | | | |
| **Comparator** | | | | |
| **Type of comparator** | Another type of early intervention | | | |

| | | | |
|---|---|---|---|
| **Outcomes** | | **Assessment tool** | Comment |
| **Relevant, but *not eligible* outcomes for meta-analysis** | | | |
| **Behavior** | **6-13 months** | Coding of videotapes | Missing data |
| **Parent-child interaction** | **6-13 months** | Coding of videotapes | Missing data |
| **Outcomes *not* relevant for our study** | | | |
| **Maternal behavior** | **6-13 months** | Coding of videotapes | |

| | | | | |
|---|---|---|---|---|
| **Study ID** | Gianni 2006 | | | |
| **DOI** | 10.1016/j.earlhumdev.2006.01.011 | | | |
| **Title** | **The effects of an early developmental mother-child intervention program on neurodevelopment outcome in very low birth weight infants: a pilot study** | | | |
| **Type of RCT** (parallel/multiple groups) | Parallel group study | | | |
| **Additional study reports available** | None identified | | | |
| **Study objective:**<br>To examine … | …the effect of promoting of a secure maternal-infant attachment and child's self-regulation on neurodevelopment. | | | |
| **Setting** (single-/multi-centric) | single-centric | | | |
| **Country in which the study conducted** | Italy | | | |
| **Start date** | xx/01/2001 | | | |
| **End date** | xx/08/2001 | | | |
| **Funding source** | NR | | | |
| **Trial registration number(s)** | NR | | | |
| **Conflict of interest** | NR | | | |
| **Inclusion criteria** | - BW <1250 g<br>- Singleton<br>- Infant are fed preterm formula | | | |
| **Exclusion criteria** | - Congenital diseases and/or chromosomal abnormalities<br>- Abnormal brain magnetic resonance imaging (MRI) at 40 CA<br>- Dead of infant during post-partum hospital stay | | | |
| **preterms as...** | Full population | | | |
| **gestational age (weeks)** | NR | | | |
| **birth weight** | <1250g | | | |
| **If multiple group study, define intervention 1, intervention 2, intervention 3** | **IG 1** | Intervention | | |
| | **IG 2** | control | | |
| | **IG 3** | N/A | | |
| **Sociodemographic Characteristics:** | **IG 1** | **IG 2** | **IG 3** | **total** |
| **sociodemographic characterstics of parents** – **education** | *Mother (%)*<br>low 5.5<br>medium 66.7<br>high 27.8 | *Mother (%)*<br>low 5.5<br>medium 77.8<br>high 16.7 | N/A | NR |
| **income** | NR | NR | N/A | NR |
| **employment** | NR | NR | N/A | NR |
| **marital status** | NR | NR | N/A | NR |
| **Number of participants allocated** | 18 | 18 | N/A | 36 |
| **baseline population characterstics** | **IG 1** | **IG 2** | **IG 3** | **total** |
| **No. of participants** | 18 | 18 | N/A | 36 |
| **Mean birthweight (SD) in gramme** | 892 (240) | 836 (162) | N/A | NR |
| **Mean gestational age (SD) in weeks** | 28.3 (2.8) | 27.5 (1.8) | N/A | NR |
| **No. of male/female** | 8/10 | 9/9 | N/A | 17/19 |
| **percentage of male/female (in %)** | 44.4/55.6 | 50/50 | N/A | 47.2/52.8 |
| **multiples** | NR | NR | N/A | NR |

| | | | | |
|---|---|---|---|---|
| **Rate of BPD** | NR | NR | N/A | NR |
| **Rate of IVH** | NR | NR | N/A | NR |
| **Rate of PVL/white matter injury** | NR | NR | N/A | NR |
| **Rate of Cerebral Palsy** | NR | NR | N/A | NR |
| **Intervention** | | | | |
| **Specific interventional concepts** | - | | | |
| **Description of intervention** | - Group meetings of 4-6 mother-infant dyads with a psychologist and a psychometrician<br>- Psychological support for mother to cope with preterm birth by a psychologist<br>- Promotion of perceptual and social-cognitive skills in PCI by a psychometrician | | | |
| **Aim of intervention:**<br>Improvement of**...** | - Mothers' mental health<br>- Secure attachment between infant and mother | | | |
| **Age of participants at start of intervention** | 3 months CA | | | |
| **Setting** | Outpatient | | | |
| **Number of sessions** | 18 | | | |
| **Frequency of sessions** | Twice/month | | | |
| **Duration of program** | 9 month | | | |
| **Comparator** | | | | |
| **Type of comparator** | SoC | | | |
| **Outcomes** | **Assessment tool** | | | |
| **Outcomes relevant for our study but *without* eligible assessment tool for meta-analysis** | | | | |
| **Executive functioning 3 years** | GMDS – Hand and eye coordination, performance, practical reasoning | | Not overall-score for EF | |
| **Outcomes *not* relevant for our study** | | | | |
| **Behavior 3 years** | Griffiths Mental Development Scale(GMDS) – personal social | | Not externalizing/internalizing | |
| **Participation (language) 3 years** | GMDS – hearing and speech | | Too early (<4y CA) | |
| **Motor 3 years** | GMDS - locomotor | | | |

| | |
|---|---|
| **Study ID** | Gaden 2022 |
| **DOI** | 10.1001/jamanetworkopen.2023.15750 |
| **Title** | **Short-term Music Therapy for Families With Preterm Infants: A Randomized Trial** |
| **Type of RCT** (parallel/multiple groups) | Multiple group study |
| **Additional study reports available** | Secondary journal pulication (e.g. follow-up study);<br>Study protocol;<br>Trial registry record without results;<br>Other |
| **Study objective:**<br>To examine … | - treatment fidelity of music therapy (MT) for premature infants and their parents; and to present the theoretical framework and intervention protocol for this resource-oriented MT approach<br>- the feasibility (acceptability, integrability, and safety) of the music therapy intervention<br>- short-term effects on the effect on parental well-being, mother-infant bonding, parental anxiety, and maternal depression<br>- the effect on bonding between primary caregivers and preterm infants.<br>- the effect on weight gain among preterm infants hospitalized in Neonatal Intensive Care Units<br>- the effect on language development |
| **Setting** (single-/multi-centric) | multi-centric |
| **Country in which the study conducted** | Argentina<br>Colombia<br>Israel<br>Norway<br>Poland |
| **Start date** | 25/08/201 |
| **End date** | 31/08/2022 |
| **Funding source** | Research Council of Norway (RCN, project number 273534), under the program High-quality and Reliable Diagnostics, Treatment and Rehabilitation (BEHANDLING);<br>Faculty of Fine Art, Music, and Design (KMD) at the University of Bergen, and the POLYFON Knowledge Cluster for Music Therapy. |
| **Trial registration number(s)** | NCT03564184 |
| **Conflict of interest** | None |
| **Inclusion criteria** | Infants:<br>- GA <35 weeks<br>- Hospitalization likely for a minimum of 2 weeks from inclusion<br>- Medically stable to start MT<br>Parents: |

| | | | | | |
|---|---|---|---|---|---|
| | - Written, site-specific informed consent<br>- Willingness to engage in at least 2 of 3 MT weekly sessions<br>- Sufficient understanding of the respective national language(s) to answer the questionnaires and participate in MT<br>- Capacity to complete the intervention and questionnaires; Living within reasonable commuting distance from the treating NICU | | | | |
| **Exclusion criteria** | - Dropout due to mental illness of parents or else | | | | |
| **preterms as...** | Full population | | | | |
| **gestational age (weeks)** | <35 weeks | subgroups:<br>a) <28<br>b) 28 to <32<br>c) 32 to <35 | | | |
| **birth weight** | NR | | | | |
| **If multiple group study, define intervention 1, intervention 2, intervention 3** | **IG 1** | Intervention (music therapy in NICU and post-discharge) | | | |
| | **IG 2** | Control (standard of care in NICU and post-discharge) | | | |
| | **IG 3** | N/A | | | |
| **Sociodemographic Characteristics:** | | **IG 1** | **IG 2** | **IG 3** | **total** |
| | **education** | NR | NR | N/A | Mother education, mean years (SD)<br>16.4 (2.7)<br><br>Father education, mean years (SD)<br>15.6 (3.2) |
| **sociodemographic characterstics of parents** | **income** | NR | NR | N/A | |
| | **Employment no. (%)** | NR | NR | N/A | **Mother**<br>Full-time- or self-employed<br>12 (67%)<br><br>Other<br>6 (33%)<br><br>**Father**<br>Full-time- or self-employed<br>17 (94%)<br><br>Other<br>1 (6%) |

| | | | | |
|---|---|---|---|---|
| **marital status no. (%)** | NR | NR | N/A | **Mother**<br>Married<br>13 (72%)<br><br>Living together but not married<br>5 (28%) |
| **Number of participants allocated** | NR | NR | N/A | NR |
| **baseline population characterstics** | **IG 1** | **IG 2** | **IG 3** | **total** |
| **No. of participants** | 52 | 50 | N/A | 102 |
| **Mean birthweight (SD) in gramme** | 1391.3 (420.9) | 1422.9 (428.2) | N/A | NR |
| **Mean gestational age (SD) in weeks** | 30.4 (2.6) | 30.9 (2.6) | N/A | NR |
| **No. of male/female** | 27/25 | 25/25 | N/A | NR |
| **percentage of male/female (in %)** | 52/48 | 50/50 | N/A | NR |
| **multiples** | NR | NR | N/A | NR |
| **Rate of BPD** | NR | NR | N/A | NR |
| **Rate of IVH** | NR | NR | N/A | NR |
| **Rate of PVL/white matter injury** | NR | NR | N/A | NR |
| **Rate of Cerebral Palsy** | NR | NR | N/A | NR |
| **Intervention** | | | | |
| **Specific interventional concepts** | - | | | |
| **Description of intervention** | NICU<br>- Three individual 30-minutes MT sessions per week (max. 27 sessions)<br>- parent-led, infant-directed singing; adapted to infants' postmenstrual age and their reaction<br>- parent education about reading infants' cues<br>- psychotherapeutic support of parents according to trauma-preventive models<br>post-discharge:<br>- individual 45-minutes MT sessions at home or in follow-up clinics<br>- parent-led, infant-directed singing adapted to infant maturity<br>- virtual MT sessions starting April 1, 2020 (due to COVID-19 pandemic) | | | |
| **Aim of intervention:**<br>Improvement of... | - mother-infant bonding<br>- parental mental health | | | |
| **Age of participants at start of intervention (years)** | NR | | | |
| **Setting** | NR | | | |
| **Number of sessions** | NR | | | |
| **Frequency of sessions** | NR | | | |
| **Duration of program** | NR | | | |
| **Comparator** | | | | |
| **Type of comparator** | Standard of Care | | | |
| **Outcomes** | **Assessment tool** | | | |
| **Outcomes relevant for our study** | | | | |
| **Rehospitalisation 12 months** | time from initial discharge until first rehospitalisation | | | |
| **Outcomes relevant for our study but *without* eligible assessment tool for meta-analysis** | | | | |

| | | | |
|---|---|---|---|
| **Parent-Child Interaction** | **6 months**<br>**12 months** | Postpartum Bonding Questionnaire | |
| **Outcomes *not* relevant for our study** | | | |
| **Parental Anxiety** | **At discharge**<br>**6/12 months** | Generalised Anxiety Disorder Assess-ment (GAD-7) | |
| **Parental Stress** | **6/12 months** | Parental Stress Scale | |
| **Cognition** | **24 months** | Bayley-III | |
| **Motor (gross/fine)** | **24 months** | Bayley-III | |
| **Participation (language)** | **6/12 months** | Ages and Stages Questionnaire 3 | Too early |
| | **24 months** | Bayley-III | |
| **Socioemotional Behavior** | **6/12 months** | Ages and Stages Questionnaire: Social-Emotional, second edition | |
| **Growth development** | **Birth,**<br>**Study enrolment,**<br>**At discharge** | Fenton fetal–infant growth chart | |

| | | | | |
|---|---|---|---|---|
| **Study ID** | Edraki 2015 | | | |
| **DOI** | NR | | | |
| **Title** | **Effect of home visit training program on growth and development of preterm infants: a double blind randomized controlled trial** | | | |
| **Type of RCT** (parallel/multiple groups) | Parallel group study | | | |
| **Additional study reports available** | Trial registry record without results | | | |
| **Study objective:**<br>To examine … | - the effect on growth and development within 6 months. | | | |
| **Setting** (single-/multi-centric) | multi-centric | | | |
| **Country in which the study conducted** | Iran | | | |
| **Start date** | xx/04/2010 | | | |
| **End date** | xx/05/2011 | | | |
| **Funding source** | Shiraz University of Medical<br>Sciences, Shiraz, Iran (ICR-87-4275) | | | |
| **Trial registration number(s)** | IRCT2014082013690N3 | | | |
| **Conflict of interest** | None | | | |
| **Inclusion criteria** | - GA <37 weeks<br>- BW <2500 g<br>- Being fed orally<br>- Living in Shiraz<br>- No participation in another program or intervention<br>- Parents not part of the medical staff<br>- Mother's age 18 years or above | | | |
| **Exclusion criteria** | - Brain disorders<br>- Congenital cardiovascular diseases, metabolic or endocrine disorders<br>- Known genetic and chromosomal abnormalities<br>- Previous history of preterm infants in their family | | | |
| **preterms as...** | Full population | | | |
| **gestational age (weeks)** | Preterm (<37 weeks) | | | |
| **birth weight** | LBW (< 2500g) | | | |
| **If multiple group study, define intervention 1, intervention 2, intervention 3** | **IG 1** | Intervention | | |
| | **IG 2** | control | | |
| | **IG 3** | N/A | | |
| **Sociodemographic Characteristics:** | **IG 1** | **IG 2** | **IG 3** | **total** |
| **education**<br>**sociodemographic characterstics of parents** | *Mother, n (%)*<br>High school and lower<br>14(48.3)<br><br>diploma and College degree<br>15(51.7) | *Mother, n (%)*<br>High school and lower<br>11(37.9)<br><br>diploma and College degree<br>18(62.1) | N/A | NR |

| | | | | |
|---|---|---|---|---|
| | *Father, n (%)*<br>High school and lower<br>12(41.4)<br><br>Diploma and College degree<br>17(58.6) | *Father, n (%)*<br>High school and lower<br>11(37.9)<br><br>Diploma and College degree<br>18(62.1) | | |
| **income** | NR | NR | N/A | NR |
| **employment** | NR | NR | N/A | NR |
| **marital status** | NR | NR | N/A | NR |
| **Number of participants allocated** | 30 | 30 | N/A | 60 |
| **baseline population characterstics** | **IG 1** | **IG 2** | **IG 3** | **total** |
| **No. of participants** | 30 | 30 | N/A | 60 |
| **Mean birthweight (SD) in gramme** | 1860.0 (560.35) | 2084.0 (298.45) | N/A | NR |
| **Mean gestational age (SD) in weeks** | 35.10 (2.79) | 35.60 (1.61) | N/A | NR |
| **No. of male/female** | 14/16 | 15/15 | N/A | 29/31 |
| **percentage of male/female (in %)** | 46.7/53.3 | 50/50 | N/A | 48.3/51.7 |
| **multiples** | NR | NR | N/A | NR |
| **Rate of BPD** | NR | NR | N/A | NR |
| **Rate of IVH** | NR | NR | N/A | NR |
| **Rate of PVL/white matter injury** | NR | NR | N/A | NR |
| **Rate of Cerebral Palsy** | NR | NR | N/A | NR |
| **Intervention** | | | | |
| **Specific interventional concepts** | - | | | |
| **Description of intervention** | - Parental training about caretaking of preterm babies<br>- Provided by a pediatric nurse | | | |
| **Aim of intervention:**<br>Improvement of**...** | - Growth and development | | | |
| **Age of participants at start of intervention (years)** | 0-1 | | | |
| **Setting** | Home-visits | | | |
| **Number of sessions** | 7 | | | |
| **Frequency of sessions** | first 3 visits: 1st day / 2nd day / 1 week post-discharge<br>following 4 visits: 1st, 2nd, 3rd, 6th months after birth | | | |
| **Duration of program** | 6 months | | | |
| **Comparator** | | | | |
| **Type of comparator** | SoC | | | |
| **Outcomes** | **Assessment tool** | | | |
| **Outcomes *not* relevant for our study** | | | | |
| **Growth** **6 months** | measurements | | | |
| **Neurodevelopment** **6 months** | Developmental indexes | | | |

| | | | | |
|---|---|---|---|---|
| **Study ID** | Dusing 2015 | | | |
| **DOI** | 10.1097/PEP.0000000000000161 | | | |
| **Title** | **Supporting Play Exploration and Early Development Intervention From NICU to Home: A Feasibility Study** | | | |
| **Type of RCT** (parallel/multiple groups) | Parallel group study | | | |
| **Additional study reports available** | None identified | | | |
| **Study objective:**<br>To examine … | - the feasibility and efficacy of a clinical trial of Supporting Play Exploration and Early Development Intervention (SPEEDI). | | | |
| **Setting** (single-/multi-centric) | single-centric | | | |
| **Country in which the study conducted** | United States | | | |
| **Start date** | NR | | | |
| **End date** | NR | | | |
| **Funding source** | grant from the Virginia Commonwealth University School of Allied Health Professions Promotion of Research Program | | | |
| **Trial registration number(s)** | NR | | | |
| **Conflict of interest** | None | | | |
| **Inclusion criteria** | - GA ≤ 34 weeks<br>- medically stable, off ventilator support, demonstrate thermoregulation by 35 weeks postmenstrual age<br>- Living within 30 minutes of the hospital | | | |
| **Exclusion criteria** | - Genetic syndrome<br>- Musculoskeletal deformity | | | |
| **preterms as...** | Full population | | | |
| **gestational age (weeks)** | ≤ 34 weeks | | | |
| **birth weight** | NR | | | |
| **If multiple group study, define intervention 1, intervention 2, intervention 3** | **IG 1** | Intervention | | |
| | **IG 2** | control | | |
| | **IG 3** | N/A | | |
| **Sociodemographic Characteristics:** | **IG 1** | **IG 2** | **IG 3** | **total** |
| **sociodemographic characterstics of parents** – **education** | Mothers with a HS degree or less 60% | Mothers with a HS degree or less 40% | N/A | NR |
| **income** | Mothers living in poverty 40% | Mothers living in poverty 40% | N/A | NR |
| **employment** | NR | NR | N/A | NR |
| **marital status** | NR | NR | N/A | NR |
| **Number of participants allocated** | 5 | 5 | N/A | 10 |
| **baseline population characterstics** | **IG 1** | **IG 2** | **IG 3** | **total** |
| **No. of participants** | 5 | 5 | N/A | 10 |
| **Median birthweight (IQR) in gramme** | 785 (690-1600) | 1375 (920-183) | N/A | NR |

| | | | | |
|---|---|---|---|---|
| **Median gestational age (IQE) in weeks** | 27 (25-30 | 31 (28-33) | N/A | NR |
| **No. of male/female** | 2/3 | 3/2 | N/A | 5/5 |
| **percentage of male/female (in %)** | 40/60 | 60/40 | N/A | 50/50 |
| **multiples** | 0 | 0 | N/A | 0 |
| **Rate of BPD** | NR | NR | N/A | NR |
| **Rate of IVH** | 1 | 0 | N/A | 1 |
| **Rate of PVL/white matter injury** | 0 | 0 | N/A | 0 |
| **Rate of Cerebral Palsy** | NR | NR | N/A | NR |

| | | |
|---|---|---|
| **Intervention** | | |
| **Specific interventional concepts** | SPEEDI | |
| **Description of intervention** | Phase 1 (NICU):<br>- 35 weeks PMA to 0 months CA<br>- By physical therapist<br>- varying degrees of positioning and interaction support to encourage the infant's self-directed movements<br>- parent coaching in said interventions<br>Phase 2 (post-discharge):<br>- parent coaching to read child's cues<br>- promoting development-appropriate play | |
| **Aim of intervention:**<br>Improvement of... | - parental support for child's development<br>- PCI | |
| **Age of participants at start of intervention** | 35 weeks post-menstrual age | |
| **Setting** | Phase 1: NICU<br>Phase 2: NR | |
| **Number of sessions** | 6 | |
| **Frequency of sessions** | every two weeks | |
| **Duration of program** | From 35 weeks' PMA until 3 months CA | |
| **Comparator** | | |
| **Type of comparator** | SoC | |
| **Outcomes** | **Assessment tool** | |
| **Outcomes *not* relevant for our study** | | |
| **Study feasibility** | | |
| **Cognition** **3 months** | Early Problem Solving Indicator (EPSI) oft he Individual Growth and DevelopmentIndicators (IGDI) | |
| **4 months** | Early Problem Solving Indicator (EPSI) oft he Individual Growth and DevelopmentIndicators (IGDI) | |
| **6 months** | Early Problem Solving Indicator (EPSI) oft he Individual Growth and DevelopmentIndicators (IGDI);<br>Bayley | |
| **Participation (language)** **6 months** | Bayley-III | Too early (<4y CA) |
| **Motor** **0 months** | Test of Infant Motor Performance (TIMP) | |
| **Motor** **3/4 months** | Test of Infant Motor Performance (TIMP);<br>(Pre-)Reaching behavior (video-based coding) | |
| **Motor** **6 months** | Test of Infant Motor Performance (TIMP);<br>(Pre-)Reaching behavior (video-based coding);<br>Bayley-III | |

| | | | | |
|---|---|---|---|---|
| **Study ID** | Cameron 2021 | | | |
| **DOI** | 10.1080/03004430.2020.1723571 | | | |
| **Title** | **Efficacy of the portage early intervention programme 'growing: Birth to three' for children born prematurely** | | | |
| **Type of RCT** (parallel/multiple groups) | Parallel group study | | | |
| **Additional study reports available** | None identified | | | |
| **Study objective:**<br>To examine ... | - the effects of a family-centred early intervention on cognitive and language development.<br>- the effect on parental well-beeing. | | | |
| **Setting** (single-/multi-centric) | single-centric | | | |
| **Country in which the study conducted** | Norway | | | |
| **Start date** | xx/10/2004 | | | |
| **End date** | xx/xx/2010 | | | |
| **Funding source** | NR | | | |
| **Trial registration number(s)** | NR | | | |
| **Conflict of interest** | None | | | |
| **Inclusion criteria** | - GA <30 weeks and/or BW <1500g<br>- born in the period 2004–2007 | | | |
| **Exclusion criteria** | need for comprehensive medical care | | | |
| **preterms as...** | Full population | | | |
| **gestational age (weeks)** | <30 weeks | | | |
| **birth weight** | VLBW (< 1500g) | | | |
| **If multiple group study, define intervention 1, intervention 2, intervention 3** | **IG 1** | Intervention | | |
| | **IG 2** | control | | |
| | **IG 3** | N/A | | |
| **Sociodemographic Characteristics:** | **IG 1** | **IG 2** | **IG 3** | **total** |
| **sociodemographic characterstics of parents** — **education** | 15.1 (1.8)y | 15.7 (2.0)y | N/A | Tertiary: >50% of mothers |
| **income** | NR | NR | N/A | NR |
| **employment** | NR | NR | N/A | NR |
| **marital status** | NR | NR | N/A | NR |
| **Number of participants allocated** | 19 | 19 | N/A | 38 |
| **baseline population characterstics** | **IG 1** | **IG 2** | **IG 3** | **total** |
| **No. of participants** | 19 | 17 | N/A | 36 |
| **Mean birthweight (SD) in gramme** | 1114.4 (265.9) | 1145.6 (250.1) | N/A | NR |
| **Mean gestational age (SD) in weeks** | 28.5 (2.5) | 29.4 (2.3) | N/A | NR |
| **No. of male/female** | 7/12 | 7/10 | N/A | 14/22 |
| **percentage of male/female (in %)** | 36.8/63.2 | 41.2/58.8 | N/A | 38.9/61,1 |

| | | | | |
|---|---|---|---|---|
| **multiples** | 11 singleton | 10 singletons | N/A | 21 singletons |
| **Rate of BPD** | NR | NR | N/A | NR |
| **Rate of IVH** | NR | NR | N/A | NR |
| **Rate of PVL/white matter injury** | NR | NR | N/A | NR |
| **Rate of Cerebral Palsy** | NR | NR | N/A | NR |
| **Intervention** | | | | |
| **Specific interventional concepts** | Growing: Brith to three (based on Portage) | | | |
| **Description of intervention** | - family-centered<br>- promotion of parent–child interactions<br>- consideration of cultural background<br>- decision-making based on recorded observations and discussion between parents and advisors | | | |
| **Aim of intervention:**<br>Improvement of**…** | - sensitivity and responsivity regarding the child | | | |
| **Age of participants at start of intervention** | 4 months CA | | | |
| **Setting** | Home-visits | | | |
| **Number of sessions** | 30 | | | |
| **Frequency of sessions** | Until 12 months CA: 1/months<br>Thereafter: 1/every other month | | | |
| **Duration of program** | 3 years | | | |
| **Comparator** | | | | |
| **Type of comparator** | SoC | | | |
| **Outcomes** | **Assessment tool** | | | |
| **Outcomes *not* relevant for our study** | | | | |
| **Participation (language)** **36 months** | Reynell Developmental Language Scales (RDLS) | | | Too early (<4y CA) |
| **Global Infant Development** **4/9/18/36 months** | Bayley-II (overall score of cognition, motor, and language scores) | | | |

| | |
|---|---|
| **Study ID** | Butera 2022 |
| **DOI** | 0.3390/jpm12122024 |
| **Title** | **Effect of a NICU to Home Physical Therapy Intervention on White Matter Trajectories, Motor Skills, and Problem-Solving Skills of Infants Born Very Preterm: A Case Series** |
| **Type of RCT** (parallel/multiple groups) | Parallel group study / Case study |
| **Additional study reports available** | Study protocol;<br>Trial registry record without results |
| **Study objective:**<br>To examine … | - infant's general movement to determine the risk of cerebral palsy.<br>- the neuromotor, motor and cognitive development.<br>- the correlation of changes in white matter microstructure and macrostructure with cognitive, motor and motor-based problem solving. |
| **Setting** (single-/multi-centric) | multi-centric |
| **Country in which the study conducted** | United States |
| **Start date** | 06/02/2019 |
| **End date** | 01/07/2025 (estimated) |
| **Funding source** | Eunice Kennedy Shriver National Institute of Child Health and Human Development, grant number 1R01HD093624;<br>Virginia Commonwealth University Clinical and Translational Science Award (CTSA), grant number UL1TR002649;<br>University of Southern California Provost's Undergrad Research Fellowship |
| **Trial registration number(s)** | NCT03518736 |
| **Conflict of interest** | None |
| **Inclusion criteria** | - GA <29 weeks;<br>- between 35 and 42 weeks of gestation when the baseline developmental assessment was completed<br>- Admission in one of the three participating NICUs<br>- Living within 100 miles (protocol: 60 miles) of the participating hospitals<br>- Medically stable, off ventilator support |
| **Exclusion criteria** | - Non-English-speaking families<br>- Genetic abnormality at the time of enrolment |
| **preterms as…** | Full population |
| **gestational age (weeks)** | <29 weeks |
| **birth weight** | VLBW (< 1500g) |

| | | |
|---|---|---|
| **If multiple group study, define intervention 1, intervention 2, intervention 3** | **IG 1** | Intervention |
| | **IG 2** | control |
| | **IG 3** | N/A |

| **Sociodemographic Characteristics:** | **IG 1** | **IG 2** | **IG 3** | **total** |
|---|---|---|---|---|

| | | | | |
|---|---|---|---|---|
| sociodemographic characterstics of parents — education | Infant 1: high school (mother & father)<br><br>Infant 2: <college (mother), professional school Degree (father)<br><br>Infant 6: NR | Infant 3: professional school degree (mother), bachelor's degree (father)<br><br>Infant 4: college (mother), some high school (father)<br><br>Infant 5: NR | N/A | NR |
| income | NR | NR | N/A | NR |
| employment | NR | NR | N/A | NR |
| marital status | NR | NR | N/A | NR |
| **Number of participants allocated** | 3<br>(Children 1, 2, and 6) | 3<br>(Children 3, 4, and 5) | N/A | 6 |
| **baseline population characterstics** | **IG 1** | **IG 2** | **IG 3** | **total** |
| **No. of participants** | 3 | 3 | N/A | 6 |
| **Mean birthweight (SD) in gramme** | 819,3 (SD not reported) | 1000 (SD not reported) | N/A | 909.7 (SD not reported) |
| **Mean gestational age (SD) in weeks** | 25.7 (SD not reported) | 27 (SD not reported) | N/A | 26.3 (SD not reported) |
| **No. of male/female** | 1 / 2 | 1 / 2 | N/A | 2/4 |
| **percentage of male/female (in %)** | 33.3/66.7 | 33.3/66.7 | N/A | 33.3/66.7 |
| **multiples** | 0 | 0 | N/A | 0 |
| **Rate of BPD** | NR | NR | N/A | NR |
| **Rate of IVH** | 33.3% | 0 | N/A | 16.7% |
| **Rate of PVL/white matter injury** | 2x minimal<br>1x normal | 1x minimal<br>2x normal | N/A | 3x minimal<br>3x normal |
| **Rate of Cerebral Palsy** | NR | NR | N/A | NR |
| **Intervention** | | | | |
| **Specific interventional concepts** | SPEEDI2 | | | |
| **Description of intervention** | - targeted environmental enrichment<br>- sensory-motor learning opportunities<br>- caregiver training in motor, cognitive, and social skill practice for their infant, e.g. with play-activies | | | |
| **Aim of intervention:**<br>Improvement of... | - caregiver knowledge about how to support their childs development | | | |
| **Age of participants at start of intervention (years)** | early group: in the NICU after baseline assessment (35-42 weeks)<br>late group: after assessment visit 2 (15 weeks post-baseline = 50-57 weeks GA) | | | |
| **Setting** | started inpatient in NICU (N/A for Occupational therapy); outpatient (hospital, office etc.);<br><br>home-visits | | | |

| | | |
|---|---|---|
| **Number of sessions** | | 10 |
| **Frequency of sessions** | | phase 1: 5 sessions over 3 weeks<br>phase 2: 5 sessions over three months |
| **Duration of program** | | 15 weeks |
| **Comparator** | | |
| **Type of comparator** | | SoC |
| **Outcomes** | | **Assessment tool** |
| **Outcomes *not* relevant for our study** | | |
| **Cognition** | **15 weeks** | Bayley-III;<br>Assessment of Problem Solving in Play (APSP-4) |
| **Cognition** | **30 weeks** | Bayley-III;<br>Assessment of Problem Solving in Play (APSP-4) |
| **Cognition** | **12 months** | Bayley-III;<br>Assessment of Problem Solving in Play (APSP-4) |
| **Motor** | **Baseline** | Prechtl's General Movement Assessment (GMA);<br>Test of Infant Motor Performance (TIMP) |
| **Motor** | **15 weeks** | Prechtl's General Movement Assessment (GMA);<br>Test of Infant Motor Performance (TIMP);<br>Bayley-III |
| **Motor** | **30 weeks** | Test of Infant Motor Performance (TIMP);<br>Bayley-III |
| **Motor** | **12 months** | Bayley-III |
| **changes in white matter microstructure and macrostructure** | **Baseline**<br>**15 weeks**<br>**30 weeks**<br>**12 months** | Qualitative and descriptive changes for developmental measures, FD, FC and FDC, and brain injury classification |

| | | | | |
|---|---|---|---|---|
| **Study ID** | Beckwith 1988 | | | |
| **DOI** | 10.1080/00332747.1988.11024398 | | | |
| **Title** | **Intervention with disadvantaged parents of sick preterm infants** | | | |
| **Type of RCT** (parallel/multiple groups) | Parallel group study | | | |
| **Additional study reports available** | | | | |
| **Study objective:**<br>To examine ... | - the effectiveness of supportive home visitor services to parents of infants who were biologically and socially at risk | | | |
| **Setting** (single-/multi-centric) | NR | | | |
| **Country in which the study conducted** | United States | | | |
| **Start date** | NR | | | |
| **End date** | NR | | | |
| **Funding source** | Center of Prevention Research, Division of Prevention and Special Mental Health Programs, National Institute for Mental Health, Grant No. MH36902 | | | |
| **Trial registration number(s)** | NR | | | |
| **Conflict of interest** | NR | | | |
| **Inclusion criteria** | Infants:<br>- GA ≤35 weeks<br>- BW ≤2000g<br>- more than 3 days in NICU following birth<br>Parents:<br>- English speaking<br>- no more than high school education and/or neither of the parents to be working at more than unskilled or semi-skilled job | | | |
| **Exclusion criteria** | NR | | | |
| **preterms as...** | Full population | | | |
| **gestational age (weeks)** | ≤35 | | | |
| **birth weight** | ≤2000g | | | |
| **If multiple group study, define intervention 1, intervention 2, intervention 3** | **IG 1** | Intervention | | |
| | **IG 2** | control | | |
| | **IG 3** | N/A | | |
| **Sociodemographic Characteristics:** | **IG 1** | **IG 2** | **IG 3** | **total** |
| **sociodemographic characterstics of parents** — **education** | NR | NR | N/A | NR |
| **income** | NR | NR | N/A | 50% on welfare or support by family members |
| **employment** | NR | NR | N/A | NR |
| **marital status** | NR | NR | N/A | 39% single mothers |
| **Number of participants allocated** | 37 | 55 | N/A | 92 |
| **baseline population characterstics** | **IG 1** | **IG 2** | **IG 3** | **total** |
| **No. of participants** | NR | NR | N/A | NR |
| **Mean birthweight (SD) in gramme** | NR | NR | N/A | <1500 |

| | | | | |
|---|---|---|---|---|
| **Mean gestational age (SD) in weeks** | NR | NR | N/A | <31 |
| **No. of male/female** | NR | NR | N/A | NR |
| **percentage of male/female (in %)** | NR | NR | N/A | NR |
| **multiples** | NR | NR | N/A | NR |
| **Rate of BPD** | NR | NR | N/A | NR |
| **Rate of IVH** | NR | NR | N/A | NR |
| **Rate of PVL/white matter injury** | NR | NR | N/A | NR |
| **Rate of Cerebral Palsy** | NR | NR | N/A | NR |
| **Intervention** | | | | |
| **Specific interventional concepts** | program developed by team of UCLA preterm infant study | | | |
| **Description of intervention** | - regular meetings of professional home visitor and parents<br>- focus on mother-child interaction<br>- individualized and parent-directed; based on trusting and supportive relationship<br>- mediation between family and community resources; concrete support (clothes, equipment, ...) | | | |
| **Aim of intervention:**<br>Improvement of**...** | - parental self-confidenc and competence<br>- parents' observational skills in relation to their infants | | | |
| **Age of participants at start of intervention (years)** | 0-1 | | | |
| **Setting** | started inpatient in NICU (N/A for Occupational therapy);<br><br>home-visits | | | |
| **Number of sessions** | NR | | | |
| **Frequency of sessions** | NR | | | |
| **Duration of program** | until the infants reached 13 months of age | | | |
| **Comparator** | | | | |
| **Type of comparator** | NR | | | |
| **Outcomes** | **Assessment tool** | | | |
| **Outcomes relevant for our study but *not* eligible for meta-analysis** | | | | Comment |
| **Parent-Child Interaction** **1/9 month** | Pre-coded checklist | | | Missing data |
| **Behavior** **13/20 months** | NR | | | Missing data |
| **Outcomes *not* relevant for our study** | | | | |
| **Cognition** **13/20 months** | Balyley - MDI | | | |

| | | | | |
|---|---|---|---|---|
| **Study ID** | Barrera 1986 | | | |
| **DOI** | 10.2307/1130634 | | | |
| **Title** | **Early home intervention with low-birth-weight infants and their parents** | | | |
| **Type of RCT** (parallel/multiple groups) | Multiple group study | | | |
| **Additional study reports available** | Secondary journal pulication (e.g. follow-up study) | | | |
| **Study objective:**<br>To examine ... | - the effect of parent-child-interaction at 4, 16, and 24 months.<br>- the social competence and emotional, cognitive, and behavioural development at 4, 8, 12, and 16 months CA.<br>- motor development at 4, 8, 12, and 16 months an 5 years CA. | | | |
| **Setting** (single-/multi-centric) | multi-centric | | | |
| **Country in which the study conducted** | Canada | | | |
| **Start date** | xx/04/1979 | | | |
| **End date** | NR | | | |
| **Funding source** | Ministry of Community and Social Services, Province of Ontario, Canada;<br><br>5years follow-up: Grant No. 8-41854 from the Ministry of Community and Social Services, through the Ontario Mental Health Foundation, April 1983 to March 1985. | | | |
| **Trial registration number(s)** | NR | | | |
| **Conflict of interest** | NR | | | |
| **Inclusion criteria** | - GA < 37 weeks<br>- BW < 2,000g<br>- Discharge from hospital at least 2 weeks prior to enrollment in the project<br>- Infant's prognosis for survival after discharge judged to be good by a pediatrician<br>- Family living within the geographic region funded for services | | | |
| **Exclusion criteria** | NR | | | |
| **preterms as...** | Full population | | | |
| **gestational age (weeks)** | preterm (<37 weeks) | | | |
| **birth weight** | < 2000g | | | |
| **If multiple group study, define intervention 1, intervention 2, intervention 3** | **IG 1** | Developmental Programming Intervention (DPI) | | |
| | **IG 2** | Parent-Infant Intervention (PII) | | |
| | **IG 3** | Control | | |
| **Sociodemographic Characteristics:** | **IG 1** | **IG 2** | **IG 3** | **total** |
| **sociodemographic characterstics of parents** **education** | Mother (years)<br>12 | Mother (years)<br>12 | Mother (years)<br>12 | NR |

| | | | | |
|---|---|---|---|---|
| | Father (years)<br>12 | Father (years)<br>13 | Father (years)<br>12 | |
| **income** | % in socioeconomic classes III + IV<br>47 | % in socioeconomic classes III + IV<br>50 | % in socioeconomic classes III + IV<br>62 | NR |
| **employment** | NR | NR | N/A | NR |
| **marital status** | Married (%)<br>88 | Married (%)<br>100 | Married (%)<br>95 | NR |
| **Number of participants allocated** | 16 | 22 | 21 | 59 |
| **baseline population characterstics** | **IG 1** | **IG 2** | **IG 3** | **total** |
| **No. of participants** | NR | NR | NR | NR |
| **Mean birthweight (SD) in gramme** | 1637 (SD not reported) | 1670 (SD not reported) | 1723 (SD not reported) | NR |
| **Mean gestational age (SD) in weeks** | 33 (SD not reported) | 33 (SD not reported) | 33 (SD not reported) | NR |
| **No. of male/female** | NR | NR | NR | NR |
| **percentage of male/female (in %)** | 53/47 | 55/45 | 57/43 | NR |
| **multiples** | NR | NR | NR | NR |
| **Rate of BPD** | NR | NR | NR | NR |
| **Rate of IVH** | NR | NR | NR | NR |
| **Rate of PVL/white matter injury** | NR | NR | NR | NR |
| **Rate of Cerebral Palsy** | NR | NR | NR | NR |
| **Intervention** | | | | |
| **Specific interventional concepts** | - | | | |
| **Description of intervention** | - Focuse on parent-infant dyad;<br>parent applied intervention after training by therapist (infant development and understanding their infant's behavior)<br>- Facilitated access to social community resources<br>- DPI: specific curricular activities to support overall child development (motor function, cognition, communication, behavioral skills,...)<br>- PII: individual plan adjusted to specific PCI-situations (e.g. feeding); adjustment of parents' behavior to child's needs | | | |
| **Aim of intervention:**<br>Improvement of**...** | - DPI: the child's functional development<br>- PII: parents' sensitivity and responsivity to infant's needs | | | |
| **Age of participants at start of intervention** | 4 months CA | | | |
| **Setting** | Home visits | | | |
| **Number of sessions** | 12-28 (mean=23) | | | |
| **Frequency of sessions** | weekly for the first 4 months, every other week thereafter, once a month during the last quarter of the year. | | | |
| **Duration of program** | 12 months | | | |
| **Comparator** | | | | |
| **Type of comparator** | No treatment | | | |
| **Outcomes** | **Assessment tool** | | | |
| **Outcomes relevant for our study but *not* eligible for meta-analysis** | | | | |
| **Parent-Child Interaction** **4/8/12/16 months** | Behavioral coding of video-taped freeplay | | Missing data | |

| | | | |
|---|---|---|---|
| **Parent-Child Interaction** | **5 years** | Parenting Stress Index | No data |
| **Executive Functioning** | **5 years** | Visual Motor Integration | Missing data/no overall EF score |
| **Home environment** | **4/8/12/16/28 months<br>5 years** | HOME inventory | Missing data |
| **Outcomes *not* relevant for our study** | | | |
| **Overall Child Development** | **4/8/12/16 months** | Minnesota Child Development Inventory<br><br>Vineland Social Maturity Scale | |
| **Overall Child Development** | **5 years** | McCarthy Scales of Children's Abilities | |
| **Infant security** | **4/8/12/16 months** | Flint Infant security Scale | |
| **Child temperament** | **4/8/12/16 months** | Infant and Toddler Temperament Questionnaire | |
| **Child temperament** | **5 years** | Behavioral Style Questionnaire | |
| **Cognition** | **4/8/12/16 months** | Bayley Scale (mental development index) | |
| **Cognition** | **5 years** | Peabody Individual Achievement Test | |
| **Motor** | **4/8/12/16 months** | Bayley Scale (psychomotor development index) | |

| Abbreviations | |
|---|---|
| **AGA** | Appropriate for Gestational Age |
| **BPD** | Bronchopulmonary Disease |
| **BW** | Birth Weight |
| **CA** | Corrected Age |
| **CG** | Controll Group |
| **ELBW** | Extremely-Low-Birth-Weight (< 1000g) |
| **EF** | Executive Functioning |
| **GA** | Gestational Age |
| **GMDS** | Griffiths Mental Development Scale |
| **IG** | Intervention Group |
| **IVH** | Intraventricular Haemorrhage |
| **LBW** | Low Birth Weight |
| **NICU** | Neonatal Intensive Care Unit |
| **PCI** | Parent-Child Interaction |
| **PMA** | Post-menstrual age |
| **PVL** | Periventricular Leukomalacia |
| **RCT** | Randomized Controlled Trial |
| **SD** | Standard Deviation |
| **SGA** | Small for Gestational Age |
| **VLBW** | Very-Low-Birth-Weight (< 1500g) |
