## Supplementary material for "Effects of interdisciplinary early developmental intervention programs on behavior, executive functioning and participation in children born preterm: A systematic review with meta-analysis": S1 - RoB and characteristics of included studies

| | |
|---|---|
| Wrong intervention | 145 |
| Wrong study design | 81 |
| Conference abstract | 51 |
| <6 interventions post-discharge | 52 |
| Wrong timepoint of intervention | 46 |
| Wrong patient population | 23 |
| No involvement of professionals | 15 |
| Author request | 24 |
| Completed, not yet published | 6 |
| Ongoing | 14 |

| Title | Authors | Published Year | Journal | Volume | Issue | Pages | DOI | Reason for exclusion |
|---|---|---|---|---|---|---|---|---|
| **Randomised controlled trial of PREMM: early somatosensory stimulation (massage) in preterm infants** | | 2015 | Developmental Medicine & Child Neurology | 57 | | 94-95 | 10.1111/dmcn.34_12886 | Wrong intervention |
| **Neonatal and parental predictors of executive function in very preterm children** | Aarnoudse-Moens et al. | 2013 | Acta Paediatrica | 102 | 3 | 282-286 | 10.1111/apa.12101 | Wrong intervention |
| **A randomised controlled trial of an adaptive working memory training intervention in very preterm children: The IMPRINT study** | Actrn | 2012 | | | | | | Wrong intervention |

| | | | | | | | | |
|---|---|---|---|---|---|---|---|---|
| **Effect of massage on behavioural responses of preterm infants in an educational hospital in Iran** | Baniasadi et al. | 2019 | Journal of Reproductive and Infant Psychology | 37 | 3 | 302-310 | 10.1080/02646838.2019.1578866 | Wrong intervention |
| **Stockholm preterm interaction-based intervention (SPIBI) - study protocol for an RCT of a 12-month parallel-group post-discharge program for extremely preterm infants and their parents** | Baraldi et al. | 2020 | BMC Pediatr | 20 | 1 | 49 | 10.1186/s12887-020-1934-4 | Wrong intervention |
| **Effects of early crawling training on the motor development of very premature infants** | Barbu-Roth et al. | 2022 | Developmental medicine and child neurology | 64 | SUPPL 3 | 37 | 10.1111/dmcn.15214 | Wrong intervention |
| **Functions of the CNS in early discharge and home followup of very low birthweight infants** | Brooten et al. | 1991 | Clin Nurse Spec | 5 | 4 | 196-201 | 10.1097/00002800-199100540-00006 | Wrong intervention |
| **A randomized clinical trial of early hospital discharge and home follow-up of very-low-birth-weight infants** | Brooten et al. | 1986 | N Engl J Med | 315 | 15 | 934-9 | 10.1056/NEJM198610093151505 | Wrong intervention |
| **A randomized clinical trial of early hospital discharge and home follow-up of very-low-birth-weight infants** | Brooten et al. | 1987 | NLN publications | | 21?2194 | 95?106 | | Wrong intervention |
| **Physiotherapy intervention to improve motor co-ordination in ELBW preschool children: a randomised controlled trial** | Brown et al. | 2012 | Developmental medicine and child neurology | 54 | | 67 | 10.1111/j.1469-8749.2012.04289.x | Wrong intervention |

| | | | | | | | | |
|---|---|---|---|---|---|---|---|---|
| **A randomised controlled trial of group-based physiotherapy intervention for non-disabled ELBW children: motor and postural outcomes** | Brown et al. | 2016 | Journal of paediatrics and child health | 52 | | 87 | 10.1111/jpc.13194 | Wrong intervention |
| **Effects of an early family intervention on children's memory: The mediating effects of cortisol levels** | Bugental et al. | 2010 | Mind, Brain, and Education | 4 | 4 | 159-170 | 10.1111/j.1751-228X.2010.01095.x | Wrong intervention |
| **The effects of an early physical therapy intervention for very preterm, very low birth weight infants: a randomized controlled clinical trial** | Cameron et al. | 2005 | Pediatr | 17 | 2 | 107-19 | 10.1097/01.pep.0000163073.50852.58 | Wrong intervention |
| **Earlier discharge with community-based intervention for low birth weight infants: a randomized trial** | Casiro et al. | 1993 | Pediatrics | 92 | 1 | 128-34 | | Wrong intervention |
| **Assisted exercise improves bone strength in very low birthweight infants by bone quantitative ultrasound** | Chen et al. | 2010 | J Paediatr Child Health | 46 | 11 | 653-9 | 10.1111/j.1440-1754.2010.01822.x | Wrong intervention |
| **The effectiveness of Parent-Centered supportive Intervention on parenting sense of competence in parent of premature infants** | ChiCtr | 2022 | trial registry | | | | | Wrong intervention |
| **A Family Centered Intervention Study on Breastfeeding Support for Premature Infants** | ChiCtr | 2023 | trial registry | | | | | Wrong intervention |
| **Effect of daily physical exercise on bone health in premature babies** | Ctri | 2016 | trial registry | | | | | Wrong intervention |

| | | | | | | | | |
|---|---|---|---|---|---|---|---|---|
| **To see the Effect of movement therapy on premature babies can help to assess the development using general movement assessment - A pre post experimental study** | Ctri | 2020 | | | | | | Wrong intervention |
| **Impact of nurse led telephonic support on home care for mothers of premature babies after discharge from hospital** | Ctri | 2024 | trial registry | | | | | Wrong intervention |
| **To Compare the Effects of Preterm Infant Oral Motor Intervention versus Preterm Infant Oral Motor Intervention with Massage Therapy on Social-Emotional Development in Preterm Infants: A Randomized Controlled Trial Study...56th All India Occupational Thera** | Dandavate, et al. | 2019 | Indian Journal of Occupational Therapy (Wolters Kluwer India Pvt Ltd) | 51 | 2 | 63-63 | | Wrong intervention |
| **Stable Preterm Infants Gain More Weight and Sleep Less after Five Days of Massage Therapy** | Dieter et al. | 2003 | Journal of Pediatric Psychology | 28 | 6 | 403-411 | https://dx.doi.org/10.1093/jpepsy/jsg030 | Wrong intervention |
| **Promoting academic resilience in preterm infants with adaptive computerized training - a multi-center randomised controlled trial** | Drks | 2015 | | | | | | Wrong intervention |
| **Effects of Martial Arts on Neurocognition in Children Born Preterm** | Drks | 2019 | | | | | | Wrong intervention |
| **"The effects of family-centered physiotherapy on the cognitive and motor performance in premature** | Elbasan et al. | 2017 | Infant behav | 49 | | 214-219 | 10.1016/j.infbeh.2017.09.007 | Wrong intervention |

| | | | | | | | | |
|---|---|---|---|---|---|---|---|---|
| **Control of reaching movements in 6-year-old prematurely born children with motor problems -- an intervention study** | Eliasson et al. | 2003 | Advances in Physiotherapy | 5 | 1 | 33-48 | 10.1080/14038190310005780 | Wrong intervention |
| **Mother-very preterm infant relationship quality: RCT of Baby Triple P** | Evans et al. | 2017 | Journal of Child and Family Studies | 26 | 1 | 284-295 | 10.1007/s10826-016-0555-x | Wrong intervention |
| **Sunflower oil versus no oil moderate pressure massage leads to greater increases in weight in preterm neonates who are low birth weight** | Fallah et al. | 2013 | Early Hum Dev | 89 | 9 | 769-72 | 10.1016/j.earlhumdev.2013.06.002 | Wrong intervention |
| **Feasibility of a Preventive Parenting Intervention for Very Preterm Children at 18 Months Corrected Age: A Randomized Pilot Trial** | Flierman et al. | 2016 | J Pediatr | 176 | | 79-85.e1 | 10.1016/j.jpeds.2016.05.071 | Wrong intervention |
| **Efficacy of a Neuro-Developmental Treatment program to improve motor control in infants born prematurely** | Girolami, et al. | 1994 | Pediatric Physical Therapy | 6 | 4 | 175-184 | 10.1097/00001577-199406040-00002 | Wrong intervention |
| **Baby CareLink: using the Internet and telemedicine to improve care for high-risk infants** | Gray et al. | 2000 | Pediatrics | 106 | 6 | 1318-1324 | 10.1542/peds.106.6.1318 | Wrong intervention |
| **ezPreemie study protocol: a randomised controlled factorial trial testing web-based parent training and coaching with parents of children born very preterm** | Greene et al. | 2022 | BMJ Open | 12 | 6 | e063706 | 10.1136/bmjopen-2022-063706 | Wrong intervention |

| | | | | | | | | |
|---|---|---|---|---|---|---|---|---|
| **Computerized working memory training has positive long-term effect in very low birthweight preschool children** | Grunewaldt et al. | 2016 | Developmental Medicine & Child Neurology | 58 | 2 | 195-201 | 10.1111/dmcn.12841 | Wrong intervention |
| **Comparison of video and in-hospital consultations during early in-home care for premature infants and their families: A randomised trial** | Hägi-Pedersen et al. | 2022 | Journal of Telemedicine & Telecare | 28 | 1 | 24-36 | 10.1177/1357633X20913411 | Wrong intervention |
| **Multicentre randomised study of the effect and experience of an early inhome programme (PreHomeCare) for preterm infants using video consultation and smartphone applications compared with inhospital consultations: protocol of the PreHomeCare study** | Hagi-Pedersen et al. | 2017 | BMJ Open | 7 | 3 | e013024 | 10.1136/bmjopen-2016-013024 | Wrong intervention |
| **Effect and Experience of PreHomeCare of Preterm Infants Using Telecommunication and Smartphone Application** | Nct | 2015 | trial registry | | | | | Wrong intervention |
| **Tactile/kinesthetic stimulation (TKS) increases tibial speed of sound and urinary osteocalcin (U-MidOC and unOC) in premature infants (29-32weeks PMA)** | Haley et al. | 2012 | Bone | 51 | 4 | 661-6 | 10.1016/j.bone.2012.07.016 | Wrong intervention |
| **Daily physical exercise stimulates growth in premature infants: a randomized controlled trial** | Hassanein et al. | 2002 | Pediatric research | LB 2?10 | | | | Wrong intervention |

| | | | | | | | | |
|---|---|---|---|---|---|---|---|---|
| **Impact of massage therapy on motor outcomes in very low-birthweight infants: randomized controlled pilot** | Ho et al. | 2010 | Pediatr Int | 52 | 3 | 378-85 | 10.1111/j.1442-200X.2009.02964.x | Wrong intervention |
| **Could audiovisual training be used to improve cognition in extremely low birth weight children?** | Huotilainen et al. | 2011 | Acta Paediatr | 100 | 11 | 1489-94 | 10.1111/j.1651-2227.2011.02345.x | Wrong intervention |
| **Influence of tagrop along with conventional exercises (developmental supportive care) on motor and behaviour pattern among preterm infants** | Iasvariya Vijayalakshmie et al. | 2018 | Biomedicine (india) | 38 | 3 | 390?395 | | Wrong intervention |
| **Effect of Early Physical Therapy Treatment on Improvement of GMFM Score in Different Type of Cerebral Palsy Patients in Selected Urban Community Ahmedabad, Gujarat** | Indravadan et al. | 2017 | Indian Journal of Physiotherapy & Occupational Therapy | 11 | 3 | 43-45 | 10.5958/0973-5674.2017.00070.3 | Wrong intervention |
| **The effect of physical activity on Physiological and Behavioral Responses, Motor performance and Neuro-mascular Development** | Irct201405208315N | 2014 | trial registry | | | | | Wrong intervention |
| **The effect of massage of preterm neonates on anxiety of mothers** | Irct2016012918945N | 2016 | trial registry | | | | | Wrong intervention |
| **The effect of massage on physical development, reflex state and motor development in preterm neonates** | Irct2017012318945N | 2017 | trial registry | | | | | Wrong intervention |
| **Study Effect of Massage Therapy With or Without Physical Exercises on the weight gain Premature Infants** | Irct20170520034039N | 2018 | trial registry | | | | | Wrong intervention |

| | | | | | | | | |
|---|---|---|---|---|---|---|---|---|
| **The effect of peer support through social networks on the stress and caring ability of mothers' premature infants after discharge** | Irct20230719058841N | 2023 | trial registry | | | | | Wrong intervention |
| **Music is the Key: The impact of therapeutic listening on the development of children born preterm** | Isrctn | 2016 | | | | | | Wrong intervention |
| **Premature Infant Oral Motor Intervention (PIOMI) with and without Massage Therapy on Social Emotional Development in Preterm Infants** | Jaywant et al. | 2020 | Indian Journal of Occupational Therapy | 52 | 3 | 95-100 | 10.4103/ijoth.ijoth_13_20 | Wrong intervention |
| **Investigating the brain structural connectome following working memory training in children born extremely preterm or extremely low birth weight** | Kelly et al. | 2021 | J Neurosci Res | 99 | 10 | 2340-2350 | 10.1002/jnr.24818 | Wrong intervention |
| **Maternal coping strategies and emotional distress: results of an early intervention program for low birth weight young children** | Klebanov et al. | 2001 | Dev Psychol | 37 | 5 | 654-67 | | Wrong intervention |
| **Early intervention for children at risk of visual processing dysfunctions from 1 year of age: a randomized controlled trial protocol** | Kooiker et al. | 2020 | Trials | 21 | 1 | Jan 14 | 10.1186/s13063-019-3936-9 | Wrong intervention |
| **Assisted exercise and bone strength in preterm infants** | Litmanovitz et al. | 2007 | Calcif Tissue Int | 80 | 1 | 39-43 | 10.1007/s00223-006-0149-5 | Wrong intervention |
| **Early physical activity intervention prevents decrease of bone strength in very low birth weight infants** | Litmanovitz et al. | 2003 | Pediatrics | 112 | 1 Pt 1 | 15. Sep | 10.1542/peds.112.1.15 | Wrong intervention |

| | | | | | | | | |
|---|---|---|---|---|---|---|---|---|
| **A feasibility study of a novel early participation-focused physiotherapy intervention for preterm infants in a regional Australian context** | Mobbs et al. | 2020 | Developmental medicine and child neurology | 62 | SUPPL 3 | 86 | 10.1111/dmcn.14662 | Wrong intervention |
| **Impact of early and consistent occupational therapy with preterm infants' oral feedings** | Mohr et al. | 2004 | Pediatric research | 56 | 4 | 672 | | Wrong intervention |
| **Examining the effectiveness of body massage on physical status of premature neonates and their mothers' psychological status** | Mokaberian et al. | 2022 | Early Child Development and Care | 192 | 14 | 2311-2325 | 10.1080/03004430.2021.2006194 | Wrong intervention |
| **Can a daily program of massage and physical exercise affect bone mineralization? A randomized controlled trial in premature infants** | Moustafa et al. | 2003 | Pediatric research | 53 | | 84 | | Wrong intervention |
| **Effect of physical activity on bone mineralization in premature infants** | Moyer-Mileur et al. | 1995 | Journal of pediatrics | 127 | 4 | 620?625 | 10.1016/s0022-3476(95)70127-3 | Wrong intervention |
| **Mechanical-tactile stimulation increases tibial bone strength in preterm infants** | Moyer-Mileur et al. | 2011 | Journal of bone and mineral research | 26 | | | | Wrong intervention |
| **Daily physical activity program increases bone mineralization and growth in preterm very low birth weight infants** | Moyer-Mileur et al. | 2000 | Pediatrics | 106 | 5 | 1088?109 | 10.1542/peds.106.5.1088 | Wrong intervention |
| **Brief reaching training with "sticky mittens" in preterm infants: Randomized controlled trial** | Nascimento et al. | 2019 | Hum Mov Sci | 63 | | 138-147 | 10.1016/j.humov.2018.11.015 | Wrong intervention |

| | | | | | | | | |
|---|---|---|---|---|---|---|---|---|
| **Randomized controlled trial on effectiveness of mHealth (mobile/smartphone) based Preterm Home Care Program on developmental outcomes of preterms: Study protocol** | Nayak et al. | 2019 | J Adv Nurs | 75 | 2 | 452-460 | 10.1111/jan.13879 | Wrong intervention |
| **The Norwegian Physical Therapy Study in Preterm Infants** | Nct | 2010 | trial registry | | | | | Wrong intervention |
| **Helping Our Premature Infants ON to Better Motor Skills (HOP-ON)** | Nct | 2011 | | | | | | Wrong intervention |
| **Early Intervention in Very Preterm Children** | Nct | 2011 | trial registry | | | | | Wrong intervention |
| **Computerized Working Memory Training in Very-low-birth-weight Children at Preschool Age** | Nct | 2012 | | | | | | Wrong intervention |
| **Initial Efficacy Study of Supporting Play, Exploration, & Early Development Intervention** | Nct | 2014 | trial registry | | | | | Wrong intervention |
| **Brain Training in Preterm Children at Risk for Inattention, Hyperactivity, and Executive Function Impairment** | Nct | 2015 | | | | | | Wrong intervention |
| **GPS (Giving Parents Support): parent Navigation After NICU Discharge** | Nct | 2015 | trial registry | | | | | Wrong intervention |
| **Effect of Assisted Exercise on Musculoskeletal System and Growth in Preterm Infants** | Nct | 2016 | trial registry | | | | | Wrong intervention |
| **Early Physical Therapy Intervention in Preterm Infants** | Nct | 2017 | trial registry | | | | | Wrong intervention |
| **Use of a Tummy Time Intervention and Parent Education in Infants Born Preterm** | Nct | 2018 | trial registry | | | | | Wrong intervention |

| | | | | | | | | |
|---|---|---|---|---|---|---|---|---|
| **Whole-body Vibration in Spastic Hemiplegic Cerebral Palsy** | Nct | 2019 | trial registry | | | | | Wrong intervention |
| **Effectiveness of Family Collaborative Physiotherapy Programs With High-risk Infants** | Nct | 2019 | trial registry | | | | | Wrong intervention |
| **Physical Therapy to Prevent Osteopenia in Preterm Infants** | Nct | 2020 | trial registry | | | | | Wrong intervention |
| **Evidence for exercise-induced bone formation in premature infants** | Nemet et al. | 2002 | Int J Sports Med | 23 | 2 | 82-5 | 10.1055/s-2002-20134 | Wrong intervention |
| **Maternal holding of preterm infants during the early weeks after birth and dyad interaction at six months** | Neu et al. | 2010 | J Obstet Gynecol Neonatal Nurs | 39 | 4 | 401-14 | 10.1111/j.1552-6909.2010.01152.x | Wrong intervention |
| **Influence of holding practice on preterm infant development** | Neu, et al. | 2013 | MCN Am J Matern Child Nurs | 38 | 3 | 136-43 | 10.1097/NMC.0b013e31827ca68c | Wrong intervention |
| **Pilot study on the feasibility of a novel computerized cognitive training program to improve executive function in very preterm children** | Nl, Omon | 2013 | | | | | | Wrong intervention |
| **Blik Vooruit: visuele screening en revalidatie bij te vroeg geboren kinderen** | Ntr | 2018 | trial registry | | | | | Wrong intervention |
| **Study protocol: an early intervention program to improve motor outcome in preterm infants: a randomized controlled trial and a qualitative study of physiotherapy performance and parental experiences** | Oberg et al. | 2012 | BMC Pediatr | 12 |  | 15 | 10.1186/1471-2431-12-15 | Wrong intervention |

| | | | | | | | | |
|---|---|---|---|---|---|---|---|---|
| **Early Physiotherapy Intervention Program for Preterm Infants and Parents: A Randomized, Single-Blind Clinical Trial** | Ochandorena-Acha et al. | 2022 | Children (Basel) | 9 | 6 | 15 | 10.3390/children 9060895 | Wrong intervention |
| **Early intervention physical therapy for an infant born preterm, extremely low birth weight with inraventricular hemorrhage** | Phelps, L. K.; Ceele, C. | 2012 | Pediatric Physical Therapy | 24 | 4 | 379-379 | | Wrong intervention |
| **Early physical therapy effects on the high-risk infant: a randomized controlled trial** | Piper et al. | 1986 | Pediatrics | 78 | 2 | 216-24 | | Wrong intervention |
| **General Movements in preterm infants undergoing craniosacral therapy: a randomised controlled pilot-trial** | Raith et al. | 2016 | BMC Altern Med | 16 | | 12 | 10.1186/s12906-016-0984-5 | Wrong intervention |
| **Six-year follow-up of early physiotherapy intervention in very low birth weight** | Rothberg et al. | 1991 | Pediatrics | 88 | 3 | 547-52 | | Wrong intervention |
| **Factors that predict which preterm infants benefit most from massage therapy** | Scafidi et al. | 1993 | J Dev Behav Pediatr | 14 | 3 | 176-80 | | Wrong intervention |
| **Daily Exercise Program in very low Birth Weight Preterm Infants...Sezer EY, Erdem E, Gunes T. The effect of daily exercise program on bone mineral density and cortisol level in preterm infants with very low birth weight: A randomized controlled trial. Jou** | Shaw, Subhash Chandra | 2020 | Journal of Pediatric Nursing | 51 | | 108-108 | 10.1016/j.pedn.2019.10.001 | Wrong intervention |
| **The effect of a short bout of practice on reaching behavior in late preterm infants at the onset of reaching: a randomized controlled trial** | Soares Dde et al. | 2013 | Res Dev Disabil | 34 | 12 | 4546-58 | 10.1016/j.ridd.2013.09.028 | Wrong intervention |

| | | | | | | | | | |
|---|---|---|---|---|---|---|---|---|---|
| **Parent education programme on stimulation of language development in preterm infants at risk of brain damage** | Soberon et al. | 2019 | Revista de Logopedia, Foniatria y Audiologia | 39 | 1 | 32-40 | | 10.1016/j.rlfa.2018.06.003 | Wrong intervention |
| **Physical activity intervention improved the number and functionality of endothelial progenitor cells in low birth weight children** | Souza et al. | 2020 | Nutrition, Metabolism & Cardiovascular Diseases | 30 | 1 | 60-70 | | 10.1016/j.numecd.2019.08.011 | Wrong intervention |
| **Efficacy of reflex locomotion to prevent osteopenia in preterm infants** | Torro Ferrero et al. | 2018 | Pediatric critical care medicine | 19 | 6 | | 139 | | Wrong intervention |
| **Effect of physical therapy in the promotion of bone mineralization in preterm infants. A randomized clinical trial** | Torro et al. | 2020 | Developmental medicine and child neurology | 62 | SUPPL 4 | 21?22 | | 10.1111/dmcn.14688 | Wrong intervention |
| **Efficacy of reflex locomotion to prevent osteopenia in infants born preterm** | Torro-Ferrero et al. | 2018 | Developmental medicine and child neurology | 60 | | | 35 | 10.1111/dmcn.13790 | Wrong intervention |
| **Early Parent-Administered Physical Therapy for Preterm Infants: A Randomized Controlled Trial** | Ustad et al. | 2016 | Pediatrics | 138 | 2 | | 8 | 10.1542/peds.2016-0271 | Wrong intervention |

| | | | | | | | | | |
|---|---|---|---|---|---|---|---|---|---|
| **General movements in a group of infants born preterm who have participated in an early intervention study** | Ustad et al. | 2018 | Developmental medicine and child neurology | | 60 | | 58 | 10.1111/dmcn.13790 | Wrong intervention |
| **Physical therapy reduces bone resorption and increases bone formation in preterm infants** | Vignochi et al. | 2012 | Am J Perinatol | | 29 | 8 | 573-8 | 10.1055/s-0032-1310520 | Wrong intervention |
| **Interventional physical and occupational therapy services and motor coordination among low birth weight infants** | Watkins, Stephanie Elaine | 2012 | | Ph.D. | | | 193 p-193 p | | Wrong intervention |
| **Interventional physical and occupational therapy services and motor coordination among low birth weight infants.DP - 2013** | Watkins, Stephanie Elaine | 2013 | | | 74 | 5-B(E) | No Pagination Specified | | Wrong intervention |
| **Impact of physiotherapy on neuromotor development of premature newborns** | Xavier Coutinho et al. | 2014 | Fisioterapia em Movimento | | 27 | 3 | 413-420 | 10.1590/0103-5150.027.003.AO12 | Wrong intervention |
| **Effects of Vestibular Stimulation through Positioning in Hammocks on the development of the premature baby compared to Motor Physiotherapy** | z8tzjf, R. B. R. | 2022 | trial registry | | | | | | Wrong intervention |
| **Exploring Effect of Postdischarge Developmental Support Program on Preterm Infant Neurodevelopment and BDNF Gene DNA Methylation** | Zhang et al. | 2023 | Adv Neonat Care | | 23 | 2 | E50-E58 | 10.1097/ANC.0000000000001046 | Wrong intervention |
| **Analysis on the Application Effect of Abdominal Acupoint Massage on Feeding Intolerance in Premature Infants** | Zhu et al. | 2021 | J | | 2021 | | 2883597 | https://dx.doi.org/10.1155/2021/2883597 | Wrong intervention |

| | | | | | | | | | |
|---|---|---|---|---|---|---|---|---|---|
| **Therapeutic Listening for Preterm Children with Sensory Dysregulation, Attention and Cognitive Problems.** | Slevin et al. | 2020 | Ir Med J | 113 | | 1 | 4 | | Wrong intervention |
| **Best Start Trial: early intervention physiotherapy to improve motor outcomes in infants at high risk of cererbal palsy or motor delay** | Actrn | 2019 | trial registry | | | | | | Wrong intervention |
| **Treadmill training in moderate risk preterm infants promotes stepping quality--results of a small randomised controlled trial** | Angulo-Barroso et al. | 2013 | Res Dev Disabil | 34 | 11 | | 3629-38 | 10.1016/j.ridd.2013.07.037 | Wrong intervention |
| **Treadmill Training in Infants At-Risk for Neuromotor Delay** | Actrn | 2012 | trial registry | | | | | | Wrong intervention |
| **Body composition and neuromotor development in the year after NICU discharge in premature infants** | Cooper et al. | 2020 | Pediatr Res | 88 | 3 | | 459-465 | 10.1038/s41390-020-0756-2 | Wrong intervention |
| **A study to assess abnormal thinking and behavioral patterns using package among very low birth weight,early born** | Ctri | 2018 | trial registry | | | | | | Wrong intervention |
| **Role of Early Physiotherapy in improving motor development of Preterm and low-birth weight infants** | Ctri | 2018 | trial registry | | | | | | Wrong intervention |
| **Benefits of physiotherapy techniques and movement therapy on breathing and overall development of premature** | Ctri | 2020 | trial registry | | | | | | Wrong intervention |
| **Developing and testing a Structured Early Physiotherapy Intervention for Babies Born Moderate to Late Preterm to Promote their Neuromotor Development** | Ctri | 2023 | trial registry | | | | | | Wrong intervention |

| | | | | | | | | |
|---|---|---|---|---|---|---|---|---|
| **Promotion of motor development and movement quality in preterm infants through the early application of "Movement Imitation Therapy for Preterm Babies (MIT-PB)" in the first three months of life** | Drks | 2021 | trial registry | | | | | Wrong intervention |
| **Improvement of executive functions after the application of a neuropsychological intervention program (PEFEN) in pre-term children** | Garcia-Bermudez, et al. | 2019 | Children and Youth Services Review | 98 | | 328-336 | 10.1016/j.childyou | Wrong intervention |
| **Movement training advances the emergence of reaching in infants born at less than 33 weeks of gestational age: a randomized clinical trial** | Heathcock et al. | 2008 | Phys Ther | 88 | 3 | 310-22 | 10.2522/ptj.20070145 | Wrong intervention |
| **Exploring objects with feet advances movement in infants born preterm: a randomized controlled trial** | Heathcock, J. C.; Galloway, J. C. | 2009 | Phys Ther | 89 | 10 | 1027-38 | 10.2522/ptj.20080278 | Wrong intervention |
| **Development Training in Babies Born Preterm** | Nct | 2005 | trial registry | | | | | Wrong intervention |
| **Goal-directed training to the level of trunk control and reaching in preterm** | hyn, R. B. R. | 2018 | trial registry | | | | | Wrong intervention |
| **Improving the quality of parental interaction with very low birth weight children: A longitudinal study using a mediated learning experience model.DP - Win, 1991** | Klein, Pnina S. | 1991 | Infant Mental Health Journal | 12 | 4 | 321-337 | 10.1002/1097-0355%28199124%2912:4%3C321::AID-IMHJ2280120406 | Wrong intervention |

| | | | | | | | |
|---|---|---|---|---|---|---|---|
| **A Comparitive Study of Early Intervention Programme vs Home Intervention Programme in Preterm Infants** | Mann et al. | 2012 | Indian Journal of Physiotherapy & Occupational Therapy | 6 | 3 | 167-171 | Wrong intervention |
| **Language and Motor Skills of Preterm Infants in Pre-school Age: diagnosis and Early Intervention** | Nct | 2011 | trial registry | | | | Wrong intervention |
| **Effectiveness of Vojta Therapy in Motor Development of Preterm Children** | Nct | 2013 | | | | | Wrong intervention |
| **Infant Aquatics Neurodevelopment Premature Infants** | Nct | 2013 | trial registry | | | | Wrong intervention |
| **Early Behavioral Intervention for Preterm Infants** | Nct | 2015 | trial registry | | | | Wrong intervention |
| **Randomized Controlled Trial of Early Intensive Leg Exercise to Improve Walking in Children With Diplegia** | Nct | 2018 | trial registry | | | | Wrong intervention |
| **Home-based Visual-motor Training Program on Kindergarteners** | Nct | 2022 | trial registry | | | | Wrong intervention |
| **Neonatal Neurobehavioral And Motor Behavior In Ultra Early Physical Therapy Intervention** | Nct | 2022 | trial registry | | | | Wrong intervention |
| **Investigation of the Effectiveness of Environmental Enrichment-Based Intervention in Preterm Infants** | Nct | 2022 | trial registry | | | | Wrong intervention |
| **Effectiveness of Home Based Early Intervention of Extremely Premature Infant by Parent** | Nct | 2022 | trial registry | | | | Wrong intervention |

| | | | | | | | | |
|---|---|---|---|---|---|---|---|---|
| **Early Intervention Based on Neonatal Crawling in Very Premature Infants at Risk For Neurodevelopmental Disorder** | Nct | 2023 | trial registry | | | | | Wrong intervention |
| **The Effect of Physical Therapy Intervention on Motor Performance in Bhutanese Preterm Infants** | Nct | 2023 | trial registry | | | | | Wrong intervention |
| **Cuevas Medek Exercises on Balance and Postural Control in Children With Spastic Cerebral Palsy** | Nct | 2023 | trial registry | | | | | Wrong intervention |
| **Examining the Effect of Occupational Therapy-Based Parent Coaching on Feeding Problems in Children With a Preterm Birth** | Nct | 2024 | trial registry | | | | | Wrong intervention |
| **Power Move: a randomized controlled pilot study on a computerized motor intervention program to improve motor function in very preterm children at 5 years of age** | Nl, Omon | 2020 | | | | | | Wrong intervention |
| **Power Move: a randomized waitlist-controlled study on a computerized motor intervention program to improve motor function in very preterm children at five years of ageâ€™** | Nl | 2019 | trial registry | | | | | Wrong intervention |
| **Multicentre prospective randomised single-blind controlled study protocol of the effect of an additional parent-administered sensorimotor stimulation on neurological development of preterm infants: Primebrain** | Pelc, K.; Daniel, I.; Wenderickx, B.; Dan, B. | 2017 | BMJ Open | 7 | 12 | e018084 | 10.1136/bmjopen-2017-018084 | Wrong intervention |
| **Primebrain Stimulation** | Nct | 2014 | trial registry | | | | | Wrong intervention |

| | | | | | | | | |
|---|---|---|---|---|---|---|---|---|
| **Training and Manual Behavior in Premature Infants** | rjwrx, R. B. R. | 2013 | | | | | | Wrong intervention |
| **Effects of Prone Positioning on Head Control in Preterm Infants: randomized and Controlled Clinical Trial Protocol** | Santos Sampaio et al. | 2023 | International journal of environmental research and public health | 20 3 | | | 10.3390/ijerph20032375 | Wrong intervention |
| **Belly time in the development of premature newborns** | nwkr, R. B. R. | 2022 | trial registry | | | | | Wrong intervention |
| **Effects of home guidance provided by parents on the functionality of infants at biological risk** | xrzjs, R. B. R. | 2020 | trial registry | | | | | Wrong intervention |
| **The profile of executive function in very preterm children at 4 to 12 years** | Aarnoudse-Moens et al. | 2012 | Developmental Medicine & Child Neurology | 54 | 3 | 247-253 | 10.1111/j.1469-8749.2011.04150.x | Wrong study design |
| **The Newborn Individualized Developmental Care and Assessment Program (NIDCAP) as a Model for Clinical Music Therapy Interventions with Premature Infants** | Abromeit, Deanna Hanson | 2003 | Music Therapy Perspectives | 21 | 2 | 60-68 | 10.1093/mtp/21.2.60 | Wrong study design |

| | | | | | | | | |
|---|---|---|---|---|---|---|---|---|
| **Influence of in-home nursing care on the weight of the early discharged preterm newborn** | Alvarez Miro et al. | 2014 | Anales de pediatria (Barcelona, Spain : 2003) | 81 | 6 | 352?359 | 10.1016/j.anpedi.2013.10.024 | Wrong study design |
| **Effectiveness of a longitudinal psychosocial intervention to strengthen mother-child interactions: The role of biological and contextual moderators** | Alves et al. | 2022 | Children and Youth Services Review | 133 | | | 10.1016/j.childyouth.2021.106333 | Wrong study design |
| **Early intervention promotes intellectual development of premature infants: a preliminary report. Early Intervention of Premature Infants Cooperative Research Group** | Bao et al. | 1999 | Chin Med J | 112 | 6 | 520-3 | | Wrong study design |
| **Motor organization in very low birth weight infants during caregiving: effects of a developmental intervention** | Becker et al. | 1999 | J Dev Behav Pediatr | 20 | 5 | 344-54 | 10.1097/00004703-199910000-00009 | Wrong study design |
| **Implementation and effectiveness of a home-based early intervention program for blind infants and preschoolers.DP - May-Jun 1998** | Beelmann et al. | 1998 | Research in Developmental Disabilities | 19 | 3 | 225-244 | 10.1016/S0891-4222%2898%2900004-3 | Wrong study design |
| **Study on low birthweight infants** | Black, J. | 1991 | Nebraska Nurse | 24 | 1 | 30-33 | | Wrong study design |
| **Alleviating the impact of stress and trauma in the neonatal unit and beyond** | Cantle, Alison | 2013 | Infant Observation | 16 | 3 | 257-269 | 10.1080/13698036.2013.852723 | Wrong study design |

| | | | | | | | | |
|---|---|---|---|---|---|---|---|---|
| **The role of the Occupational Therapist with children with neuropsychomotor development delay** | Carramaschi de Souza, Ariana; de Souza Fazio Marino, Milena | 2013 | Cadernos de Terapia Ocupacional da UFSCar | 21 | 1 | 149-153 | 10.4322/cto.2013.019 | Wrong study design |
| **An efficacy study of occupational therapy with high-risk neonates.DP - Aug 1988** | Case-Smith, Jane | 1988 | American Journal of Occupational Therapy | 42 | 8 | 499-506 | 10.5014/ajot.42.8.499 | Wrong study design |
| **Effects of parent-child interaction program on neurodevelopment of premature infants after discharge** | ChiCtr | 2023 | | | | | | Wrong study design |
| **Pre-term birth and the mother-infant relationship. Intervention programmes** | Costabile, Angela; Veltri, Rosa | 2003 | Eta Evolutiva | 75 | 1 | 115-122 | | Wrong study design |
| **effect of developmental therapy for preterm infants at 35-36 weeks gestational age** | Ctri | 2018 | | | | | | Wrong study design |
| **Impact of Follow up Counseling package on the Follow up Rate and Neonatal Outcomes of High Risk Neonates** | Ctri | 2019 | | | | | | Wrong study design |
| **Parent delivered early movement experience on motor skill development in preterm low birth weight infants** | Ctri | 2021 | | | | | | Wrong study design |
| **Grip strength and its impact on fine motor skills in preterm children at 3-5 years** | Ctri | 2022 | | | | | | Wrong study design |

| | | | | | | | | |
|---|---|---|---|---|---|---|---|---|
| **EFFICACY OF RHYTHMIC MOVEMENT ON MOTOR PROFICIENCY IN PRIMARY SCHOOL GOING PRETERM BORN CHILDREN** | Ctri | 2022 | | | | | | Wrong study design |
| **Acceptability and feasibility of home based kangaroo mother care among mothers of low birth weight babies** | Ctri | 2022 | | | | | | Wrong study design |
| **Development of a parent education module to read and understand behavioral signs of their preterm infant** | Ctri | 2023 | | | | | | Wrong study design |
| **Implementing community supports to lessen health disparities at kindergarten entry for very preterm survivors** | Dmowska et al. | 2016 | International Journal of Child Health and Human Development | 9 | 4 | 535-544 | | Wrong study design |
| **Biological and behavioral pathways from prenatal depression to offspring cardiometabolic risk: Testing the developmental origins of health and disease hypothesis** | Doom et al. | 2024 | Developmental Psychology | | | No Paginat | 10.1037/dev0001704 | Wrong study design |
| **Exploring parents' musical agency in resource-oriented music therapy with their preterm infants in the nicu and at home** | Epstein, Shulamit | 2023 | Nordic Journal of Music Therapy | | | No Paginat | 10.1080/08098131.2023.2280979 | Wrong study design |

| | | | | | | | | |
|---|---|---|---|---|---|---|---|---|
| **Effects of an Infant Care Education Program for Mothers of Late-preterm Infants on Parenting Confidence, Breastfeeding Rates, and Infants' Growth and Readmission Rates** | Eun Hye, Jang; Hyeon Ok, Ju | 2020 | Child Health Nursing Research | 26 | 1 | Nov 22 | 10.4094/chnr.2020.26.1.11 | Wrong study design |
| **Effects of a Community-based Follow-up Program for Parents with Premature Infants on Parenting Stress, Parenting Efficacy, and Coping** | Eun Sun, Ji; Ka Ka, Shim | 2020 | Child Health Nursing Research | 26 | 3 | 366-375 | 10.4094/chnr.2020.26.3.366 | Wrong study design |
| **Development of a multidisciplinary medical home program for NICU graduates** | Feehan et al. | 2020 | Maternal and Child Health Journal | 24 | 1 | Nov 21 | 10.1007/s10995-019-02818-0 | Wrong study design |
| **Effectiveness of the early intervention program with preterm infants** | Formiga et al. | 2004 | Paideia: Cadernos de Psicologia e Educacao | 14 | 29 | 301-311 | 10.1590/S0103-863X2004000300006 | Wrong study design |
| **Developmental care for very low-birth-weight infants** | Garland et al. | 1995 | JAMA | 273 | 20 | 1575?1578 | | Wrong study design |
| **The effect of telenursing on the rate of newborn readmission** | Gholami et al. | 2022 | Journal of Neonatal Nursing | 28 | 1 | 26-30 | 10.1016/j.jnn.2021.03.001 | Wrong study design |
| **Adaptation and Acceptability of a Digitally Delivered Intervention for Parents of Very Low Birth Weight Infants** | Greene et al. | 2020 | Nursing Research | 69 | | S47-S56 | 10.1097/NNR.000000000000445 | Wrong study design |

| | | | | | | | | |
|---|---|---|---|---|---|---|---|---|
| **Neurodevelopmental Outcomes at Two Years' Corrected Age of Very Preterm Infants after Implementation of a Post-Discharge Responsive Parenting Intervention Program (TOP Program)** | Halbmeijer et al. | 2023 | J Pediatr | 257 | | 113381 | 10.1016/j.jpeds.2023.02.025 | Wrong study design |
| **Early Intervention Post-Hospital Discharge for Infants Born Preterm** | Hilderman, Courtney G. E.; Harris, Susan | 2014 | Physical Therapy | 94 | 9 | 1211-1219 | 10.2522/ptj.20130392 | Wrong study design |
| **CareToy project for early intervention in infants born preterm: Preliminary findings on parent-infant interaction** | Inguaggiato et al. | 2015 | Giornale di Neuropsichiatria dell'Eta Evolutiva | 35 | 2 | 147-154 | | Wrong study design |
| **Influence of family based intervention on neuro-developmental status of preterm neonates** | Irct201705079568N | 2017 | trial registry | | | | | Wrong study design |
| **Studying the effect of Telenursing on hope and perceived self-efficacy of mothers with premature infants after discharging from neonatal intensive care unit** | Irct2016102403047 4N | 2021 | | | | | | Wrong study design |
| **The effect of multisensory intervention (ATVV) on self-esteem and mental health of mothers of preterm infants** | Irct2021012705015 2N | 2021 | trial registry | | | | | Wrong study design |
| **Supporting parents of preterm infants with digital information** | Isrctn | 2022 | trial registry | | | | | Wrong study design |

| | | | | | | | | |
|---|---|---|---|---|---|---|---|---|
| **The Influence of Components of The Neuro-Developmental Treatment Program and Sensoric Integration on Children with Damaged Central Neural-System** | Jokovic-Turalija, Ines; Horvat, Dejana; Stefanec, Maja | 2003 | Hrvatska Revija Za Rehabilitacijska Istrazivanja | 39 | 2 | 203-209 | | Wrong study design |
| **Development and mother and child interaction of low birth weight infants purpose of child care support** | Jprn, Umin | 2017 | | | | | | Wrong study design |
| **Analysis of movement of high-risk premature infants in response to tactile stimulation** | Kalscheur, J. A. | 1985 | | 23 | | 282 | | Wrong study design |
| **A treatment response to attachment deficits, depression and preterm birth: Equine assisted psychotherapy.DP - 2022** | Kasper, Dana L. | 2022 | | 83 | 8-B | No Pagination Specified | | Wrong study design |
| **The Infant Behavioral Assessment and Intervention Program to support preterm infants after hospital discharge: a pilot study** | Koldewijn et al. | 2005 | Dev Med Child Neurol | 47 | 2 | 105-12 | 10.1017/s0012162205000198 | Wrong study design |
| **Effect of early intervention using state modulation and cue reading on mother-infant interactions in preterm infants and their mothers in Japan** | Kusanagi et al. | 2011 | J Med Dent Sci | 58 | 3 | 89-96 | | Wrong study design |
| **MSc dissertation abstract session. A study of two different chest physiotherapy percussion techniques on some physiological changes in neonates** | Leung, A. K. P. | 1998 | Hong Kong Physiotherapy Journal | 16 | | 19-19 | | Wrong study design |

| | | | | | | | | |
|---|---|---|---|---|---|---|---|---|
| **The efficacy of early intervention for low birth weight infants: What works, how, and for whom?DP - Apr 1992** | Liaw, Fong-ruey | 1992 | | 52 | 10-A | 3521 | | Wrong study design |
| **Patterns of low-birth-weight children's cognitive development** | Liaw, Fong-ruey; Brooks-Gunn, Jeanne | 1993 | Developmental Psychology | 29 | 6 | 1024-1035 | 10.1037/0012-1649.29.6.1024 | Wrong study design |
| **Taking care of their baby at home but with nursing staff as support: The use of videoconferencing in providing neonatal support to parents of preterm infants** | Lindberg et al. | 2009 | Journal of Neonatal Nursing | 15 | 2 | 47-55 | 10.1016/j.jnn.2009.01.004 | Wrong study design |
| **Effects of tactile-kinesthetic stimulation in preterms: a controlled trial** | Mathai et al. | 2001 | Indian Pediatr | 38 | 10 | 1091-8 | | Wrong study design |
| **Neuromotor development and visual acuity in premature infants submitted to early visuo-motor stimulation** | Mazzitelli et al. | 2008 | Psychology & Neuroscience | 1 | 1 | 41-45 | 10.3922/j.psns.2008.1.007 | Wrong study design |
| **Home Visiting for NICU Graduates: Impacts of Following Baby Back Home** | McKelvey, Lorraine M.; Lewis, Kanna N.; Beavers, Jared; Casey, Patrick H.; Irby, Carmen; Goudie, Anthony | 2021 | Pediatrics | 148 | 1 | 01. Okt | 10.1542/peds.2020-029397 | Wrong study design |
| **Effectiveness of Part C Early Intervention Physical, Occupational, and Speech Therapy Services for Preterm or Low Birth Weight Infants in Wisconsin, United** | McManus, Beth M.; Carle, Adam C.; Poehlmann, Julie | 2012 | Academic Pediatrics | 12 | 2 | 96-103 | 10.1016/j.acap.2011.11.004 | Wrong study design |
| **CDC Kerala 2: developmental Intervention Package for Babies <1,800 g – Outcome at 6 mo Using DASII** | Nair et al. | 2014 | Indian journal of pediatrics | 81 | 2 | 73?79 | 10.1007/s12098-014-1624-z | Wrong study design |

| | | | | | | | | |
|---|---|---|---|---|---|---|---|---|
| **The Effects of Family Centered Intervention Program on Preterm Infants** | Nct | 2017 | | | | | | Wrong study design |
| **What Are the Effects of Supporting Early Parenting by Increasing the Understanding of the Infant?** | Nct | 2017 | trial registry | | | | | Wrong study design |
| **Effect of Tactile-Kinesthetic Stimulation on Preterm Neonates** | Nct | 2021 | trial registry | | | | | Wrong study design |
| **Using NICU Discharge Education Tools to Enhance Discharge Preparation for Parents of Moderate to Late Preterm Infants** | Nct | 2023 | | | | | | Wrong study design |
| **Interdisciplinary E-health Based Follow-up of Preterm Born Children** | Nct | 2024 | | | | | | Wrong study design |
| **Development and evaluation of the feasibility of an ageappropriate additional, preventive intervention for very preterm children at 18 months** | Nl, Omon | 2013 | | | | | | Wrong study design |
| **E-TOP - Digital information for parents of very and moderat preterm born infants** | Nl, Omon | 2022 | | | | | | Wrong study design |
| **Neurophysiological development in premature infants following stimulation** | Rice, Ruth D. | 1977 | Developmental Psychology | 13 | 1 | 69-76 | 10.1037/0012-1649.13.1.69 | Wrong study design |
| **Home intervention for premature infants of low-income families** | Ross, Gail S. | 1984 | American Journal of Orthopsychiatry | 54 | 2 | 263-270 | 10.1111/j.1939-0025.1984.tb01493.x | Wrong study design |

| | | | | | | | | |
|---|---|---|---|---|---|---|---|---|
| **Healthy start program and feto-infant morbidity outcomes: Evaluation of program effectiveness** | Salihu et al. | 2009 | Maternal and Child Health Journal | 13 | 1 | 56-65 | 10.1007/s10995-008-0400-y | Wrong study design |
| **The effects of early stimulation on low-birth-weight infants.DP - Mar 1973** | Scarr-Salapatek, Sandra; Williams, Margaret L. | 1973 | Child Developm ent | 44 | 1 | 94-101 | 10.2307/112768 4 | Wrong study design |
| **A pilot study on early home-based intervention through an intelligent baby gym (CareToy) in preterm infants** | Sgandurra et al. | 2016 | Res Dev Disabil | 53-54 | | 32-42 | 10.1016/j.ridd.20 16.01.013 | Wrong study design |
| **Influences of a dedicated parental training program on parent-child interaction in preterm infants** | Steinhardt et al. | 2015 | Early Human Developm ent | 91 | 3 | 205-210 | 10.1016/j.earlhu mdev.2015.01.01 2 | Wrong study design |
| **Neurodevelopmental follow-up of very preterm infants after proactive treatment at a gestational age of > or =** | Steinmacher et al. | 2008 | Journal of Pediatrics | 152 | 6 | 771-2 | 10.1016/j.jpeds.2 007.11.004 | Wrong study design |
| **The emotional experiences and supports for parents with babies in a neonatal nursery** | Turner et al. | 2013 | Adv Neonat Care | 13 | 6 | 438-46 | 10.1097/ANC.00 0000000000003 0 | Wrong study design |
| **A program of early intervention, P.M.S.E. (Holdings, Manipulations, Stimulations with the "touch sensori-tonic-motor", Environment)** | Vaivre-Douret, L. | 2009 | A N A E Approche Neuropsy chologiqu e des Apprentis sages chez l'Enfant | 21 | 4-5[104-10 | 429-438 | | Wrong study design |

| | | | | | | | | |
|---|---|---|---|---|---|---|---|---|
| **Does the development of executive functioning in infants born preterm benefit from maternal directiveness?** | van de Weijer-Bergsma et al. | 2016 | Early Human Development | 103 | | 155-160 | 10.1016/j.earlhumdev.2016.09.012 | Wrong study design |
| **Neonatal music therapy and cerebral oxygenation in extremely and very preterm infants: A pilot study** | van Dokkum et al. | 2021 | Music and Medicine | 13 | 2 | 91-98 | 10.47513/mmd.v13i2.813 | Wrong study design |
| **Factors associated with rehospitalizations of very low birthweight infants: Impact of a transition home support and education** | Vohr et al. | 2012 | Early Human Development | 88 | 7 | 455-460 | 10.1016/j.earlhumdev.2011.10.011 | Wrong study design |
| **Effects of developmental music groups for parents and premature or typical infants under two years on parental responsiveness and infant social development** | Walworth, D. D. | 2009 | J Music Ther | 46 | 1 | 32-52 | 10.1093/jmt/46.1.32 | Wrong study design |
| **Preschool motor skills following physical and occupational therapy services among non-disabled very low birth weight children** | Watkins et al. | 2014 | Maternal and Child Health Journal | 18 | 4 | 821-828 | 10.1007/s10995-013-1306-x | Wrong study design |
| **Evaluating neonatal developmental care** | Westrup et al. | 2003 | Journal of Pediatrics | 142 | | 591-592 | | Wrong study design |
| **Relationship between mother-infant mutual dyadic responsiveness and premature infant development as measured by the Bayley III at 6weeks corrected age** | White-Traut et al. | 2018 | Early Hum Dev | 121 | | 21-26 | 10.1016/j.earlhumdev.2018.04.018 | Wrong study design |

| | | | | | | | | |
|---|---|---|---|---|---|---|---|---|
| **Kentucky Health Access Nurturing Development Services Home Visiting Program improves maternal and child health** | Williams et al. | 2017 | Maternal and Child Health Journal | 21 | 5 | 1166-1174 | 10.1007/s10995-016-2215-6 | Wrong study design |
| **Early intervention in preterm infants after discharge from hospital...Als H, Duffy FH, McAnulty GB et al. Early experience alters brain function and structure. Pediatrics. 2004;113:846-857** | Wolf, et al. | 2004 | Pediatrics | 114 | 6 | 1738-1739 | 10.1542/peds.2004-1629 | Wrong study design |
| **Influence of music therapy on physical and neurobehavioral development of preterm infants** | Yang et al. | 2014 | Chinese Nursing Research | 28 | 4A | 1227-1228 | 10.3969/j.issn.1009-6493.2014.10.03 | Wrong study design |
| **The Norwegian Physiotherapy Study in Preterm Infants, a randomized controlled study of early intervention: outcome at week 37 postmenstrual age** | | 2016 | Developmental Medicine & Child Neurology | 58 | | 59-59 | 10.1111/dmcn.129_13241 | Conference abstract |
| **Impact of assisted exercise on total energy expenditure and body composition in preterm infants** | Ahmad et al. | 2015 | | | | | | Conference abstract |
| **The coping with and caring for infants with special needs (COPCA) intervention is associated with improved motor development in preterm infants** | Akhbari Ziegler, S. | 2022 | Developmental medicine and child neurology | 64 | SUPPL 3 | 88 | 10.1111/dmcn.15215 | Conference abstract |
| **Explorer baby early intervention program effects on infants born preterm: a stratified randomized controlled study** | Altunalan et al. | 2021 | Developmental medicine and child neurology | 63 | SUPPL 3 | 51?52 | 10.1111/dmcn.15004 | Conference abstract |

| | | | | | | | | |
|---|---|---|---|---|---|---|---|---|
| **Short term effect of SAFE early intervention approach in infants born preterm in Turkey: a randomised controlled single blinded study** | Apaydin et al. | 2021 | Developmental medicine and child neurology | 63 | SUPPL 3 | 72?73 | 10.1111/dmcn.15005 | Conference abstract |
| **Music Therapy in Preterm Neonates: is There a Correlation between Music Tempo and Biological Effects?** | Aung et al. | 2011 | | | | | | Conference abstract |
| **Chest physiotherapy is associated with encephaloclastic porencephaly in extremely premature babies** | Becroft et al. | 1998 | Modern pathology | 11 | | 1P | | Conference abstract |
| **8 Year Growth Status of Preterm Infants Who Receive Early Educational Intervention** | Bradley | 2007 | Pediatric academic society | http://www | 5140.2 | | | Conference abstract |
| **Short-term effects of home-based early intervention with caretoy system: RCT in preterm infants** | Cioni et al. | 2017 | Developmental medicine and child neurology | 59 | | 73?74 | 10.1111/dmcn.113-13511 | Conference abstract |
| **Early post hospital discharge psychomotor therapy intervention program: effect on the development in very preterm infants (VPI) at 2-years corrected age** | Corinne, A.; Virginie, E.; Nathalie, N.; Catherine, B.; Bruno, C.; Francoise, H. A.; Julie, A.; Isabelle, G.; Charlotte, C.; Nathalie, M.; et al. | 2016 | European journal of pediatrics | 175 | 11 | 1525 | 10.1007/s00431-016-2785-8 | Conference abstract |

| | | | | | | | |
|---|---|---|---|---|---|---|---|
| **Randomized Controlled Trial of Surfactant Administration by Laryngeal Mask Airway (LMA) Massage and Kinesthetic Stimulation (Exercise) Improves Weight Gain in Very Low Birth Weight (VLBW) Preterm Infants; Results from a Randomized Controlled Trial** | Corrine et al. | 2008 | American pediatric society/society for pediatric research abstract | | | | Conference abstract |
| **Early literacy intervention in very low birth weight (VLBW) Infants: a randomised control trial** | Daniel et al. | 2015 | | | | | Conference abstract |
| **PASSIVE RANGE-OF-MOTION EXERCISE AND BONE MINERALIZATION IN PRETERM INFANTS: a RANDOMIZED CONTROLLED TRIAL** | El Farrash et al. | 2023 | Aging clinical and experimental research | 35 | S490 | 10.1007/s40520-023-02442-7 | Conference abstract |
| **Early intervention program after discharge improves parents-infant relationship and behavioral and cognitive outcomes for preterm infants** | Guillois et al. | 2013 | Archives of women's mental health | 16 | S92 | 10.1007/s00737-013-0355-x | Conference abstract |
| **Early intervention effects on emotion regulation for VLBW preterm infants at age 6 months during arm restrain procedure** | Ho et al. | 2015 | Physiotherapy (united kingdom) | 101 | eS578 | 10.1016/j.physio.2015.03.3400 | Conference abstract |
| **Early interventions for very low birth weight preterm infants: effects and mediators** | Jeng et al. | 2015 | Physiotherapy (united kingdom) | 101 | eS676 | 10.1016/j.physio.2015.03.3518 | Conference abstract |

| | | | | | | | | |
|---|---|---|---|---|---|---|---|---|
| **PreEMPT: preterm infant Early intervention for Movement and Participation Trial-a feasibility study in regional Australia** | Johnston et al. | 2020 | Developmental medicine and child neurology | 62 | | 58 | 10.1111/dmcn.14469 | Conference abstract |
| **Structured neonatal physical therapy (SNP) program in moderate to late preterm (MLP) infants: outcome on motor, cognition and language development at 3 and 6 months of age** | Khurana et al. | 2020 | Developmental medicine and child neurology | 62 | SUPPL 3 | 11 | 10.1111/dmcn.14661 | Conference abstract |
| **The reliability, sensitivity and responsiveness of the infant behavioral assessment (IBA) to evaluate neurobehavioral organization in very preterm born infants** | Koldewijn et al. | 2011 | Physiotherapy (united kingdom) | 97 | | eS628?eS6 | 10.1016/j.physio.2011.04.002 | Conference abstract |
| **A randomized controlled trial of an early intervention program in low birth weight children: outcome at 12 months** | Kyriakidou et al. | 2011 | Journal of perinatal medicine | 39 | | | 10.1515/jpm-2012-1008 | Conference abstract |
| **PARTICIPATION OF INFANTS AT BIOLOGICAL RISK IS FACILITATED BY REMOTE INTERVENTION CARRIED OUT BY PARENTS – STEP PROTOCOL: RANDOMIZED CLINICAL TRIAL...1st Student Scientific Conference of the Brazilian Association for Research and** | Lima et al. | 2024 | Brazilian Journal of Physical Therapy | 28 | | N.PAG-N.P | 10.1016/j.bjpt.2024.100674 | Conference abstract |
| **Effectiveness of a family-centered intervention program in very low birth weight preterm infants at term age: a randomized controlled trial** | Liu et al. | 2015 | Physiotherapy (united kingdom) | 101 | | eS894?eS8 | 10.1016/j.@physio.2015.03.1730 | Conference abstract |

| | | | | | | | | |
|---|---|---|---|---|---|---|---|---|
| **The Leiden developmental care study: the effect of developmental care on the ventilation of preterm infants <32 weeks gestational age** | Maguire et al. | 2003 | Pediatric research | 53 | | 138 | | Conference abstract |
| **A Randomized Controlled Trial Comparing Early Maternal Holding Styles on Maternal-Infant Interaction at 6 Months Infant Age** | Neu, M. M. | 2011 | JOGNN: journal of obstetric, gynecologic & neonatal nursing | 40 | | S113 | 10.1111/j.1552-6909.2011.01243_35.x | Conference abstract |
| **Low Birth Weight Children: effect of Early Intervention on Behavioral Outcome at 5 y – A Randomized Controlled Trial** | Nordhov | 2010 | Pediatric academic society | http://www | 4350.3 | | | Conference abstract |
| **Low Birth Weight Children: effect of an Early Intervention Program on Cognitive Outcome A Randomized Controlled Trial** | Nordhov et al. | 2009 | | | | | | Conference abstract |
| **Usefulness of an Early Intervention on Mother-Child Relationship To Improve Neurodevelopmental Outcome of Very-Low-Birth Weight Children** | Picciolini et al. | 2005 | | | NO: | | | Conference abstract |
| **Early intervention for at risk premature neonates** | Rothberg et al. | 1984 | Pediatric research | 18 | | 344 | | Conference abstract |
| **Home-based early intervention in infants at highrisk for cerebral palsy: a protocol of a randomised controlled trial study with Care Toy** | Sgandurra et al. | 2018 | Developmental medicine and child neurology | 60 | | 20?21 | 10.1111/dmcn.13789 | Conference abstract |

| | | | | | | | |
|---|---|---|---|---|---|---|---|
| **Effect of CareToy early intervention on visual development in preterm infants** | Sgandurra et al. | 2016 | Developmental medicine and child neurology | 58 | 58 | 10.1111/dmcn.12341 | Conference abstract |
| **Physical exercise for improving bone strength in preterm neonates less than 35 weeks gestation– a randomized controlled trial** | Shaw et al. | 2015 | | | | | Conference abstract |
| **Effect of Neonatal Developmental Intervention Program on Growth and Motor Development in Premature from Neonatal Period to 6 Months Corrected** | Shin et al. | 2012 | | | | | Conference abstract |
| **Experimental study on mother-infant signalling during breastfeeding: biological and psychological aspects** | Shukri et al. | 2020 | Maternal & child nutrition | 16 | | 10.1111/mcn.12933 | Conference abstract |
| **A Randomised Controlled Trial of a Home-Based Preventative Care Program for Preterm Infants and Their Caregivers: outcomes at 4 Years** | Spencer-Smith, M.; Spittle, A.; Doyle, L.; Lee, K.; Lorefice, L.; Suetin, A. | 2012 | | | | | Conference abstract |
| **Long-term benefits of home-based preventative care for preterm infants: outcomes of a randomised trial** | Spencer-Smith et al. | 2012 | | 54 | 24?25 | | Conference abstract |
| **The effects of a rct of a preventative care program on motor outcome for preterm infants over the first year** | Spittle et al. | 2011 | Physiotherapy (united kingdom) | 97 | eS1165?eS | 10.1016/j.physio.2011.04.002 | Conference abstract |

| | | | | | | | | |
|---|---|---|---|---|---|---|---|---|
| **An early preventative care program for infants born very preterm and their parents improves long term parental mental health** | Spittle et al. | 2016 | Developmental medicine and child neurology | 58 | | 23 | 10.1111/dmcn.13069 | Conference abstract |
| **A randomized controlled trial of a preventative care program at home over the first year of life in very preterm infants: outcomes of children and caregivers at 2 years of age** | Spittle et al. | 2010 | Developmental medicine and child neurology | 52 | | 52?53 | 10.1111/j.1469-8749.2010.03759.x | Conference abstract |
| **A randomized controlled trial of a preventative care program at home over the first year of life in very preterm infants: outcomes of children and caregivers at 2 years of age** | Spittle et al. | 2010 | Developmental medicine and child neurology | 52 | | 70?71 | 10.1111/j.1469-8749.2010.03682.x | Conference abstract |
| **A randomised controlled trial of a home-based preventative care program for preterm infants and their parents: outcomes at 4 years** | Spittle et al. | 2012 | Journal of paediatrics and child health | 48 | | 37?38 | 10.1111/j.1440-1754.2012.02411.x | Conference abstract |
| **Promoting Development in High Risk Preterm Infants: new Results of an Early Intervention Program** | Teti; Et, A. L. | 2005 | | | NO: | | | Conference abstract |
| **Improved parent-child relationships from a web-based early intervention after preterm birth** | Treyvaud et al. | 2020 | Journal of paediatrics and child health | 56 | SUPPL 1 | 47?48 | 10.1111/jpc.14831 | Conference abstract |

| | | | | | | | | |
|---|---|---|---|---|---|---|---|---|
| **The infant behavioral assessment and intervention program improves functional skills in infants born preterm at the age of 44 months** | Verkerk et al. | 2011 | Physiotherapy (united kingdom) | 97 | | eS1309 | 10.1016/j.physio.2011.04.002 | Conference abstract |
| **Preschool outcome in children born very prematurely and cared for according to NIDCAP** | Westrup et al. | 2003 | Pediatric research | 54 | 4 | 557 | | Conference abstract |
| **Integrated Mother-Premature Infant Intervention and Mother-Infant Interaction at 6-Week Corrected Age** | White-Traut et al. | 2013 | JOGNN: journal of obstetric, gynecologic & neonatal nursing | 42 | | S90 | 10.1111/1552-6909.12183 | Conference abstract |
| **Integrated Mother-Premature Infant Intervention and Mother-Infant Interaction at 6- Weeks Corrected Age** | White-Traut et al. | 2012 | Pediatric academic societies annual meeting | | | | | Conference abstract |
| **The effect of home program adherence on developmental outcomes in preterm infants following early intervention** | Yu et al. | 2011 | Physiotherapy (united kingdom) | 97 | | eS1375 | 10.1016/j.physio.2011.04.002 | Conference abstract |
| **Short-term effect of a family-centered intervention program on the cortical auditory processing function in very low birth weight preterm infants** | Yu et al. | 2015 | Physiotherapy (united kingdom) | 101 | | eS1708?eS | 10.1016/j.physio.2015.03.121 | Conference abstract |

| | | | | | | | | |
|---|---|---|---|---|---|---|---|---|
| **The impact of music therapy on stress, functional gastrointestinal disorders and psychomotor development: a prospective randomized controlled study in preterm infants** | Zampatti et al. | 2021 | Journal of pediatric gastroenterology and nutrition | 72 | SUPPL 1 | 212 | 10.1097/MPG.0000000000003177 | Conference abstract |
| **Vestibular Stimulation Alters Sensorimotor Integration of the Respiratory and Orofacial Central Pattern Generators in Preterm Infants** | Zimmerman et al. | 2011 | | | | | | Conference abstract |
| **Nine-year outcome of the Vermont intervention program for low birth weight infants** | Achenbach et al. | 1993 | Pediatrics | 91 | 1 | 45-55 | | <6 interventions post-discharge |
| **Seven-year outcome of the Vermont intervention program for low-birthweight infants.DP - Dec 1990** | Achenbach et al. | 1990 | Child Development | 61 | 6 | 1672-1681 | 10.2307/1130830 | <6 interventions post-discharge |
| **Improving neurobehavioural development in preterm infants** | Actrn | 2006 | trial registry | | | | | <6 interventions post-discharge |
| **Increasing Mothers' Confidence and Ability by Creating Opportunities for Parent Empowerment (COPE): A Randomized, Controlled Trial** | Askary Kachoosangy et al. | 2020 | Iran | 14 | 1 | 77-83 | | <6 interventions post-discharge |
| **Pilot RCT of the use of video interactive guidance with preterm babies** | Barlow et al. | 2016 | Journal of Reproductive and Infant Psychology | 34 | 5 | 511-524 | 10.1080/02646838.2016.1217404 | <6 interventions post-discharge |

| | | | | | | | | |
|---|---|---|---|---|---|---|---|---|
| **Early intervention with interactive guidance and neuroendocrine reactivity modification of formal preterm born infants now 12 months old and their mothers** | Borghini et al. | 2009 | Cahiers Critiques de therapie familiale et de pratiques de reseaux | 43 | | 117-149 | 10.3917/ctf.043.0117 | <6 interventions post-discharge |
| **Early preventive attachment-oriented psychotherapeutic intervention program with parents of a very low birthweight premature infant: results of attachment and neurological development** | Brisch et al. | 2003 | Attach Hum Dev | 5 | 2 | 120-35 | 10.1080/1461673031000108504 | <6 interventions post-discharge |
| **The Impact of an Interactive Guidance Intervention on Sustained Social Withdrawal in Preterm Infants in Chile: Randomized Controlled Trial** | Bustamante Loyola et al. | 2022 | Front | 10 | | 803932 | 10.3389/fped.2022.803932 | <6 interventions post-discharge |
| **Infant mental health intervention for preterm infants in Japan: Promotions of maternal mental health, mother-infant interactions, and social support by providing continuous home visits until the corrected infant age of 12 months** | Cho et al. | 2013 | Infant Mental Health Journal | 34 | 1 | 47-59 | 10.1002/imhj.21352 | <6 interventions post-discharge |

| | | | | | | | | |
|---|---|---|---|---|---|---|---|---|
| **Effect of a home visit educational program on mortality and morbidity of preterm newborn** | Edraki et al. | 2012 | Journal of Shaheed Sadoughi University of Medical Sciences & Health Services | 19 | 6 | 1p-1p | | <6 interventions post-discharge |
| **Follow-up of the Cues and Care trial: Mother and infant outcomes at 6 months** | Feeley et al. | 2012 | Journal of Early Intervention | 34 | 2 | 65-81 | 10.1177/1053815112453767 | <6 interventions post-discharge |
| **Behavioral and electrophysiological study of attention process in preterm infants with cerebral white matter injury** | Gutierrez-Hernandez et al. | 2018 | Psychology & Neuroscience | 11 | 2 | 132-145 | 10.1037/pne0000127 | <6 interventions post-discharge |
| **Early stress exposure and later cortisol regulation: Impact of early intervention on mother-infant relationship in preterm infants** | Habersaat et al. | 2014 | Psychological Trauma: Theory, Research, Practice, and Policy | 6 | 5 | 457-464 | 10.1037/a0033878 | <6 interventions post-discharge |
| **Effects of an online family-focused parenting support intervention on preterm infants' physical development and parents' sense of competence and care ability: A randomized controlled** | Huang et al. | 2024 | Int J Nurs Stud | 149 | | 104625 | 10.1016/j.ijnurstu.2023.104625 | <6 interventions post-discharge |

| | | | | | | | | |
|---|---|---|---|---|---|---|---|---|
| **The effect of Home Visit Program based on the Continued kangaroo Mother Care on Maternal Resiliency and Development of Premature Infant** | Irct20181121041718N | 2019 | trial registry | | | | | <6 interventions post-discharge |
| **Effects of a Continuity of Preterm Infant Care Program on Parenting outcomes and Service Utilization Rates** | Kaewwimol et al. | 2022 | Open public health journal | 15 | | | 10.2174/18749445-v15-e2206080 | <6 interventions post-discharge |
| **Effect of an early intervention programme on development of moderate and late preterm infants at 36 months: a randomized controlled study** | Kyno et al. | 2012 | Infant behav | 35 | 4 | 916-26 | 10.1016/j.infbeh.2012.09.004 | <6 interventions post-discharge |
| **Stability and Change in Longitudinal Associations between Child Behavior Problems and Maternal Stress in Families with Preterm Born Children, Follow-Up after a RCT-Study** | Landsem et al. | 2019 | Children (Basel) | 6 | 2 | 31 | 10.3390/children6020019 | <6 interventions post-discharge |
| **Early intervention program reduces stress in parents of preterms during childhood, a randomized controlled trial** | Landsem et al. | 2014 | Trials | 15 | | 387 | 10.1186/1745-6215-15-387 | <6 interventions post-discharge |
| **Temperamental Development among Preterm Born Children. An RCT Follow-Up Study** | Landsem et al. | 2020 | Children (Basel) | 7 | 4 | 23 | 10.3390/children7040036 | <6 interventions post-discharge |
| **Does An Early Intervention Influence Behavioral Development Until Age 9 in Children Born Prematurely?** | Landsem et al. | 2015 | Child Dev | 86 | 4 | 1063-1079 | 10.1111/cdev.12368 | <6 interventions post-discharge |
| **Effects of the Newborn Individualized Developmental Care and Assessment Program (NIDCAP) at age 8 years: preliminary data** | McAnulty et al. | 2010 | Clin Pediatr (Phila) | 49 | 3 | 258-70 | 10.1177/0009922809335668 | <6 interventions post-discharge |

| | | | | | | | | |
|---|---|---|---|---|---|---|---|---|
| **Maternal anxiety and depression after a premature infant's discharge from the neonatal intensive care unit: explanatory effects of the creating opportunities for parent empowerment program** | Melnyk et al. | 2008 | Nurs Res | 57 | 6 | 383-94 | 10.1097/NNR.0b013e3181906f59 | <6 interventions post-discharge |
| **Early Intervention in Preterm Infants: short and Long Term Developmental Outcome After a Parental Training** | Nct | 2016 | trial registry | | | | | <6 interventions post-discharge |
| **Attention Control Training (ACT) and Very Preterm Infants** | Nct | 2019 | | | | | | <6 interventions post-discharge |
| **The Effects of SAFE Early Intervention Approach in Premature Infants in Turkey** | Nct | 2021 | trial registry | | | | | <6 interventions post-discharge |
| **A Mobile Web-Based Parenting Intervention to Strengthen Social-Emotional Development of Low Birth Weight Infants** | Nct | 2022 | trial registry | | | | | <6 interventions post-discharge |
| **Early intervention improves cognitive outcomes for preterm infants: randomized controlled trial** | Nordhov et al. | 2010 | Pediatrics | 126 | 5 | e1088-94 | 10.1542/peds.2010-0778 | <6 interventions post-discharge |
| **Early intervention improves behavioral outcomes for preterm infants: randomized controlled trial** | Nordhov et al. | 2012 | Pediatrics | 129 | 1 | e9-e16 | 10.1542/peds.2011-0248 | <6 interventions post-discharge |
| **An intervention program for mothers of low-birthweight infants: Preliminary results.DP - May 1984** | Nurcombe et al. | 1984 | Journal of the American Academy of Child Psychiatry | 23 | 3 | 319-325 | 10.1016/S0002-7138%2809%2960511-2 | <6 interventions post-discharge |

| | | | | | | | | |
|---|---|---|---|---|---|---|---|---|
| **Regulatory competence and social communication in term and preterm infants at 12 months corrected age. Results from a randomized controlled** | Olafsen et al. | 2012 | Infant behav | 35 | 1 | 140-9 | 10.1016/j.infbeh.2011.08.001 | <6 interventions post-discharge |
| **Joint attention in term and preterm infants at 12 months corrected age: the significance of gender and intervention based on a randomized controlled trial** | Olafsen et al. | 2006 | Infant behav | 29 | 4 | 554-63 | 10.1016/j.infbeh.2006.07.004 | <6 interventions post-discharge |
| **A randomized, controlled trial of the effectiveness of an early-intervention program in reducing parenting stress after preterm birth** | Kaaresen et al. | 2006 | Pediatrics | 118 | 1 | e9-19 | 10.1542/peds.2005-1491 | <6 interventions post-discharge |
| **Maternal ratings of infant regulatory competence from 6 to 12 months: influence of perceived stress, birth-weight, and intervention: a randomized controlled trial** | Olafsen et al. | 2008 | Infant behav | 31 | 3 | 408-21 | 10.1016/j.infbeh.2007.12.005 | <6 interventions post-discharge |
| **Attention and social communication skills of very preterm infants after training attention control: Bayesian analyses of a feasibility study** | Perra et al. | 2022 | PLoS ONE | 17 | 9 | e0273767 | 10.1371/journal.pone.0273767 | <6 interventions post-discharge |
| **Minimizing adverse effects of low birthweight: four-year results of an early intervention program** | Rauh et al. | 1988 | Child Dev | 59 | 3 | 544-53 | | <6 interventions post-discharge |
| **Stress in fathers of moderately and late preterm infants: A randomised controlled trial** | Ravn et al. | 2012 | Early Child Development and Care | 182 | 5 | 537-552 | 10.1080/03004430.2011.564279 | <6 interventions post-discharge |

| | | | | | | | | |
|---|---|---|---|---|---|---|---|---|
| **Effect of early intervention on social interaction between mothers and preterm infants at 12 months of age: a randomized controlled trial** | Ravn et al. | 2011 | Infant behav | 34 | 2 | 215-25 | 10.1016/j.infbeh.2010.11.004 | <6 interventions post-discharge |
| **Effects of early mother-infant intervention on outcomes in mothers and moderately and late preterm infants at age 1 year: a randomized controlled** | Ravn et al. | 2012 | Infant behav | 35 | 1 | 36-47 | 10.1016/j.infbeh.2011.09.006 | <6 interventions post-discharge |
| **Primary Care Triple P for parents of NICU graduates with behavioral problems: a randomized, clinical trial using observations of parent-child interaction** | Schappin et al. | 2014 | BMC Pediatr | 14 | | 305 | 10.1186/s12887-014-0305-4 | <6 interventions post-discharge |
| **Brief parenting intervention for parents of NICU graduates: a randomized, clinical trial of Primary Care Triple P** | Schappin et al. | 2013 | BMC Pediatr | 13 | | 69 | 10.1186/1471-2431-13-69 | <6 interventions post-discharge |
| **Promoting developmental outcomes of premature infants by Creating Opportunities for Parent Empowerment (COPE)** | Shafaroodi et al. | 2022 | Iranian Rehabilitation Journal | 20 | 1 | Nov 17 | 10.32598/irj.20.1.125.4 | <6 interventions post-discharge |
| **Early Intervention in Families with Preterm Infants: A Review of Findings from a Randomized Controlled Trial Following Children Up to 9 Years of Age** | Ulvund, S. E. | 2022 | Children (Basel) | 9 | 4 | 30 | 10.3390/children9040474 | <6 interventions post-discharge |
| **Health Care Use Outcomes of an Integrated Hospital-to-Home Mother-Preterm Infant Intervention** | Vonderheid et al. | 2016 | J Obstet Gynecol Neonatal Nurs | 45 | 5 | 625-38 | 10.1016/j.jogn.2016.05.007 | <6 interventions post-discharge |
| **Mother-infant interaction improves with a developmental intervention for mother-preterm infant dyads** | White-Traut et al. | 2013 | Infant behav | 36 | 4 | 694-706 | 10.1016/j.infbeh.2013.07.004 | <6 interventions post-discharge |

| | | | | | | | | |
|---|---|---|---|---|---|---|---|---|
| **A randomised controlled trial of an early preventative care program for infants born very preterm: the role of social risk on cognitive outcomes throughout early childhood** | | 2017 | Developmental Medicine & Child Neurology | 59 | | 44-44 | 10.1111/dmcn.64_13511 | Wrong timepoint of intervention |
| **Is the Newborn Individualized Developmental Care and Assessment Program (NIDCAP) effective for preterm infants with intrauterine growth restriction?** | Als et al. | 2010 | Journal of perinatology | | | | | Wrong timepoint of intervention |
| **Early experience alters brain function and structure** | Als et al. | 2004 | Pediatrics | 113 | 4 | 846-57 | 10.1542/peds.113.4.846 | Wrong timepoint of intervention |
| **A three-center, randomized, controlled trial of individualized developmental care for very low birth weight preterm infants: medical, neurodevelopmental, parenting, and caregiving effects** | Als et al. | 2003 | J Dev Behav Pediatr | 24 | 6 | 399-408 | 10.1097/00004703-200312000-00001 | Wrong timepoint of intervention |
| **Developmental care does not alter sleep and development of premature infants** | Ariagno et al. | 1997 | Pediatrics | 100 | | 1?7 | | Wrong timepoint of intervention |
| **Effect of a Supportive-Training Intervention on Mother-Infant Attachment** | Bostanabad et al. | 2017 | Iranian journal of pediatrics | 27 | 6 | 1?7 | 10.5812/ijp.10565 | Wrong timepoint of intervention |
| **Construction and Empirical Study of Sensitivity Intervention Program for Mothers of Hospitalized Premature Infants Based on Attachment Theory** | ChiCtr | 2020 | trial registry | | | | | Wrong timepoint of intervention |

| | | | | | | | | |
|---|---|---|---|---|---|---|---|---|
| **The effects of multisensory stimulation bundles on sleep and neurobehavioral development during first year after birth in very preterm infants: a randomized crossover-controlled study protocol** | ChiCtr | 2022 | trial registry | | | | | Wrong timepoint of intervention |
| **The Effects of Early Multi-Sensory Stimulation on the Physical and Neurological Development of Hospitalized Premature Infants** | ChiCtr | 2023 | trial registry | | | | | Wrong timepoint of intervention |
| **Does a parent-administrated early motor intervention influence general movements and movement character at 3months of age in infants born preterm?** | Fjortoft et al. | 2017 | Early Hum Dev | 112 | | 20-24 | 10.1016/j.earlhumdev.2017.06.008 | Wrong timepoint of intervention |
| **Effects of Early Intervention on Visual Function in Preterm Infants: A Randomized Controlled Trial** | Fontana et al. | 2020 | Front | 8 | | 291 | 10.3389/fped.2020.00291 | Wrong timepoint of intervention |
| **Neurodevelopmental Outcome Of Very Preterm And Moderate To Late Preterm Babies At The Corrected Age Of First Year** | Jeba et al. | 2022 | European journal of molecular and clinical medicine | 9 | 8 | 280?290 | | Wrong timepoint of intervention |
| **Auditory stimulation and developmental behavior of the premature infant.DP - May 1971** | Katz, Violet | 1971 | Nursing Research | 20 | 3 | 196-201 | 10.1097/00006199-197105000-00002 | Wrong timepoint of intervention |

| | | | | | | | | |
|---|---|---|---|---|---|---|---|---|
| **Creative Music Therapy with Premature Infants and Their Parents: A Mixed-Method Pilot Study on Parents' Anxiety, Stress and Depressive Symptoms and Parent-Infant Attachment** | Kehl et al. | 2020 | Int J Environ Res Public Health | 18 | 1 | 31 | 10.3390/ijerph18010265 | Wrong timepoint of intervention |
| **Follow-up outcomes at 1 and 2 years of infants born less than 32 weeks after Newborn Individualized Developmental Care and Assessment Program** | Maguire et al. | 2009 | Pediatrics | 123 | 4 | 1081-7 | 10.1542/peds.2008-1950 | Wrong timepoint of intervention |
| **[Early intervention for very-low-birth-weight infant]** | Matsuishi et al. | 1996 | No To Hattatsu | 28 | 2 | 149-55 | | Wrong timepoint of intervention |
| **Individualized developmental care for a large sample of very preterm infants: health, neurobehaviour and neurophysiology** | McAnulty et al. | 2009 | Acta Paediatr | 98 | 12 | 1920-6 | 10.1111/j.1651-2227.2009.01492.x | Wrong timepoint of intervention |
| **Self-Help Groups in a Premature Nursery: a Controlled Evaluation** | Minde, K. | 1978 | C2-SPECTR | | | | | Wrong timepoint of intervention |
| **Vermont Intervention: effect on Joint Attention Skills Between Parents and Moderate/Late Preterm Infants in the First Year of Life** | Nct | 2005 | trial registry | | | | | Wrong timepoint of intervention |
| **Very Preterm Children With Language Delay and Parent Intervention** | Nct | 2014 | trial registry | | | | | Wrong timepoint of intervention |
| **Creative Music Therapy for Premature Infants** | Nct | 2015 | trial registry | | | | | Wrong timepoint of intervention |
| **Effects of Early Behavioral and Transaction Interventions on Preterm Infants' and Parents' Biopsychosocial Well-being** | Nct | 2016 | trial registry | | | | | Wrong timepoint of intervention |

| | | | | | | | | |
|---|---|---|---|---|---|---|---|---|
| **Actions for Empowered Maternal Neonatal Care (ACUNE): a Nursing Intervention** | Nct | 2021 | trial registry | | | | | Wrong timepoint of intervention |
| **Parent Training for Parents of Toddlers Born Very Premature** | Nct | 2022 | trial registry | | | | | Wrong timepoint of intervention |
| **Oral Motor Intervention for Preterm Babies** | Nct | 2023 | | | | | | Wrong timepoint of intervention |
| **Effect Of Early Intervent?on Program Appl?ed To Premature Infants** | Nct | 2023 | trial registry | | | | | Wrong timepoint of intervention |
| **Early intervention for speech sound disorder in very preterm or very low birth weight children: a randomized controlled trial** | Nl, Omon | 2015 | | | | | | Wrong timepoint of intervention |
| **The effects of mothers' voice on the long term development of premature infants: A prospective randomized study** | Nocker-Ribaupierre et al. | 2015 | Music and Medicine | 7 | 3 | 20-25 | | Wrong timepoint of intervention |
| **Effect of Massage on treatment of preterm feeding intolerance: Study protocol for a randomized controlled** | Peng et al. | 2023 | Nurs | 10 | 7 | 4817-4824 | 10.1002/nop2.1733 | Wrong timepoint of intervention |
| **Comparative Effect of Massage Therapy versus Kangaroo Mother Care on Physiological Responses, Chest Expansion and Body Weight in Low Birthweight Preterm Infants** | Rangey et al. | 2014 | Disability, CBR & Inclusive Development | 25 | 3 | 103-110 | 10.5463/DCID.v25i3.290 | Wrong timepoint of intervention |
| **The effectiveness of structured discharge education on maternal confidence, caring knowledge and growth of premature newborns S-J Shieh et al. The effectiveness of structured discharge education on mothers and premature newborns** | Shwu-Jiuan et al. | 2010 | Journal of clinical nursing (john wiley & sons, inc.) | 19 | 23?24 | 3307?331[illegible] | 10.1111/j.1365-2702.2010.03382.x | Wrong timepoint of intervention |

| | | | | | | | | |
|---|---|---|---|---|---|---|---|---|
| **Developmental care affects pain and stress expression in preterm newborns** | Sizun et al. | 2001 | Pediatric research | 49 | 4 | 356A | | Wrong timepoint of intervention |
| **Tactile stimulation and behavioral development among low-birthweight infants** | Solkoff, N.; Matuszak, D. | 1975 | Child Psychiatry Hum Dev | 6 | 1 | 33-7 | 10.1007/BF01434430 | Wrong timepoint of intervention |
| **The preterm infant-parent programme for attachment-PIPPA Study: a randomised controlled trial** | Twohig et al. | 2021 | Pediatr Res | 90 | 3 | 617-624 | 10.1038/s41390-020-01262-z | Wrong timepoint of intervention |
| **Protocol for a randomized pilot study (FIRST STEPS): implementation of the Incredible Years-ASLD R program in Spanish children with autism and preterm children with communication and/or socialization difficulties** | Valencia et al. | 2021 | Trials | 22 | 1 | 291 | 10.1186/s13063-021-05229-1 | Wrong timepoint of intervention |
| **Parental stress and child behavior and temperament in the first year after the newborn individualized developmental care and assessment program** | van der Pal et al. | 2008 | Journal of Early Intervention | 30 | 2 | 102-115 | 10.1177/1053815107313485 | Wrong timepoint of intervention |
| **Preschool outcome in children born very prematurely and cared for according to the Newborn Individualized Developmental Care and Assessment** | Westrup et al. | 2004 | Acta Paediatr | 93 | 4 | 498-507 | 10.1080/08035250410023548 | Wrong timepoint of intervention |
| **No indications of increased quiet sleep in infants receiving care based on the newborn individualized developmental care and assessment program (NIDCAP)** | Westrup et al. | 2002 | Acta Paediatr | 91 | 3 | 318-22; dis | 10.1080/08035250252833996 | Wrong timepoint of intervention |
| **Preterm infants' orally directed behaviors and behavioral state responses to the integrated H-HOPE** | White-Traut et al. | 2014 | Infant behav | 37 | 4 | 583-96 | 10.1016/j.infbeh.2014.08.001 | Wrong timepoint of intervention |

| | | | | | | | | |
|---|---|---|---|---|---|---|---|---|
| **Exploring the Impact of Child-Centered Play Therapy on Academic Achievement of At-Risk Kindergarten Students** | Blanco et al. | 2019 | International journal of play therapy | 28 | 3 | 133?143 | 10.1037/pla0000086 | Wrong patient population |
| **School-age outcomes for early intervention participants who experienced intraventricular hemorrhage and low birth weight** | Boyce et al. | 2004 | Children's Health Care | 33 | 4 | 257-274 | 10.1207/s15326888chc3304_2 | Wrong patient population |
| **Enhancing the development of low-birthweight, premature infants: Changes in cognition and behavior over the first three years.DP - Jun 1993** | Brooks-Gunn et al. | 1993 | Child Development | 64 | 3 | 736-753 | 10.2307/1131215 | Wrong patient population |
| **Effects of an at-home video course on maternal learning, infant care and infant health** | Brown et al. | 2000 | Early Child Development and Care | 160 | | 47-65 | 10.1080/0030443001600105 | Wrong patient population |
| **Differential effects of high-quality early care: Lessons from the Infant Health and Development Program** | Chaparro et al. | 2019 | | | | 287-302 | 10.1017/9781108349352.014 | Wrong patient population |
| **Impact of tele-consultation for following up newborns during COVID-19 pandemic** | Ctri | 2020 | | | | | | Wrong patient population |
| **A psychomotor education program** | Fernandez Losa, Nicolas | 1996 | Psicothema | 8 | 1 | 77-88 | | Wrong patient population |
| **Effect of early neurodevelopmental therapy in normal and at-risk survivors of neonatal intensive care** | Goodman et al. | 1985 | Lancet | 2 | 8468 | 1327-30 | 10.1016/s0140-6736(85)92626-1 | Wrong patient population |
| **A randomized controlled trial of an early intervention program in low birth weight children: outcome at 2 years** | Kaaresen et al. | 2008 | Early Hum Dev | 84 | 3 | 201-9 | 10.1016/j.earlhumdev.2007.07.003 | Wrong patient population |

| | | | | | | | | |
|---|---|---|---|---|---|---|---|---|
| **A public health nursing early intervention program for adolescent mothers: outcomes from pregnancy through 6 weeks postpartum** | Koniak-Griffin et al. | 2000 | Nurs Res | 49 | 3 | 130-8 | 10.1097/00006199-200005000-00003 | Wrong patient population |
| **Effect of an Intensive Nurse Home Visiting Program on Adverse Birth Outcomes in a Medicaid-Eligible Population: A Randomized Clinical Trial** | McConnell et al. | 2022 | Jama | 328 | 1 | 27-37 | 10.1001/jama.2022.9703 | Wrong patient population |
| **Effect of an Intensive Nurse Home Visiting Program on Adverse Birth Outcomes in a Medicaid-Eligible Population: a Randomized Clinical Trial** | McConnell et al. | 2023 | Obstetrical & gynecological survey | 78 | 1 | 7?9 | 10.1097/OGX.00000000000001121 | Wrong patient population |
| **Implementation of Incredible Years for Autism and Language Delay Program in Spain** | Nct | 2020 | trial registry | | | | | Wrong patient population |
| **Multi Sensory Stimulation And Priming (MuSSAP) in Infants at Risk of Unilateral Cerebral Palsy** | Nct | 2022 | trial registry | | | | | Wrong patient population |
| **Early Intervention in Children at Risk of Developmental Delay** | Nct | 2023 | trial registry | | | | | Wrong patient population |
| **Nurse Parental Support Using a Mobile App to Enhance Parental Self-efficacy in Symptom Management for the Children With Medical Complexity: a Randomized Control Trial** | Nct | 2023 | trial registry | | | | | Wrong patient population |
| **Effect of an early intervention programme on low birthweight infants with cerebral injuries** | Ohgi et al. | 2004 | J Paediatr Child Health | 40 | 12 | 689-95 | 10.1111/j.1440-1754.2004.00512.x | Wrong patient population |

| | | | | | | | | |
|---|---|---|---|---|---|---|---|---|
| **A stimulation program for low birthweight infants** | Scarr-Salapatek, S.; Williams, M. L. | 1972 | Amer j pub health | | | 662?667 | | Wrong patient population |
| **Shortened hospital stay for low-birth-weight infants: nuts and bolts of a nursing intervention project** | Shapiro, C. | 1995 | J Obstet Gynecol Neonatal Nurs | 24 | 1 | 56-62 | 10.1111/j.1552-6909.1995.tb02379.x | Wrong patient population |
| **Prevention of behavior problems in a selected population: Stepping stones triple P for parents of young children with disabilities** | Shapiro et al. | 2014 | Res Dev Disabil | 35 | 11 | 2958-75 | 10.1016/j.ridd.2014.07.036 | Wrong patient population |
| **Effectiveness of 'Parent Empowerment' program on anxiety and stress in mothers who have preterm infants** | Soheila Jafari et al. | 2012 | Payesh (Health Monitor) | 11 | 2 | 1p-1p | | Wrong patient population |
| **Community initiated kangaroo mother care and early child development in low birth weight infants in India-a randomized controlled trial** | Taneja et al. | 2020 | BMC Pediatr | 20 | 1 | 150 | 10.1186/s12887-020-02046-4 | Wrong patient population |
| **Multisensory Stimulation and Priming (MuSSAP) in 4-10 Months Old Infants with a Unilateral Brain Lesion: A Pilot Randomised Clinical Trial.** | Verhaegh et al. | 2023 | Occup Ther Int | 2023 | | 8128407 | 10.1155/2023/8128407 | Wrong patient population |
| **Supporting Play Exploration and Early Developmental Intervention (SPEEDI) for Preterm Infants - Feasibility Study** | Actrn | 2018 | trial registry | | | | | No involvement of professionals |
| **Short-term effects of SAFE early intervention approach in infants born preterm: A randomized controlled single-blinded study** | Apaydin et al. | 2023 | Brain Behav | 13 | 10 | e3199 | 10.1002/brb3.3199 | No involvement of professionals |

| | | | | | | | | |
|---|---|---|---|---|---|---|---|---|
| **Effectiveness of 5 integrated-sensory stimulation program on social-emotional development in preterm infants in China** | ChiCtr | 2020 | trial registry | | | | | no involvement of professionals |
| **A study to determine the efficacy of Embrace Care device against the currently available solutions in maintaining normal body temperature in low-birth-weight babies in a home** | Ctri | 2013 | trial registry | | | | | No involvement of professionals |
| **A clinical trial to study the effects of mobile application based preterm home care program for premature infants** | Ctri | 2018 | trial registry | | | | | No involvement of professionals |
| **Impact of Sit down and play treatment on infant development** | Ctri | 2020 | trial registry | | | | | No involvement of professionals |
| **Effect of Kangaroo Mother Care on physical growth, breastfeeding and its acceptability** | Gathwala, G et al. | 2010 | Tropical doctor | 40 | 4 | 199?202 | 10.1258/td.2010.090513 | No involvement of professionals |
| **Effects of massage intervention on discharged premature infants' weight, parental stress, and parent-child attachment: A randomized controlled** | Hwu et al. | 2023 | Infant behav | 72 | | 101867 | 10.1016/j.infbeh.2023.101867 | No involvement of professionals |
| **Three-Part Program for Parents With Premature Infants** | Nct | 2003 | trial registry | | | | | No involvement of professionals |
| **Educational/Behavioral Intervention Program for Parents of Premature Infants** | Nct | 2005 | trial registry | | | | | No involvement of professionals |
| **Effect of Kangaroo Baby Massage on Mother-infant Interaction at Home** | Nct | 2021 | trial registry | | | | | No involvement of professionals |
| **Infant-Maternal Partnership and Cognitive Training Study for Preterm** | Nct | 2024 | trial registry | | | | | No involvement of professionals |

| | | | | | | | | |
|---|---|---|---|---|---|---|---|---|
| **One-year outcome of auditory-tactile-visual-vestibular intervention in the neonatal intensive care unit: effects of severe prematurity and central nervous system injury** | Nelson, et al. | 2001 | J Child Neurol | 16 | 7 | 493-8 | 10.1177/088307380101600706 | No involvement of professionals |
| **Home-based intervention after discharge for Latino families of low-birth-weight infants** | Zahr, Lina Kurdahi | 2000 | Infant Mental Health Journal | 21 | 6 | 448-463 | 10.1002/1097-0355%28200011/12%2921:6%3C448::AID- | No involvement of professionals |
| **Sensory stimulation program improves developments of preterm infants in Southwest China: A randomized controlled trial** | Zheng et al. | 2022 | Front Psychol | 13 | | 867529 | 10.3389/fpsyg.2022.867529 | No involvement of professionals |
| **Effectiveness of GAME (Goals Activity Motor Enrichment) for infants at high risk of cerebral palsy** | Actrn | 2017 | | | | | | Ongoing |
| **TEDI-Prem: telehealth for Early Developmental Intervention in babies born very preterm** | Actrn | 2021 | trial registry | | | | | Ongoing |
| **Study on language development and Intervention in late premature Infants** | ChiCtr | 2021 | trial registry | | | | | Ongoing |
| **Construction and empirical study of mindful family Participatory nursing program for premature infants** | ChiCtr | 2023 | trial registry | | | | | Ongoing |
| **influence of family-centered early intervention associated remote feedback from hospital on early development of very preterm infants** | ChiCtr | 2024 | trial registry | | | | | Ongoing |

| | | | | | | | | |
|---|---|---|---|---|---|---|---|---|
| **Impact of an integrated health, nutrition, and early child stimulation and responsive care intervention package delivered to preterm or term small for gestational age babies during infancy on growth and neurodevelopment: study protocol of an individually** | Chowdhury et al. | 2024 | trial registry | 25 | 1 | 110 | 10.1186/s13063-024-07942-z | Ongoing |
| **Effect of Integrated Nutrition, Health, Care and Support Interventions delivered to small babies during infancy, on growth and neurodevelopment** | Ctri | 2021 | trial registry | | | | | Ongoing |
| **A study comparing effect of sensory stimulation on brain which will be assessed at 4 months age among preterm neonates admitted in neonatal** | Ctri | 2023 | trial registry | | | | | Ongoing |
| **tele-education on premature's parents** | Irct20220725055554N | 2022 | trial registry | | | | | Ongoing |
| **Prodromal ImPACT Intervention for Children at Elevated Likelihood of ASD** | Nct | 2021 | trial registry | | | | | Ongoing |
| **A Randomized Control Trial of a Responsive Parenting Intervention to Support Healthy Brain Development and Self-regulation in Toddlers Born Preterm** | Nct | 2021 | trial registry | | | | | Ongoing |
| **Effects of the Newborn Behavioral Observations (NBO) as early relationship support intervention for preterm babies and their mother** | Umin | 2019 | trial registry | | | | | Ongoing |
| **Effect of Development-based Care Programs by Mothers on Growth Indices of Infants with Low Birth Weight** | Asadian et al. | 2019 | Iranian journal of neonatology | 10 | 3 | 81-87 | 10.22038/ijn.2019.35195.1532 | Author request |

| | | | | |
|---|---|---|---|---|
| **The early invention on improving survival quality of very low birth weight premature infants** | Chi, Ctr Inr | 2017 | trial registry | Author request |
| **Effects of intervention of "care for child development" for high-risk infants** | ChiCtr | 2021 | trial registry | Author request |
| **To ascertain the effectiveness of early physiotherapy intervention of infants born before due date and with low birth weight who are at high risk of developing motor developmental delay** | Ctri | 2010 | trial registry | Author request |
| **Effectiveness of an early intervention program for mothers of premature babies to care for their preemies at** | Ctri | 2017 | trial registry | Author request |
| **A clinical trial to study the effectiveness of a mobile health application for care of Preterms on the improvement of growth parameters and prevention of developmental delays of Preterm infants** | Ctri | 2018 | trial registry | Author request |
| **Evaluation of effect of early developmental physiotherapy treatment utilizing different sensory experiences versus developmental intervention to help gain head control in infants with developmental delay** | Ctri | 2018 | trial registry | Author request |
| **A study to document the long-term benefits of Kangaroo Mother Care (skin to skin contact and exclusive breast feeding) initiated at home during neonatal period on brain development at 6-7 years of child age** | Ctri | 2021 | | Author request |

| | | | | | | | | |
|---|---|---|---|---|---|---|---|---|
| **A home-based, post-discharge early intervention program promotes motor development and physical growth in the early preterm infants: a prospective, randomized controlled trial** | Fan et al. | 2021 | BMC Pediatr | 21 | 1 | 162 | https://dx.doi.org/10.1186/s12887-021-02627-x | Author request |
| **Very low birth weight infants and their families during the first year of life: comparisons of psychosocial outcomes based on after-care services** | Finello et al. | 1998 | J Perinatol | 18 | 4 | 266-71 | | Author request |
| **KMC facilitates mother baby attachment in low birth weight infants** | Gathwala et al. | 2008 | Indian J Pediatr | 75 | 1 | 43-7 | https://dx.doi.org/10.1007/s12098-008-0005-x | Author request |
| **A randomized controlled study about the use of eHealth in the home health care of premature infants** | Gund et al. | 2013 | BMC Med Inf Decis Mak | 13 | | 22 | https://dx.doi.org/10.1186/1472-6947-13-22 | Author request |
| **Family orientated early intervention enhances social-interactive behaviour of premature infants and mother-infant-interaction** | Isrctn | 2008 | trial registry | | | | | Author request |
| **Sensory integration intervention and the development of the premature infant: A controlled trial** | Lecuona et al. | 2017 | Samj, S | 107 | 11 | 976-982 | https://dx.doi.org/10.7196/SAMJ.2017.v107i11.12393 | Author request |
| **Effect of early intervention program for premature babies who were hospitalized in the Neonatal Intensive Care Unit** | n4q8v, R. B. R. | 2019 | trial registry | | | | | Author request |
| **A Longitudinal Study of Effectiveness of Early Intervention for Preterm Infants** | Nct | 2009 | trial registry | | | | | Author request |
| **Impact of Exercise on Body Composition in Premature Infants** | Nct | 2011 | trial registry | | | | | Author request |

| | | | | | | | | |
|---|---|---|---|---|---|---|---|---|
| **Effects of Family-Centered Intervention for Preterm Infants at Preschool Age** | Nct | 2015 | trial registry | | | | | Author request |
| **The Effect of a Nurse-led Continuous Support Program on Neurodevelopment of Preterm Infants** | Nct | 2017 | | | | | | Author request |
| **Family-Centered Intervention for Preterm Children: effects at School Age and Biosocial Mediators** | Nct | 2018 | trial registry | | | | | Author request |
| **A Novel Parent Education Program for Early Intervention** | Nct | 2019 | trial registry | | | | | Author request |
| **Sleep Program on Preterm Infants' Sleep, and Caregiver's Sleep, Stress, Quality of Life, and Attachment** | Nct | 2021 | | | | | | Author request |
| **The Effect of the Mobile Application Developed for Home Care of Preterm** | Nct | 2022 | | | | | | Author request |
| **Adding Motion to Contact: a New Model for Low-cost Family Centered Very-early Onset Intervention in Very Preterm-born Infants** | Nct | 2022 | trial registry | | | | | Author request |
| **Effect of holding on co-regulation in preterm infants: a randomized controlled trial** | Neu et al. | 2014 | Early Hum Dev | 90 | 3 | 141-7 | https://dx.doi.org/10.1016/j.earlhumdev.2014.01.008 | Author request |
| **Early discharge of preterm infants followed by domiciliary nursing care: parents' anxiety, assessment of infant health and breastfeeding** | Ortenstrand et al. | 2001 | Acta Paediatr | 90 | 10 | 1190-5 | https://dx.doi.org/10.1080/080352501317061639 | Author request |
| **Efficacy of the face-to-face STEP protocol compared to the remote one in infants at risk of developmental delay** | qqbykm, R. B. R. | 2024 | trial registry | | | | | Author request |

| | | | | | | | | |
|---|---|---|---|---|---|---|---|---|
| **Cognitive performance and attachment patterns at four years of age in extremely low birth weight infants after early intervention** | Sajaniemi et al. | 2001 | Eur Child Adolesc Psychiatry | 10 | 2 | 122-9 | https://dx.doi.org/10.1007/s007870170035 | Author request |
| **Motor development in premature infants: Study protocol for an interdisciplinary hospital-home intervention** | Sandoval-Cuellar et al. | 2023 | Pediatr neonatol | 64 | 5 | 577-584 | https://dx.doi.org/10.1016/j.pedneo.2022.12.015 | Author request |
| **Interdisciplinary Hospital-home Intervention on Motor Development in Premature Children** | Nct | 2020 | trial registry | | | | | Author request |
| **Intervention with African American premature infants: Four-month results of an early intervention program** | Teti et al. | 2009 | Journal of Early Intervention | 31 | 2 | 146-166 | https://dx.doi.org/10.1177/1053815109331864 | Author request |
| **A randomized controlled trial of clinic-based and home-based interventions in comparison with usual care for preterm infants: effects and mediators** | Wu, et al. | 2014 | Res Dev Disabil | 35 | 10 | 2384-93 | https://dx.doi.org/10.1016/j.ridd.2014.06.009 | Author request |
| **Early Intervention for Preterm Infants** | Nct | 2005 | trial | | | | | Author request |
| **Intervention effects on emotion regulation in preterm infants with very low birth weight: A randomize controlled trial** | Wu et al. | 2016 | Res Dev Disabil | 48 | | 1-12 | https://dx.doi.org/10.1016/j.ridd.2015.10.016 | Author request |
| **Effects of Early Intervention for Preterm Children at School Age** | Nct | 2013 | trial registry | | | | | Author request |
| **Early physiotherapy intervention in premature infants** | Yigit et al. | 2002 | Turk J Pediatr | 44 | 3 | 224-9 | | Author request |

| | | | | | | | | |
|---|---|---|---|---|---|---|---|---|
| **Family-Centered Care Enhanced Neonatal Neurophysiological Function in Preterm Infants: Randomized Controlled Trial** | Yu et al. | 2019 | Phys Ther | 99 | 12 | 1690-1702 | https://dx.doi.org/10.1093/ptj/pzz120 | Author request |
| **A Family-Centered Intervention Program for Preterm Infants: effects and Their Biosocial Pathways** | Nct | 2013 | trial registry | | | | | Author request |
| **Family-centered Care Improved Neonatal Medical and Neurobehavioral Outcomes in Preterm Infants: Randomized Controlled Trial** | Yu et al. | 2017 | Phys Ther | 97 | 12 | 1158-1168 | https://dx.doi.org/10.1093/ptj/pzx089 | Author request |
