## Supplementary material for "Effects of interdisciplinary early developmental intervention programs on behavior, executive functioning and participation in children born preterm: A systematic review with meta-analysis": S2 - Characteristics of excluded studies

### S4.1 – Predefined assessment tools

**Quality of Life**

- Toddler and Infant Questionnaire (TANDI)
- Health-Related Quality of Life (HRQoL)
- Pediatric Quality of Life Inventory (PedsQL)
- Child Health Questionnaire (CHQ)
- KIDSCREEN-9, 27, 52
- PedsQL Measurement Model
- Child Well-Being Index (OECD)
- HUI
- KINDL, SF8-36
- ILK

**Participation**

- The Child and Adolescent Scale of Participation
- Child Participation Assessment Tool
- Children Participation Questionnaire
- Questionnaire of Young People's Participation

**Parent-child interaction**

- *Quality of Caregiver-infant interaction*
    - Nursing Child Assessment Teaching Scale (NCATS)
    - Nursing Child Assessment Feeding Scale (NCAFS)
    - Emotional Availability Scale (EAS)
    - Care Index
    - Ainsworth Scale
- *Infant interaction with caregiver:*
    - Emotional Availability Scale (EAS)
    - Care Index
- *Caregiver interaction with infant:*
    - Ainsworth Scale
    - AMBIANCE
    - Care Index

### S4.2 – Assessment tools included in meta-analysis

| Assessment tool | Category | Method | Validity | Range | | Outcome direction | Description | Cite |
|---|---|---|---|---|---|---|---|---|
| | | | | Lower bound | Upper bound | | | |
| Maternal Sensitivity and Responsivity Scale (MSRS) | PCI | Coding of videotapes by professionals | Interrater reliability (Pearson's correlation coefficient):<br><br>Sensitivity 0.77<br>Undercontol 0.90<br>Overcontrol 0.70 | Likert scale: 1 (no visible signs) | Likert scale: 5 (consistent and strong signs) | Sensitivity: Higher is better<br><br>Undercontrol: Lower is better<br><br>Overcontrol: Lower is better | 3 domains:<br><br>sensitivity/responsivity scale (= consistent and appropriate behavior),<br><br>undercontrol/withdrawal scale (= disengaged behavior),<br><br>overcontrol/intrusivity scale (overstimulation, interrupting and non-consistent behavior) | [1] |
| Emotional Availability Scale (EAS) | PCI | Coding of videotapes by professionals | Interrater reliability (Cronbach's alpha):<br><br>Sensitivity 0.96,<br>Structuring 0.94,<br>Nonintrusiveness 0.82,<br>Nonhostility 0.84,<br>Child responsiveness 0.79,<br>Child involvement 0.83 | Each domain: 1 | Each domain: 7 | Higher is better | 6 domains:<br><br>adult sensitivity (warmth, perceptiveness, responsiveness to child cues),<br><br>adult structuring (appropriately guiding and scaffolding child behavior and emotions),<br><br>adult non-intrusiveness (absence of overly directive or interfering parenting),<br><br>adult non-hostility (absence of negative affect, hostility, impatience),<br><br>child responsiveness (positive affect and responsiveness to parent),<br><br>child involvement (attending to and involving the parent in the interaction). | [2] |
| Greenspan-Lieberman Observation System (GLOS) | PCI | Coding of videotapes by professionals | Overall interrater percentage of agreement 93.2-99.4% | N/A | N/A | Positive behavior: higher is better; | Coding of 5 minutes video-taped parent-child free play in 15s intervals.<br>Classification of behavior in: | [3] |

| | | | | | | | | |
|---|---|---|---|---|---|---|---|---|
| | | | | | | Negative behavior: lower is better | Positive verbal,<br>Negative verbal,<br>Positive non-verbal,<br>Negative non-verbal | |
| Interaction Rating Scale (IRS) – Parker-Loewen | PCI | Coding of videotapes by professionals | median interrater reliability: 0.88 | 1.0 | 3.0 | Higher is better | 4 scales for non-feeding related interaction;<br>4 scales for feeding related interaction | [4] |
| Interaction Rating Scale (IRS) – Silveira | PCI | Assesment by professionals | Internal consistency of each domain:<br>Cronbach's alpha 0.43-0.88<br><br>Total internal consistency:<br>Cronbach's alpha 0.85-0.91 | Behavior items:<br>0 = no<br><br>Impression items:<br>1 = not evident at all | Behavior items:<br>1 = yes<br><br>Impression items:<br>5 = evident at high level | Higher is better | 10 subscales with 70 items about behaviors:<br><br>5 subscales (25 items) - children's social skills (autonomy, responsiveness, empathy, motor regulation, and emotional regulation)<br><br>5 subscales (45 dichotomous items) - parenting skills (respect for: autonomy development, responsiveness development, empathy development, cognitive development, and social-emotional development)<br><br>overall score of synchronous interactions between mother and the child | [5, 6] |
| Nijmeegse Ouderlijke Stress Index (NOSI) | PCI | Parent questionnaire | Internal reliability of parent and child domains:<br><br>alpha coefficient >0.90 | Likert scale:<br>1<br><br>Overall:<br>0 | Likert scale:<br>6<br><br>Overall:<br>615 | Lower is better | NOSI 24 months:<br>123 items using a 6-point Likert scale<br><br>subscales related to the parent:<br>sense of competence, restriction of role,<br>attachment,<br>depression,<br>parent's health,<br>social isolation and relationship with spouse;<br><br>subscales related to the child:<br>adaptability,<br>mood,<br>distractibility/hyperactivity,<br>demandingness,<br>'reinforces parent', | [7, 8] |

| | | | | | | | | |
|---|---|---|---|---|---|---|---|---|
| | | | | | | | acceptability | |
| Parental Stress Index – Short Form (PSI-SF) | PCI | Parent questionnaire | Mother-father correlation (Pearson correlation coefficient):<br><br>0.52-0.63 | Likert scale:<br>1 (strongly disagree)<br><br>36 | Likert scale:<br>5 (strongly agree)<br><br>180 | Lower is better | 36 items using a 5-point-Likert scale;<br><br>Scores above the 85th percentile (90 raw score) are considered clinically significant. | [9] |
| Working Model of the Child Interview (WMCI) | PCA | Assesment by professionals | Interrater percentage of agreement 80%;<br>Cohen's kappa 0.56 | 16 | 90 | N/A | 6 domains items using a 5-point-Likert scale:<br><br>richness of perception,<br>openness to change,<br>intensity of involvement,<br>coherence,<br>caregiving sensitivity,<br>acceptance;<br><br>2 domains:<br>Infant difficulty,<br>fear for safety;<br><br>Classification of attachment representation in:<br>balanced (secure),<br>disengaged (insecure),<br>distorted (insecure) | [10] |
| Mother-to-Child Attachment Score (MCA) | PCA | NR | NR | NR | NR | NR | No data or source given. | [11] |
| Postpartum Bonding Questionnaire (PBQ) | PCA | Parent questionnaire | NR | 0 | 125 | Lower is better | 26 points or more indicating impaired bonding, 40 points or more indicating severe bonding disorder | [12] |
| Behavior Rating Inventory of Executive Functioning 2nd Edition (BRIEF2) | EF | Parent questionnaire | Internal consistency (alpha coefficient):<br><br>Parent forms 0.76-0.97<br>Teacher forms 0.88-0.98 | 36 | >99 | Lower is better | Standardized T-scores (mean, 50; SD, 10);<br>In clinical settings, a T-score ≥65 (≥1.5 standard deviations [SDs] from the mean, or approximately the 93rd percentile) is generally accepted as the threshold for potential difficulties on behavior rating scales | [13]<br>[14, 15] |

| | | | | | | | | |
|---|---|---|---|---|---|---|---|---|
| CNT Arrow switching | EF | Assesment by professionals | NR | N/A | N/A | Time/errors: lower is better<br><br>Self-corrections: higher is better | Sex- and age-standardized z-scores<br><br>4 subtests with increasing difficulty;<br>Performance measured by:<br>time taken for task completion,<br>errors,<br>self-corrections | [16] |
| D-KEFS Tower Test | EF | Assesment by professionals | Validity: “adequate”<br><br>Reliability: “adequate to good”<br><br>(Homack 2005) | N/A | N/A | Time/moves: lower is better | Age-standardized (mean, 10; SD, 3)<br><br>measures planning;<br>building specified patterns of towers on 3 pegs using disks in the fewest moves possible and as quickly as possible.<br><br>Scores are based on:<br>Speed,<br>solutions using the fewest moves possible | [13, 17] |
| Tower of London (TOL) | EF | Assesment by professionals | NR | 0 | 12 | Higher is better | 12 items that increase in difficulty: children moved 3 balls across 3 pegs of different heights to achieve a specific pattern while adhering to prescribed rules. There are 12 items that increase in difficulty. The number of items correctly solved (within a 60-second time limit) and the number of items correctly solved in the first attempt | [18] |
| Working Memory Test Battery (WMTB) | EF | Assesment by professionals | NR | NR | NR | Higher is better | Mean 100; SD 15<br><br>WMTB – digit recall (verbal immediate memory):<br>recall of a series of digits in forward order<br><br>WMTB – block recall (visuospatial immediate memory):<br>recalling and tapping blocks in a sequenced order as demonstrated<br><br>WMTB – backward digit recall (verbal working memory): | [13] |

| | | | | | | | | |
|---|---|---|---|---|---|---|---|---|
| | | | | | | | requires recall of a series of digits in reverse order<br><br>Score:<br>>1 SD below the normative test mean is abnormal | |
| Differential Ability Scale (DAS) | Prt | Assesment by professionals | internal reliability scores:<br>Early years 0.79 - 0.94 School age 0.74 - 0.96<br><br>Test-Retest reliability:<br>0.51-0.92<br><br>Interrater reliability:<br>0.95-0.99 | 30 | 170 | Higher is better | mean of 100 and SD of 15;<br><br>verbal composite score:<br>verbal concepts and knowledge<br><br>non-verbal composite score:<br>complex, nonverbal, inductive reasoning<br><br>spatial reasoning score:<br>complex visual processing | [19, 20] |
| Behavior Rating Scale (BRS) of the Bayley Scales of Infant Development-II (BSID-II) | B | Parent questionnaire | Internal consistency (coefficient alpha):<br>0.88<br><br>Test-retest reliability:<br>0.6 | 0 | 100 | Higher is better | Score:<br>≥26 is normal,<br>25 to 11 is questionable,<br>≤10 is nonoptimal | [21, 22] |
| Child Behavior Checklist (CBCL) | PrB, InB, ExB | Parent questionnaire | CBCL 1.5-5 (Carter 2003)<br><br>Test-retest reliability<br>r = 0.84<br><br>Mother-father agreement<br>r = 0.61<br><br>Parent-childcare agreement<br>r = 0.65<br><br>CBCL 4-18 (Achenbach 1991)<br><br>Item scores<br>(intraclass correlation coefficient): | Likert scale:<br>0<br><br>Overall:<br>0 | Likert scale:<br>2<br><br>Overall:<br>200 | Lower is better | Mean 50 with SD 10,<br>99-100 items;<br><br>Raw scores are transformed into T-scores, accounting for differences in sex and age.<br><br>T-score:<br><60 is normal,<br>60 to 63 is borderline,<br>>63 is the clinical range. | [8, 21, 23-27] |

| | | | | | | | | |
|---|---|---|---|---|---|---|---|---|
| | | | competences 0.927,<br>specific problem items 0.959<br><br>Test-retest reliability:<br>competences 0.996,<br>specific problem items 0.952<br><br>Mother-father agreement:<br>competences 0.59 – 0.87<br>specific problem items 0.65 – 0.75 | | | | | |
| Infant-Toddler Social and Emotional Assessment (ITSEA) | InB, ExB | Parent questionnaire | Internal consistency (Cronbach's alpha):<br>Externalizing 0.92<br>Internalizing 0.75<br>Dysregulation 0.78<br>Competencies 0.84<br><br>Test-retest reliability:<br>domains 0.82 - 0.90<br>scales 0.69 - 0.85<br><br>mother-father correlation (intraclass correlation):<br>domains 0.58 - 0.79<br>scales 0.43 - 0.78 | Likert scale:<br>0 | Likert scale:<br>3 | Lower is better | 4 broad domains of behavior:<br>dysregulation,<br>externalizing,<br>internalizing,<br>competencies;<br><br>3 point scale (0 = rarely/not true, 1 = somewhat true/sometime and 3 = Very true/often).<br><br>m-ITSEA: questionnaire consisting of 82 items, derived from the original 169 item | [26, 28] |
| Strength and Difficulties Questionnaire (SDQ) | AtB | Parent questionnaire | Parent-teacher correlation:<br>Total score $Chi^2 = 5.90$ | 0 | 40 | Higher is better | total difficulty score,<br>5 subscales consisting of 5 items each:<br>hyperactivity/inattention,<br>conduct problems,<br>peer problems,<br>emotional symptoms,<br>prosocial behavior;<br><br>parental score:<br>> 80th percentile is abnormal,<br><br>total difficulty score:<br>≥17 points is abnormal | [8, 29, 30] |

| | | | | | | | | |
|---|---|---|---|---|---|---|---|---|
| Test of Everyday Attention for children (TEA-Ch) | AtB | Assesment by professionals | Test-retest correlation (Pearson correlation coefficient):<br><br>Score! 0.64<br>Map Mission 0.88<br>Score Dual Task 0.74 | Score!<br>0<br><br>Map Mission<br>0<br><br>Score Dual Task<br>0 | Score<br>10<br><br>Map Mission<br>80<br><br>Score Dual Task<br>10 | Higher is better | Sex- and age-standardized (mean, 10; SD, 3)<br><br>nine subtests<br><br>Score!42: measure of sustained attention; requires counting the number of intermittent tones presented across 10 trials.<br><br>TEA-Ch Map Mission: measure of selective attention; visual cancellation task.<br><br>TEA-Ch Score Dual Task: measure of divided attention; requires counting tones while listening for target words | [31] |
