## Supplementary material for "Effects of interdisciplinary early developmental intervention programs on behavior, executive functioning and participation in children born preterm: A systematic review with meta-analysis": S3 - Predefined and included assessment tools

### Supplement S3

*Table S3.1: Summary of findings of primary outcomes*

| **Outcome** Timepoint of outcome assessment | **Assessment tool** (scale) | **No. of participants** (trials) | **Relative risk** (M-H, Random, 95% CI) | **Absolute effect** | | **Certainty of evidence** | **Comment** |
|---|---|---|---|---|---|---|---|
| | | | | Control | Early Intervention | | |
| **Behavior** | | | | | | | |
| Diagnosis of AD(H)D | | | | | | | No trials reported this outcome |
| Attention<br><br>13 years | Strengths and Difficulties Questionnaire;<br><br>Test of Everyday Attention – Children | 81 participants<br>1 trial[1] | Evidence indicates that the intervention yielded greater benefits for hyperactivity and inattentive behaviors in adolescents from lower family social risk backgrounds compared to those from higher family social risk backgrounds.<br>**SDQ:**<br>mean group difference (95% CI)<br>**-0.7 ( -2.1 to 0.7)**<br><br>Marginal intervention effects were observed for adolescent selective attention, but not for other domains of attention.<br>**TEA-Ch** Map Mission – selective attention:<br>mean group difference (95% CI)<br>**2.1 (0.2 to 4.0)** | | | **Very low**<br>due to serious risk of bias, serious indirectness[2], insufficient precision | The SDQ has also been reported in another study, but only as a total score. As all attention-related results originated from a single study, a meta-analysis was not feasible. |
| **Participation** | | | | | | | |
| Special educational needs in pre-school and early school age[1]<br><br>5-8 years<br><br>*binär* | Parent/teacher interview | 1011 participants<br>2 trials[3] | Relative risk: 0.91<br>(CI 95% 0.71 - 1.16)<br>Based on data from 1011 patients and[2] studies<br>Observation period 5-8 years | **371**<br>per 1000<br><br>Difference: 33 less per 1,000 (CI 95% 108 less - 59 more) | **338**<br>per 1000 | **Moderate**<br>due to high risk of bias[3] | EI probably has little to no effect on SEN at 5.5 and eight years CA. |
| Leisure activites | | | | | | | No trials reported this outcome. |

[1] (Stedall et al. 2022)
[2] serious indirectness because the original outcome was AD(H)D, inprecision because only one study
[3] (Van Hus et al. 2013) and (Litt et al. 2015)

*Continuation table S3.1*

| | | | | | |
|---|---|---|---|---|---|
| **Executive Functioning** | | | | | |
| Overall executive functioning<br><br>8-13 years | TOL[4],<br>BRIEF2, WMTB,<br>CNT Arrow Switching[5] | 8-yr-assessment: 100 participants;[6]<br>13-yr-assessment: 81 participants[7]<br><br>1 trial | Executive function was measured at age 8 using the Tower of London (TOL) and at age 13 using the Behavior Rating Inventory of Executive Functioning 2nd Edition (BRIEF2), the Working Memory Test Battery (WMTB), and CNT Arrow switching.<br>After implementing the VIBeS Plus early intervention program, no significant effect was found at ages 8 [Spittle 2016] or 13 [Stedall 2022. Since all results come from the same study (Spittle 2010), no meta-analysis was conducted. | **Very low**<br>due to serious risk of bias, due to very serious imprecision[22] | Since all results come from the same study (Spittle 2010), no meta-analysis was conducted. |
| **Parent-Child Interaction** | | | | | |
| Parent-Child interaction<br><br>2-12 months | Interaction Rating Scale (IRS)<br><br>Greenspan-Lieberman Observation System (GLOS) | 76 participants<br>2 trials | After implementing an early intervention program, the IRS showed no significant effect on either non-food-related or food-related interactions.[8]<br><br>However, in a study concerning a developmental program the GLOS showed a significant improvement, specifically in positive verbal interaction (+V) and negative non-verbal interaction (-NV).<br><br>Interactions per time segment: Mean (intervention/control)<br>+V: 3.03/2.40<br>-NV: 0.06/0.17[9] | **Very low**<br>due to serious risk of bias, due to very serious imprecision[21] | Due to the lack of dispersion measures, the results cannot be used for a meta-analysis. |
| Parent-Child interaction<br><br>18-24 months | Parental Stress Index – short form (PSI-SF)<br><br>Nijmeegse OuderlijkeStress Index – short version (NOSIK) | 178 participants<br>2 trials | Another intervention focused on parental education demonstrated a significant and positive effect on PSI-SF parent–child interaction at 18 months<br>(MD= -7.1; 95% CI -9.4 to -4.7).[10]<br><br>Meanwhile a family-focuesd trial could not find a significant effect using the NOSIK at 24 months CA.<br>(MD 0.00, 95%-CI -1.70 to 1.70).[11] | **Very low**<br>due to serious risk of bias, due to very serious imprecision[21] | Although both the NOSI and PSI-SF are derived from the Parental Stress Index—with the NOSI being the Dutch adaptation—their subscales differ, hence meta-analysis was not possible. |

4 (Spittle et al. 2016)
5 (Stedall et al. 2022)
6 (Spittle et al. 2016)
7 (Stedall et al. 2022)
8 (Parker-Loewen and Lytton 1987)
9 (Resnick, Armstrong, and Carter 1988)
10 (Castel et al. 2016)
11 (Meijssen et al. 2011)

*Continuation table S3.1*

| | | | | | | | |
|---|---|---|---|---|---|---|---|
| **Parent-Child Interaction** | | | | | | | |
| Parent-Child Interaction<br><br>0,5-2 years | Maternal Sensitivity and Responsivity Scales (MSRS)[12]; Emotional Availability Scales (EAS)[13] | 180 participants<br><br>2 trials | Relative risk: 0.91<br>(CI 95% 0.71 - 1.16)<br>Based on data from 1011 patients and 2 studies | **3.91**<br>Mean | **4.29**<br>Mean | **Low**<br>due to high risk of bias, and to serious imprecision | EI may increase maternal sensitivity slightly. |
| | | | | Difference: **0.38 higher (SMD)**<br>(CI 95% 0.09 higher — 0.68 higher) | | | |
| **Quality of life** | | | | | | | |
| QoL | | | | | | | No trials reported this outcome. |

[12] (Meijssen et al. 2010)
[13] (Treyvaud et al. 2022)

*Table S3.2: Summary of findings of secondary outcomes*

| **Outcome** Timepoint of outcome assessment | **Assessment tool** (scale) | **No. of participants** (trials) | **Relative risk** (M-H, Random, 95% CI) | **Absolute effect** Control | Early Intervention | **Certainty of evidence** | **Comment** |
|---|---|---|---|---|---|---|---|
| **Behaviour** | | | | | | | |
| Diagnosis of Autism Spectrum Disorder | | | | | | | No trials reported this outcome. |
| Behavioral problems in infancy<br>2-3 years | Child Behavior Checklist (CBCL) Scale<br>0-64; Lower is better | 1028 participants<br>2 studies | | **47.2** Mean | **43.56** Mean<br>Difference: **MD 3.64 smaller** (CI 95% 6.19 smaller - 1.09 smaller) | **Low** Due to very serious risk of bias[8] | . EI may reduce behavioral difficulties slightly. |
| Behavioral problems in pre-school age<br>4 years | Child Behavior Checklist (CBCL) Scale<br>0-64; Lower is better | 239 participants<br>2 studies | | **46.9** Mean | **46.68** Mean<br>Difference: **MD 0.22 smaller** (CI 95% 2.76 smaller - 2.32 larger) | **Very low** Due to very serious risk of bias and serious imprecision[10] | EI may have little to no effect on behavioral difficulties. |
| Internalizing behavioral problems in infancy<br>*2 years* | Child Behavior Checklist (CBCL) Scale; Infant-Toddler Social and Emotional Assessment (ITSEA)<br>0-64; Lower is better | 438 participants<br>4 studies | | **14.5** Mean | **14.33** Mean<br>Difference: **SMD 0.17 smaller** (CI 95% 6.19 smaller - 1.09 smaller) | **Very low** Due to very serious risk of bias and serious inconsistency[12] | EI may have little to no effect on internalizing behavioral problems in infancy. |
| Internalizing behavioral problems in pre-school age<br>4 years | Child Behavior Checklist (CBCL) Scale<br>0-64; Lower is better | 239 participants<br>2 studies | | **47.9** Mean | **44.85** Mean<br>Difference: **MD 3.05 smaller** (CI 95% 5.89 smaller - 0.22 smaller) | **Very low** Due to very serious risk of bias and serious imprecision[14] | EI may reduce internalizing behavioral problems in preschool age slightly, but evidence is very uncertain. |

*Continuation table S3.2*

| | | | | | | | |
|---|---|---|---|---|---|---|---|
| **Participation** | | | | | | | |
| Communication in pre-school age<br><br>*4 years* | Differential Ability Scale (DAS)<br><br>30 – 170;<br>Higher is better | 104 participants<br><br>1 study | Although the data are uncertain, a small positive effect may be observed in early childhood [606]. The effect is more pronounced at eight years of age in children from socially disadvantaged families [607]. | **94.9**<br>Mean<br><br>Difference: **MD 4.1 higher**<br>(CI 95% 2.2 less - 10.4 higher) | **99.0**<br>Mean | **Very low**<br>due to serious risk of bias and very serious imprecision (only one study)[16] | EI may have little to no effect on communication skills at four years, but evidence is very uncertain. |
| Mobility | | | | | | | No trials reported this outcome. |
| **Use of medical services** | | | | | | | |
| Use of paramedical medical services<br><br>5.5 years<br><br>*binary* | Parent interview | 136 participants<br><br>1 study | Relative risk: 0.88<br>(CI 95% 0.53 - 1.46) | **328**<br>per 1000<br><br>Difference: 39 less per 1,000<br>(CI 95% 154 less - 151 more) | **289**<br>per 1000 | **Very low**<br>Due to very serious risk of bias, due to serious imprecision (1 study)[4] | EI may have little to no effect on usage of physical, occupational, or speech therapy in preschool age. But evidence is very uncertain. |
| Use of psychological professions<br><br>5.5 years<br><br>*binary* | Parent interview | 136 participants<br><br>1 study | Relative risk: 1.29<br>(CI 95% 0.47 - 3.52) | **90**<br>per 1000<br><br>Difference: 26 more per 1,000<br>(CI 95%: 48 less - 227 more) | **116**<br>per 1000 | **Very low**<br>Due to very serious risk of bias, due to serious imprecision (1 study)[4] | EI may have little to no effect on usage of psychological therapy in preschool age, but evidence is very uncertain. |
