## Supplementary material for "Effects of interdisciplinary early developmental intervention programs on behavior, executive functioning and participation in children born preterm: A systematic review with meta-analysis": S4 - Summary of Findings

### Appendix: Search strategy for “Impact of early intervention programs on developmental outcomes in preterm infants: a systematic review and meta-analysis”

**<u>Ovid MEDLINE(R) ALL</u>**

1. exp Infant, Premature/
2. exp Infant, low birth weight/
3. (((premature or pre mature or preterm or pre term or prematurely) adj3 (infant* or neonat* or neo nat* or baby or babies or newborn? or born? or child or children)) or ((prematurely or preterm) adj born adolescent*)).tw.
4. ((neonatal or neo natal) adj3 (prematurity or pre maturity)).tw.
5. (low birth weight or low birthweight).tw.
6. (preemie or premie or preemies or premies).tw.
7. or/1-6
8. Early Intervention, Educational/
9. Physical Stimulation/
10. Developmental Disabilities/pc
11. ((preventative or preventive) adj care).tw.
12. (early adj2 (intervention* or program* or service* or support or stimulation or child care)).tw.
13. (intervention program* or ((infant or child*) adj2 (intervention* or program* or service* or support or stimulation))).tw.
14. ((neurobehavio?ral or neuro-behavio?ral or neurodevelopmental or neuro-developmental) adj (intervention* or therap*)).tw.
15. (development* adj2 (program* or intervention* or enrichment or support or therap* or care)).tw.
16. (behavio?ral intervention* or behavio?ral care or infant stimulation).tw.
17. (infant development or child development* or parent-infant relationship or child-centered intervention* or early detection centre* or early detection center* or play therap* or early childhood classroom* or stimulation program* or logopedic* or language stimulation or language support or speech therap* or speech-language patholog* or transaction program* or kicking intervention*).tw.
18. ((multisensory or auditory or tactile or visual or vestibular or vestibular-rocking) adj2 (intervention* or stimulation)).tw.
19. (vision school or vision services or vision support).tw.
20. (audiology service* or audiology support*).tw.
21. (family-based intervention* or family guidance or family assist* or family support or family training* or family centered or family service* or social work or environmental enrichment or discharge planning or discharge program*).tw.
22. (home intervention* or home care service* or home health nursing or home healthcare or home support or home-based support or preventive health promotion or homemaker service* or home visit* or nursing service* or public health nursing or supporting families or p?ediatric developmental assessment*).tw.
23. (((educational or educational-behavioural) adj (intervention* or program* or support or service*)) or ((parent or parental) adj1 (child interaction training* or infant relationship* or counsel?ing or advis* or support or empowerment or training* or education)) or (parenting adj (education or intervention* or skills or capacity or strateg*))).tw.
24. (((peer or psychological or community or social) adj support*) or (psychological adj (service* or therap* or counse?ling or intervention*)) or emotional counsel?ing or psychosocial support or psychotherap* or support forum).tw.

25. (healthy child program* or early years library or portage or early head start or early hearing detection intervention or portage home teaching scheme*).tw.
26. exp Physical Therapy Modalities/
27. exp Physical Therapy Specialty/
28. exp Exercise Therapy/
29. exp Exercise Movement Techniques/
30. exp Rehabilitation/
31. exp Therapeutic Touch/
32. exp Massage/
33. (physio or physical therap* or physiotherapy or physiatrics or physical exercises or physical medicine or remedial exercise*).tw.
34. (exercise* or musculoskeletal manipulation* or musculo-skeletal manipulation* or music therap* or physical therap* or physiotherapy* or rehabilitation or sensory therap* or sensory integration* or sensorimotor stimulation* or therapeutic touch).tw.
35. (movement therap* or motor training program* or motor development).tw.
36. Bobath.tw.
37. (applied physiology or body-based manipulative therap* or functional education or executive function training* or kinesitherap* or manual therap* or medical gymnastics or modality therap* or motion therap* or motor learning or neurophysiotherap* or neurological physiotherap*).tw.
38. (Margaret Rood approach or Doman-Delecto technique or Kabat-Knoss-Voss technique or Vojta approach or Vojta method or Vojta therap* or treadmill training or practice reaching or sticky mittens training*).tw.
39. (baby-massage program* or infant-massage program*).tw.
40. exp Occupational Therapy/
41. (occupational therap* or occupational rehabilitation or occupation-focused practice or occupation-based intervention or occupational medicine or ergotherap* or activity therapy or activity-based therap* or activity-based intervention or developmental programming or developmental curriculum or functional therap* or joint attention intervention*).tw.
42. or/8-41

[43-53: Cochrane Handbook 2024 RCT filter - sensitivity maximizing version]

43. randomized controlled trial.pt.
44. controlled clinical trial.pt.
45. randomi?ed.ab.
46. placebo.ab.
47. drug therapy.fs.
48. randomly.ab.
49. trial.ab.
50. groups.ab.
51. or/43-50
52. exp animals/ not humans/
53. 51 not 52
54. 7 and 42 and 53
55. remove duplicates from 54

<u>Reference for search filter:</u>

Lefebvre C, Glanville J, Briscoe S, Featherstone R, Littlewood A, Marshall C, Metzendorf MI, Noel-Storr A, Paynter R, Rader T, Thomas J, Wieland L. Searching for and selecting studies. In: Higgins J, Thomas J, Chandler J, Cumpston M, Li T, Page M, Welch V, editors. Cochrane Handbook for Systematic Reviews of Interventions [Internet] Version 6.3 (updated February 2022) ed. Cochrane; 2022. Chapter 4; Available from: https://training.cochrane.org/handbook.

**CENTRAL via Cochrane Library**

1. [mh "Infant, Premature"]
2. [mh "Infant, low birth weight"]
3. (((premature OR "pre mature" OR preterm OR "pre term" OR prematurely) NEAR/3 (infant* OR neonat* OR ("neo" NEXT nat*) OR baby OR babies OR newborn? OR born? OR child OR children)) OR ((prematurely OR preterm) NEXT ("born" NEXT adolescent*))):ti,ab,kw
4. ((neonatal OR "neo natal") NEAR/3 (prematurity OR "pre maturity")):ti,ab,kw
5. ("low birth weight" OR "low birthweight"):ti,ab,kw
6. (preemie OR premie OR preemies OR premies):ti,ab,kw
7. {or #1-#6}
8. [mh ^"Early Intervention, Educational"]
9. [mh ^"Physical Stimulation"]
10. [mh ^"Developmental Disabilities"]
11. ((preventative OR preventive) NEXT care):ti,ab,kw
12. (early NEAR/2 (intervention* OR program* OR service* OR support OR stimulation OR "child care")):ti,ab,kw
13. (("intervention" NEXT program*) OR ((infant OR child*) NEAR/2 (intervention* OR program* OR service* OR support OR stimulation))):ti,ab,kw
14. ((neurobehavioral OR neuro-behavioral OR neurobehavioural OR neuro-behavioural OR neurodevelopmental OR neuro-developmental) NEXT (intervention* OR therap*)):ti,ab,kw
15. (development* NEAR/2 (program* OR intervention* OR enrichment OR support OR therap* OR care)):ti,ab,kw
16. ((behavioral NEXT intervention*) OR (behavioural NEXT intervention*) OR (behavioral NEXT "care") OR (behavioural NEXT "care") OR "infant stimulation"):ti,ab,kw
17. ("infant development" OR ("child" NEXT development*) OR "parent-infant relationship" OR ("child-centered" NEXT intervention*) OR ("early detection" NEXT centre*) OR ("early detection" NEXT center*) OR ("play" NEXT therap*) OR ("early childhood" NEXT classroom*) OR ("stimulation" NEXT program*) OR logopedic* OR "language stimulation" OR "language support" OR ("speech" NEXT therap*) OR ("speech-language" NEXT patholog*) OR ("transaction" NEXT program*) OR ("kicking" NEXT intervention*)):ti,ab,kw
18. ((multisensory OR auditory OR tactile OR visual OR vestibular OR vestibular-rocking) NEAR/2 (intervention* OR stimulation)):ti,ab,kw
19. ("vision school" OR "vision services" OR "vision support"):ti,ab,kw
20. (("audiology" NEXT service*) OR ("audiology" NEXT support*)):ti,ab,kw
21. (("family-based" NEXT intervention*) OR "family guidance" OR ("family" NEXT assist*) OR "family support" OR ("family" NEXT training*) OR "family centered" OR ("family" NEXT service*) OR "social work" OR "environmental enrichment" OR "discharge planning" OR ("discharge" NEXT program*)):ti,ab,kw
22. (("home" NEXT intervention*) OR ("home care" NEXT service*) OR "home health nursing" OR "home healthcare" OR "home support" OR "home-based support" OR "preventive health promotion" OR ("homemaker" NEXT service*) OR ("home" NEXT visit*) OR ("nursing" NEXT service*) OR "public health nursing" OR "supporting families" OR (p?ediatric NEXT "developmental" NEXT assessment*)):ti,ab,kw
23. (((educational OR educational-behavioural) NEXT (intervention* OR program* OR support OR service*)) OR ((parent OR parental) NEAR/1 (("child interaction" NEXT training*) OR ("infant" NEXT relationship*) OR counsel?ing OR advis* OR support OR empowerment OR training* OR education)) OR (parenting NEXT (education OR intervention* OR skills OR capacity OR strateg*))):ti,ab,kw
24. (((peer OR psychological OR community OR social) NEXT support*) OR (psychological NEXT (service* OR therap* OR counse?ling OR intervention*)) OR ("emotional" NEXT counsel?ing) OR "psychosocial support" OR psychotherap* OR "support forum"):ti,ab,kw

25. (("healthy child" NEXT program*) OR "early years library" OR portage OR "early head start" OR "early hearing detection intervention" OR ("portage home teaching" NEXT scheme*)):ti,ab,kw
26. [mh "Physical Therapy Modalities"]
27. [mh "Physical Therapy Specialty"]
28. [mh "Exercise Therapy"]
29. [mh "Exercise Movement Techniques"]
30. [mh Rehabilitation]
31. [mh "Therapeutic Touch"]
32. [mh Massage]
33. (physio OR ("physical" NEXT therap*) OR physiotherapy OR physiatrics OR "physical exercises" OR "physical medicine" OR ("remedial" NEXT exercise*)):ti,ab,kw
34. (exercise* OR ("musculoskeletal" NEXT manipulation*) OR ("musculo-skeletal" NEXT manipulation*) OR ("music" NEXT therap*) OR ("physical" NEXT therap*) OR physiotherapy* OR rehabilitation OR ("sensory" NEXT therap*) OR ("sensory" NEXT integration*) OR ("sensorimotor" NEXT stimulation*) OR "therapeutic touch"):ti,ab,kw
35. (("movement" NEXT therap*) OR ("motor training" NEXT program*) OR "motor development"):ti,ab,kw
36. Bobath:ti,ab,kw
37. ("applied physiology" OR ("body-based manipulative" NEXT therap*) OR "functional education" OR ("executive function" NEXT training*) OR kinesitherap* OR ("manual" NEXT therap*) OR "medical gymnastics" OR ("modality" NEXT therap*) OR ("motion" NEXT therap*) OR "motor learning" OR neurophysiotherap* OR ("neurological" NEXT physiotherap*)):ti,ab,kw
38. ("Margaret Rood approach" OR "Doman-Delecto technique" OR "Kabat-Knoss-Voss technique" OR "Vojta approach" OR "Vojta method" OR ("Vojta" NEXT therap*) OR "treadmill training" OR "practice reaching" OR ("sticky mittens" NEXT training*)):ti,ab,kw
39. (("baby-massage" NEXT program*) OR ("infant-massage" NEXT program*)):ti,ab,kw
40. [mh "Occupational Therapy"]
41. (("occupational" NEXT therap*) OR "occupational rehabilitation" OR "occupation-focused practice" OR "occupation-based intervention" OR "occupational medicine" OR ergotherap* OR "activity therapy" OR ("activity-based" NEXT therap*) OR "activity-based intervention" OR "developmental programming" OR "developmental curriculum" OR ("functional" NEXT therap*) OR ("joint attention" NEXT intervention*)):ti,ab,kw
42. {or #8-#41}
43. #7 AND #42
44. #7 AND #42 in Trials

**<u>APA PsycInfo (Ovid)</u>**

1. Premature Birth/
2. (((premature or pre mature or preterm or pre term or prematurely) adj3 (infant* or neonat* or neo nat* or baby or babies or newborn? or born? or child or children)) or ((prematurely or preterm) adj born adolescent*)).tw.
3. ((neonatal or neo natal) adj3 (prematurity or pre maturity)).tw.
4. (low birth weight or low birthweight).tw.
5. (preemie or premie or preemies or premies).tw.
6. or/1-5
7. Early Intervention/
8. exp Early Childhood Development/
9. exp Physical Development/
10. ((preventative or preventive) adj care).tw.

11. (early adj2 (intervention* or program* or service* or support or stimulation or child care)).tw.
12. (intervention program* or ((infant or child*) adj2 (intervention* or program* or service* or support or stimulation))).tw.
13. ((neurobehavio?ral or neuro-behavio?ral or neurodevelopmental or neuro-developmental) adj (intervention* or therap*)).tw.
14. (development* adj2 (program* or intervention* or enrichment or support or therap* or care)).tw.
15. (behavio?ral intervention* or behavio?ral care or infant stimulation).tw.
16. (infant development or child development* or parent-infant relationship or child-centered intervention* or early detection centre* or early detection center* or play therap* or early childhood classroom* or stimulation program* or logopedic* or language stimulation or language support or speech therap* or speech-language patholog* or transaction program* or kicking intervention*).tw.
17. ((multisensory or auditory or tactile or visual or vestibular or vestibular-rocking) adj2 (intervention* or stimulation)).tw.
18. (vision school or vision services or vision support).tw.
19. (audiology service* or audiology support*).tw.
20. (family-based intervention* or family guidance or family assist* or family support or family training* or family centered or family service* or social work or environmental enrichment or discharge planning or discharge program*).tw.
21. (home intervention* or home care service* or home health nursing or home healthcare or home support or home-based support or preventive health promotion or homemaker service* or home visit* or nursing service* or public health nursing or supporting families or p?ediatric developmental assessment*).tw.
22. (((educational or educational-behavioural) adj (intervention* or program* or support or service*)) or ((parent or parental) adj1 (child interaction training* or infant relationship* or counsel?ing or advis* or support or empowerment or training* or education)) or (parenting adj (education or intervention* or skills or capacity or strateg*))).tw.
23. (((peer or psychological or community or social) adj support*) or (psychological adj (service* or therap* or counse?ling or intervention*)) or emotional counsel?ing or psychosocial support or psychotherap* or support forum).tw.
24. (healthy child program* or early years library or portage or early head start or early hearing detection intervention or portage home teaching scheme*).tw.
25. exp Physical Therapy/
26. exp Exercise Therapy/
27. exp Exercise/
28. exp Rehabilitation/
29. Massage/
30. (physio or physical therap* or physiotherapy or physiatrics or physical exercises or physical medicine or remedial exercise*).tw.
31. (exercise* or musculoskeletal manipulation* or musculo-skeletal manipulation* or music therap* or physical therap* or physiotherapy* or rehabilitation or sensory therap* or sensory integration* or sensorimotor stimulation* or therapeutic touch).tw.
32. (movement therap* or motor training program* or motor development).tw.
33. Bobath.tw.
34. (applied physiology or body-based manipulative therap* or functional education or executive function training* or kinesitherap* or manual therap* or medical gymnastics or modality therap* or motion therap* or motor learning or neurophysiotherap* or neurological physiotherap*).tw.
35. (Margaret Rood approach or Doman-Delecto technique or Kabat-Knoss-Voss technique or Vojta approach or Vojta method or Vojta therap* or treadmill training or practice reaching or sticky mittens training*).tw.

36. (baby massage or infant massage).tw.
37. Occupational Therapy/
38. (occupational therap* or occupational rehabilitation or occupation-focused practice or occupation-based intervention or occupational medicine or ergotherap* or activity therapy or activity-based therap* or activity-based intervention or developmental programming or developmental curriculum or functional therap* or joint attention intervention*).tw.
39. or/7-38
40. 6 and 39

[41: Eady et al. 2008 "PsycInfo Search Strategies" filter – best sensitivity version]

41. (control* or random*).tw. or exp Treatment/
42. 40 and 41
43. remove duplicates from 42

<u>Reference for search filter:</u>

Eady AM, Wilczynski NL, Haynes RB. PsycINFO search strategies identified methodologically sound therapy studies and review articles for use by clinicians and researchers. J Clin Epidemiol. 2008 Jan;61(1):34-40. doi: 10.1016/j.jclinepi.2006.09.016. PMID: 18083460.

**<u>CINAHL (Ebsco)</u>**

1. (MH "Infant, Premature")
2. (MH " Infant, Low Birth Weight+")
3. ((((TI premature OR AB premature) OR (TI "pre mature" OR AB "pre mature") OR (TI preterm OR AB preterm) OR (TI "pre term" OR AB "pre term") OR (TI prematurely OR AB prematurely)) N3 ((TI infant* OR AB infant*) OR (TI neonat* OR AB neonat*) OR (TI "neo nat*" OR AB "neo nat*") OR (TI baby OR AB baby) OR (TI babies OR AB babies) OR (TI newborn# OR AB newborn#) OR (TI born# OR AB born#) OR (TI child OR AB child) OR (TI children OR AB children))) OR (((TI prematurely OR AB prematurely) OR (TI preterm OR AB preterm)) W1 (TI "born adolescent*" OR AB "born adolescent*")))
4. (((TI neonatal OR AB neonatal) OR (TI "neo natal" OR AB "neo natal")) N3 ((TI prematurity OR AB prematurity) OR (TI "pre maturity" OR AB "pre maturity")))
5. ((TI "low birth weight" OR AB "low birth weight") OR (TI "low birthweight" OR AB "low birthweight"))
6. ((TI preemie OR AB preemie) OR (TI premie OR AB premie) OR (TI preemies OR AB preemies) OR (TI premies OR AB premies))
7. S1 OR S2 OR S3 OR S4 OR S5 OR S6
8. (MH "Early Intervention+")
9. (MH "Physical Stimulation+")
10. (MH "Developmental Disabilities")
11. (((TI preventative OR AB preventative) OR (TI preventive OR AB preventive)) W1 (TI care OR AB care))
12. ((TI early OR AB early) N2 ((TI intervention* OR AB intervention*) OR (TI program* OR AB program*) OR (TI service* OR AB service*) OR (TI support OR AB support) OR (TI stimulation OR AB stimulation) OR (TI "child care" OR AB "child care")))
13. ((TI "intervention program*" OR AB "intervention program*") OR (((TI infant OR AB infant) OR (TI child* OR AB child*)) N2 ((TI intervention* OR AB intervention*) OR (TI program* OR AB program*) OR (TI service* OR AB service*) OR (TI support OR AB support) OR (TI stimulation OR AB stimulation))))

14. (((TI neurobehavio#ral OR AB neurobehavio#ral) OR (TI neuro-behavio#ral OR AB neuro-behavio#ral) OR (TI neurodevelopmental OR AB neurodevelopmental) OR (TI neuro-developmental OR AB neuro-developmental)) W1 ((TI intervention* OR AB intervention*) OR (TI therap* OR AB therap*)))
15. ((TI development* OR AB development*) N2 ((TI program* OR AB program*) OR (TI intervention* OR AB intervention*) OR (TI enrichment OR AB enrichment) OR (TI support OR AB support) OR (TI therap* OR AB therap*) OR (TI care OR AB care)))
16. ((TI "behavio#ral intervention*" OR AB "behavio#ral intervention*") OR (TI "behavio#ral care" OR AB "behavio#ral care") OR (TI "infant stimulation" OR AB "infant stimulation"))
17. ((TI "infant development" OR AB "infant development") OR (TI "child development*" OR AB "child development*") OR (TI "parent-infant relationship" OR AB "parent-infant relationship") OR (TI "child-centered intervention*" OR AB "child-centered intervention*") OR (TI "early detection centre*" OR AB "early detection centre*") OR (TI "early detection center*" OR AB "early detection center*") OR (TI "play therap*" OR AB "play therap*") OR (TI "early childhood classroom*" OR AB "early childhood classroom*") OR (TI "stimulation program*" OR AB "stimulation program*") OR (TI logopedic* OR AB logopedic*) OR (TI "language stimulation" OR AB "language stimulation") OR (TI "language support" OR AB "language support") OR (TI "speech therap*" OR AB "speech therap*") OR (TI "speech-language patholog*" OR AB "speech-language patholog*") OR (TI "transaction program*" OR AB "transaction program*") OR (TI "kicking intervention*" OR AB "kicking intervention*"))
18. (((TI multisensory OR AB multisensory) OR (TI auditory OR AB auditory) OR (TI tactile OR AB tactile) OR (TI visual OR AB visual) OR (TI vestibular OR AB vestibular) OR (TI vestibular-rocking OR AB vestibular-rocking)) N2 ((TI intervention* OR AB intervention*) OR (TI stimulation OR AB stimulation)))
19. ((TI "vision school" OR AB "vision school") OR (TI "vision services" OR AB "vision services") OR (TI "vision support" OR AB "vision support"))
20. ((TI "audiology service*" OR AB "audiology service*") OR (TI "audiology support*" OR AB "audiology support*"))
21. ((TI "family-based intervention*" OR AB "family-based intervention*") OR (TI "family guidance" OR AB "family guidance") OR (TI "family assist*" OR AB "family assist*") OR (TI "family support" OR AB "family support") OR (TI "family training*" OR AB "family training*") OR (TI "family centered" OR AB "family centered") OR (TI "family service*" OR AB "family service*") OR (TI "social work" OR AB "social work") OR (TI "environmental enrichment" OR AB "environmental enrichment") OR (TI "discharge planning" OR AB "discharge planning") OR (TI "discharge program*" OR AB "discharge program*"))
22. ((TI "home intervention*" OR AB "home intervention*") OR (TI "home care service*" OR AB "home care service*") OR (TI "home health nursing" OR AB "home health nursing") OR (TI "home healthcare" OR AB "home healthcare") OR (TI "home support" OR AB "home support") OR (TI "home-based support" OR AB "home-based support") OR (TI "preventive health promotion" OR AB "preventive health promotion") OR (TI "homemaker service*" OR AB "homemaker service*") OR (TI "home visit*" OR AB "home visit*") OR (TI "nursing service*" OR AB "nursing service*") OR (TI "public health nursing" OR AB "public health nursing") OR (TI "supporting families" OR AB "supporting families") OR (TI "p#ediatric developmental assessment*" OR AB "p#ediatric developmental assessment*"))
23. ((((TI educational OR AB educational) OR (TI educational-behavioural OR AB educational-behavioural)) W1 ((TI intervention* OR AB intervention*) OR (TI program* OR AB program*) OR (TI support OR AB support) OR (TI service* OR AB service*))) OR (((TI parent OR AB parent) OR (TI parental OR AB parental)) N1 ((TI "child interaction training*" OR AB "child interaction training*") OR (TI "infant relationship*" OR AB "infant relationship*") OR (TI counsel#ing OR AB counsel#ing) OR (TI advis* OR AB advis*) OR (TI support OR AB support) OR (TI empowerment OR AB empowerment) OR (TI training* OR AB training*) OR (TI education OR AB education))) OR ((TI parenting OR AB parenting) W1 ((TI education OR AB education) OR (TI intervention* OR AB

intervention*) OR (TI skills OR AB skills) OR (TI capacity OR AB capacity) OR (TI strateg* OR AB strateg*))))

24. ((((TI peer OR AB peer) OR (TI psychological OR AB psychological) OR (TI community OR AB community) OR (TI social OR AB social)) W1 (TI support* OR AB support*)) OR ((TI psychological OR AB psychological) W1 ((TI service* OR AB service*) OR (TI therap* OR AB therap*) OR (TI counse#ling OR AB counse#ling) OR (TI intervention* OR AB intervention*))) OR (TI "emotional counsel#ing" OR AB "emotional counsel#ing") OR (TI "psychosocial support" OR AB "psychosocial support") OR (TI psychotherap* OR AB psychotherap*) OR (TI "support forum" OR AB "support forum"))
25. ((TI "healthy child program*" OR AB "healthy child program*") OR (TI "early years library" OR AB "early years library") OR (TI portage OR AB portage) OR (TI "early head start" OR AB "early head start") OR (TI "early hearing detection intervention" OR AB "early hearing detection intervention") OR (TI "portage home teaching scheme*" OR AB "portage home teaching scheme*"))
26. (MH "Physical Therapy+")
27. (MH "Exercise+")
28. (MH "Therapeutic Exercise+")
29. (MH Rehabilitation+)
30. (MH "Therapeutic Touch")
31. (MH Massage+)
32. ((TI physio OR AB physio) OR (TI "physical therap*" OR AB "physical therap*") OR (TI physiotherapy OR AB physiotherapy) OR (TI physiatrics OR AB physiatrics) OR (TI "physical exercises" OR AB "physical exercises") OR (TI "physical medicine" OR AB "physical medicine") OR (TI "remedial exercise*" OR AB "remedial exercise*"))
33. ((TI exercise* OR AB exercise*) OR (TI "musculoskeletal manipulation*" OR AB "musculoskeletal manipulation*") OR (TI "musculo-skeletal manipulation*" OR AB "musculo-skeletal manipulation*") OR (TI "music therap*" OR AB "music therap*") OR (TI "physical therap*" OR AB "physical therap*") OR (TI physiotherapy* OR AB physiotherapy*) OR (TI rehabilitation OR AB rehabilitation) OR (TI "sensory therap*" OR AB "sensory therap*") OR (TI "sensory integration*" OR AB "sensory integration*") OR (TI "sensorimotor stimulation*" OR AB "sensorimotor stimulation*") OR (TI "therapeutic touch" OR AB "therapeutic touch"))
34. ((TI "movement therap*" OR AB "movement therap*") OR (TI "motor training program*" OR AB "motor training program*") OR (TI "motor development" OR AB "motor development"))
35. (TI Bobath OR AB Bobath)
36. ((TI "applied physiology" OR AB "applied physiology") OR (TI "body-based manipulative therap*" OR AB "body-based manipulative therap*") OR (TI "functional education" OR AB "functional education") OR (TI "executive function training*" OR AB "executive function training*") OR (TI kinesitherap* OR AB kinesitherap*) OR (TI "manual therap*" OR AB "manual therap*") OR (TI "medical gymnastics" OR AB "medical gymnastics") OR (TI "modality therap*" OR AB "modality therap*") OR (TI "motion therap*" OR AB "motion therap*") OR (TI "motor learning" OR AB "motor learning") OR (TI neurophysiotherap* OR AB neurophysiotherap*) OR (TI "neurological physiotherap*" OR AB "neurological physiotherap*"))
37. ((TI "Margaret Rood approach" OR AB "Margaret Rood approach") OR (TI "Doman-Delecto technique" OR AB "Doman-Delecto technique") OR (TI "Kabat-Knoss-Voss technique" OR AB "Kabat-Knoss-Voss technique") OR (TI "Vojta approach" OR AB "Vojta approach") OR (TI "Vojta method" OR AB "Vojta method") OR (TI "Vojta therap*" OR AB "Vojta therap*") OR (TI "treadmill training" OR AB "treadmill training") OR (TI "practice reaching" OR AB "practice reaching") OR (TI "sticky mittens training*" OR AB "sticky mittens training*"))
38. ((TI "baby-massage program*" OR AB "baby-massage program*") OR (TI "infant-massage program*" OR AB "infant-massage program*"))
39. (MH "Occupational Therapy+")
40. ((TI "occupational therap*" OR AB "occupational therap*") OR (TI "occupational rehabilitation" OR AB "occupational rehabilitation") OR (TI "occupation-focused practice" OR AB "occupation-focused practice") OR (TI "occupation-based intervention" OR AB "occupation-based intervention") OR (TI

"occupational medicine" OR AB "occupational medicine") OR (TI ergotherap* OR AB ergotherap*) OR (TI "activity therapy" OR AB "activity therapy") OR (TI "activity-based therap*" OR AB "activity-based therap*") OR (TI "activity-based intervention" OR AB "activity-based intervention") OR (TI "developmental programming" OR AB "developmental programming") OR (TI "developmental curriculum" OR AB "developmental curriculum") OR (TI "functional therap*" OR AB "functional therap*") OR (TI "joint attention intervention*" OR AB "joint attention intervention*"))

41. S8 OR S9 OR S10 OR S11 OR S12 OR S13 OR S14 OR S15 OR S16 OR S17 OR S18 OR S19 OR S20 OR S21 OR S22 OR S23 OR S24 OR S25 OR S26 OR S27 OR S28 OR S29 OR S30 OR S31 OR S32 OR S33 OR S34 OR S35 OR S36 OR S37 OR S38 OR S39 OR S40
42. S7 AND S41

*[43: Wong et al. 2006 "therapy studies" filter - SDSSGS version]*

43. MH "treatment outcomes+" OR MH "experimental studies+" or random*
44. S42 AND S43

**WHO International Clinical Trials Registry Platform (ICTRP)**

((prematur* OR "pre mature" OR preterm OR "pre term" OR "low birth weight" OR "low birthweight") AND (infant* OR neonat* OR "neo nat*" OR bab* OR newborn* child OR children)) AND ("early intervention*" OR "preventative care" OR "preventive care" OR "intervention program*" OR "development* OR stimulation OR logopedi* OR logopaedi* OR language OR speech OR multisensory OR sensory OR sensorimotor OR auditory OR tactile OR visual OR vestibular OR vision OR audiology OR family OR "social work" OR enrichment OR discharge OR home OR education* OR relationship* OR counseling OR empowerment OR support OR training* OR skills OR capacity OR strateg* OR psychological service* OR psychological therap* OR psychological intervention* OR psychotherap* OR "physical therap*" OR physiotherap* OR exercise* OR rehabilitation OR "therapeutic touch" OR "movement therap*" OR motor OR "manual therap*" OR "modality therap*" OR "motion therap*" OR neurophysiotherap* OR massage OR "occupational therap*" OR ergotherap* OR "activity therap*" OR "functional therap*")
